# Prevalence of uncoupling protein one genetic polymorphisms and their relationship with cardiovascular and metabolic health

**DOI:** 10.1101/2021.06.09.21258571

**Authors:** Petros C. Dinas, Eleni Nintou, Maria Vliora, Paraskevi Sakellariou, Anna E. Pravednikova, Agata Witkowicz, Zaur M. Kachaev, Victor V. Kerchev, Svetlana N. Larina, James Cotton, Anna Kowalska, Paraskevi Gkiata, Alexandra Bargiota, Zaruhi A. Khachatryan, Anahit A. Hovhannisyan, Mariya A. Antonosyan, Sona Margaryan, Anna Partyka, Pawel Bogdanski, Monika Szulinska, Matylda Kregielska-Narozna, Rafał Czepczyński, Marek Ruchała, Anna Tomkiewicz, Levon Yepiskoposyan, Lidia Karabon, Yulii Shidlovskii, George S. Metsios, Andreas D. Flouris

## Abstract

The contribution of *UCP1* single nucleotide polymorphisms (SNPs) to susceptibility for cardiometabolic pathologies (CMP) and their involvement in specific risk factors for these conditions varies across populations. We tested whether *UCP1* SNPs A-3826G, A-1766G, Ala64Thr and A-112C are associated with the most common CMP (cardiovascular disease, hypertension, metabolic syndrome, and type-2 diabetes) and CMP risk factors. This case-control study included blood sample collection from 2,283 Caucasians (1,139 healthy; 1,144 CMP) across Armenia, Greece, Poland, Russia and United Kingdom for genotyping of the above-mentioned SNPs. We extended the results via a systematic review and meta-analysis, covering PubMed, Embase, and Cochrane Library databases. In Armenia the GA genotype and A allele of Ala64Thr were associated with ∼2-fold higher risk for CMP compared to the GG genotype or G allele, respectively (p<0.05). In Greece, A allele of Ala64Thr SNP decreased the risk of CMP by 39%. Healthy individuals with A-3826G GG genotype and carriers of mutant allele of A-112C and Ala64Thr had higher body mass index compared to those carrying other genotypes. In healthy Polish, higher waist-to-hip ratio (WHR) was observed in heterozygotes A-3826G compared to AA homozygotes. Heterozygosity of the A-112C and Ala64Thr SNPs was related to lower WHR in CMP individuals compared to wild type homozygotes (p<0.05). Meta-analysis in case-control studies showed no statistically significant odds ratios in different alleles across the four studied SNPs (p>0.05). Thus, we conclude that the studied SNPs could be associated with the most common CMP and their risk factors in some populations.

## 1. INTRODUCTION

Single nucleotide polymorphisms (SNPs) in a number of candidate genes are highly implicated in energy balance as well as fat and glucose metabolism, modifying disease susceptibility (1-3). One of these candidate genes codes for uncoupling protein 1 (UCP1) which is expressed predominantly in brown adipose tissue, holding a critical role in oxidative phosphorylation and overall energy balance (4, 5). More than 2300 SNPs have been recognized within the *UCP1* gene and its regulatory regions (6), but four have been commonly studied for their impact on metabolism and energy balance (7-11). These are: (i) A-3826G (rs1800592) located on the upstream region of *UCP1*, (ii) A-1766G (rs3811791) a 2kb upstream variant, (iii) A-112C (rs10011540) on the 5’UTR region, and (iv) Ala64Thr (rs45539933) a missense variant.

The four *UCP1* SNPs have been associated with a number of cardio-metabolic pathologies (CMP) (12). The G allele of A-3826G is more common in obese individuals (13, 14) and it is associated with increased body mass index (BMI), percent body fat, blood pressure (15), and high-density lipoprotein level (16). The same allele of this SNP is associated with higher BMI and glucose levels in overweight persons (17) and can increase the risk for proliferative diabetic retinopathy in individuals with type 2 diabetes (18). The other three SNPs are less prevalent but have been also associated with various risk factors for CMP (6, 7): C allele of the A-112C is more frequent in individuals with type 2 diabetes than in healthy individuals (19), the Ala64Thr mutant allele is associated with higher waist-to-hip ratio (WHR) (20), while the A-1766G SNP is related with obesity (9). Finally, the GAA haplotype (A-3826G, A-1766G, and Ala64Thr) is associated with decreased abdominal fat tissue, body fat mass, and WHR (21).

The contribution of the four *UCP1* SNPs to the susceptibility for CMP as well as their involvement in specific risk factors for these conditions varies across populations, even within the same race, probably due to environmental impacts. For instance, the frequency of AG of A-3826G in persons with CMP ranges from 24% in Italy (22), to around 50% in Colombia (10), Japan (19), and Korea (16), and to 85% in China (18). Similarly wide frequency ranges have been reported also for the other three SNPs across different populations (11, 23-25). At the same time, some studies report that *UCP1* SNPs are strongly associated with disease risk (9, 18, 26), while others report no such findings (27-29). Therefore, it remains unclear if differences in the prevalence of these four *UCP1* SNPs across different populations are associated with the prevalence of CMP.

Our incomplete understanding about the potential involvement of these four *UCP1* SNPs in disease susceptibility limits the potential for precision medicine to effectively address CMP. An even more direct effect on disease mitigation is that CMP risk factors are currently addressed with equal importance across different populations, because clinicians remain unaware of the link between the four *UCP1* SNPs and cardio-metabolic health in their country. In turn, this undermines the sustainability of healthcare systems due to limited efficacy of CMP prevention and mitigation guidelines. To address these important knowledge gaps, we investigated if differences in the frequency of A-3826G, A-1766G, Ala64Thr and A-112C SNPs across five countries (Armenia, Greece, Poland, Russia, United Kingdom) are associated with the most common CMP and their risk factors. To confirm any observed associations between the studied *UCP1* SNPs and cardio-metabolic health, we extended our findings to consider all previously-studied populations by conducting a systematic review and meta-analysis.

## 2. METHODS

### 2.1. Case-control study

This is a multicenter, multinational study conducted across five countries (Armenia, Greece, Poland, Russia, and United Kingdom) and funded by the European Commission (see Funding section). The participants were recruited via online and paper advertisements as well as word of mouth. Following approval from the relevant Bioethics Review Board in each country (see Online Supplement section 1.1.1). Written informed consent for participation was signed by the volunteers following detailed explanation of all the procedures and risks involved.

#### 2.1.1. Study design and data collection

The study involved two groups of participants: individuals with CMP as well as healthy controls. We considered the following CMP, as they present with the highest prevalence (30, 31) amongst all health abnormalities related to cardio-metabolic health: cardiovascular disease, hypertension, metabolic syndrome, and type 2 diabetes. The inclusion criteria were: 1) adult; 2) diagnosed presence of CMP for the CMP group and generally healthy for the control group; 3) non-smokers, or have quit smoking for at least one year; 4) not in a pregnancy or lactation period; 5) no history of eating disorders; 6) no acute illness and/or infection during the last four weeks.

All participants were assessed for: 1) medical history via a structured interview-based questionnaire; 2) anthropometry (body height, body mass, WHR); 3) percent fat mass via non-invasive bioelectrical impedance analysis; 4) genotypes of the aforementioned four *UCP1* SNPs detected in DNA isolated from blood samples. A detailed description of the adopted blood handling and genotyping methodologies is provided in the Online Supplement (Section 1.1.2). All participants were instructed, for 12 hours prior to assessments, to avoid the consumption of food, coffee, or alcohol and to refrain from exercise. Also, they were advised to consume two glasses of water about two hours prior to their assessment.

#### 2.1.2. Statistical analysis

The data were analyzed using a general genetic model as previously described (32, 33). We calculated Hardy-Weinberg equilibrium to ensure unbiased outcomes (34). Linkage disequilibrium between genetic loci, haplotype analysis, and allele frequencies estimation were performed via the SHEsis platform (35, 36). We used chi-square tests to determine differences in *UCP1* SNPs between groups, as well as Phi indices to report effect sizes (37). Also, we calculated odds ratios (OR) to determine associations of genotypes and alleles between groups in the overall sample as well as based on country (Online Supplement, Section 1.1.3). Finally, we used Kruskal Wallis ANOVA with post hoc Mann-Whitney *U* tests to assess differences in BMI, WHR, and fat percentage between genotype groups for each *UCP1* SNP. The level of statistical significance for the Hardy-Weinberg equilibrium was set at p<0.05 and for all other analyses at p≤0.05. Unless stated otherwise, the SPSS 26.0 (SPSS Inc., Chicago, IL, USA) software was used to perform the statistical analyses.

### 2.2. Systematic review and meta-analysis

We conducted a systematic review and meta-analysis (PROSPERO review protocol: CRD42019132376) investigating if differences in the frequency of A-3826G, A-1766G, Ala64Thr and A-112C SNPs are associated with the prevalence of the studied CMP. Following the Preferred Reporting Items for Systematic Reviews and Meta-analyses (PRISMA) guidelines (38), we searched the titles and abstracts in PubMed central, Embase, and Cochrane Library (trials) databases from the date of their inception to February 23, 2021, for studies that evaluated the prevalence of *UCP1* A-3826G, A-1766G, Ala64Thr and A-112C SNPs and their association with CMP. No date, participants’ health status, language, or study design limits were applied. A detailed description of the systematic review methodology and the searching algorithm is provided in the Online Supplement (Section 2.1).

## 3. RESULTS

### 3.1. Case-control study

The study population included 2283 Caucasian individuals (Table 1). Our Hardy-Weinberg equilibrium (HWE) analysis for the A-1766G revealed significant deviation in healthy individuals (χ^2^ = 33.34, p<0.001), indicating that this SNP should be excluded from further analysis (34), for other *UCP1* SNPs no deviation from HWE in healthy individuals was noticed. The frequencies of alleles and genotypes for the studied *UCP1* SNPs in healthy controls and in CMP individuals are shown in Figure 1, Table 2 and Tables S4-S11. Odds ratios for the association between genotype and health status (i.e., healthy vs. CMP individuals) for each of the four studied *UCP1* SNPs are shown in Table 2 and Tables S10-S11.

**Table 1.** Characteristics of the studied population. Age and BMI are presented as median (Q1, Q3)

|  | Group | (n) / (%) | Males / Females<br>(n) | Age (years) | BMI (kg/m<sup>2</sup>) |
| --- | --- | --- | --- | --- | --- |
| Entire sample | Healthy | 1139 / 50 | 762 / 528 | 45 (32,54) | 25.5 (23.9,26.9) |
|  | CMP | 1144 / 50 | 397 / 521 | 59 (50,65) | 30.5 (27.4,34.2) |
| Armenia | Healthy | 105 / 32 | - | - | - |
|  | CMP | 226 / 68 | 98 / 128 | 59 (54,64) | 29.0 (27.2,31.7) |
| Greece | Healthy | 233 / 47 | 131 / 102 | 55 (50,65) | 26.8 (24.2,29.9) |
|  | CMP | 264 / 53 | 125 / 139 | 62 (56,68) | 31.7 (28.9,34.5) |
| Poland | Healthy | 365 / 59 | 221 / 144 | 32 (25,44) | 23.8 (22.0,25.6) |
|  | CMP | 252 / 41 | 89 / 163 | 62 (54.7,67) | 31.2 (29.4,33.8) |
| Russia | Healthy | 255 / 45 | 142 / 113 | 46 (36.5,54.5) | 25.9 (25.3,26.3) |
|  | CMP | 310 / 55 | 129 / 181 | 52 (40,63) | 28.9 (26.0,34.6) |
| UK | Healthy | 181 / 66 | 140 / 41 | 43 (30,51) | 25.7 (23.2,29.8) |
|  | CMP | 92 / 34 | 54 / 38 | 54 (48,57) | 30.7 (25.9,38.4) |

Key: CMP = cardio-metabolic pathologies; BMI = body mass index; n = number of individuals tested. Q=quartile)

**Table 2.**
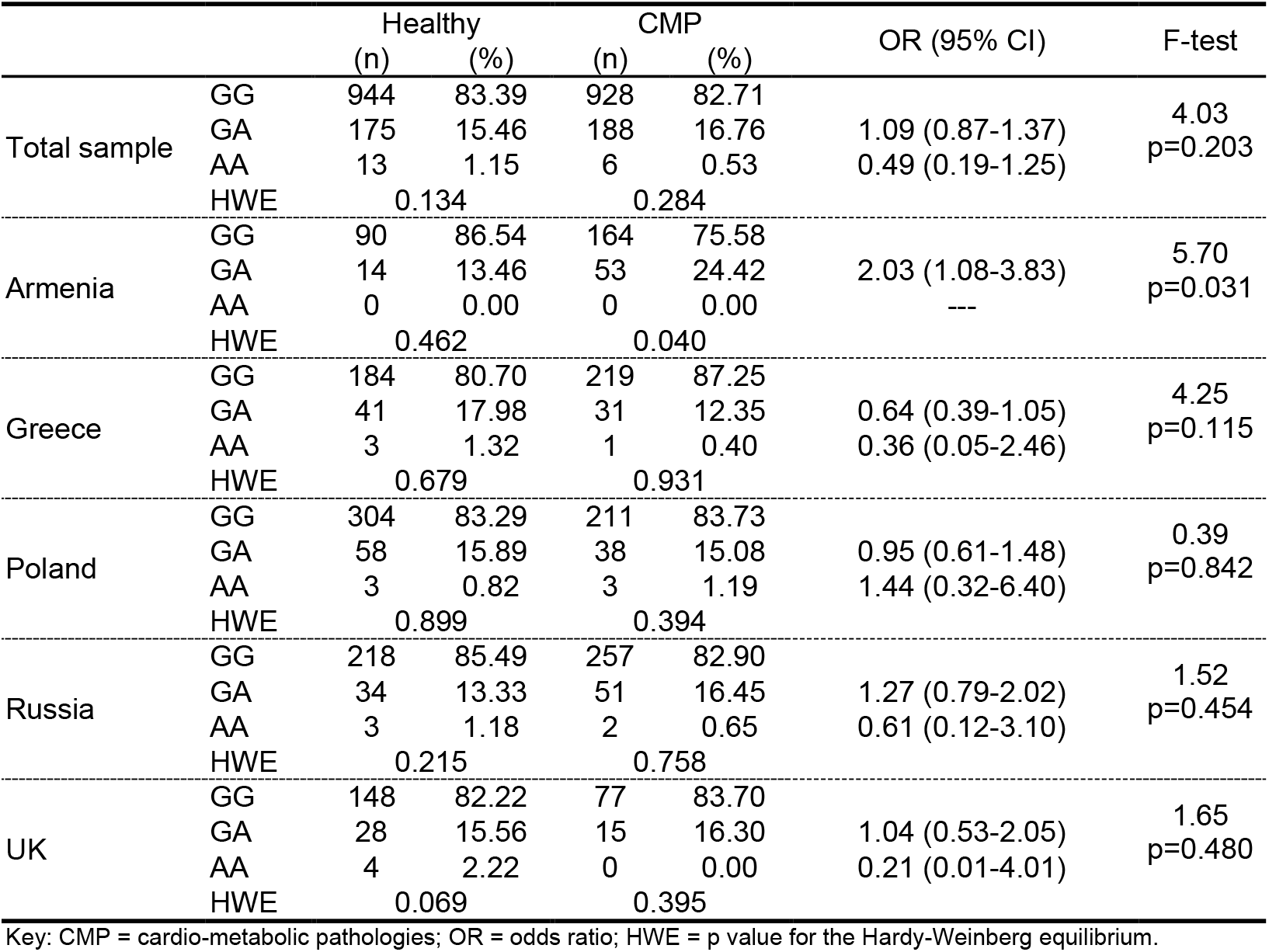
Frequency of genotypes for Ala64Thr in CMP and healthy individuals.

**Figure 1.**
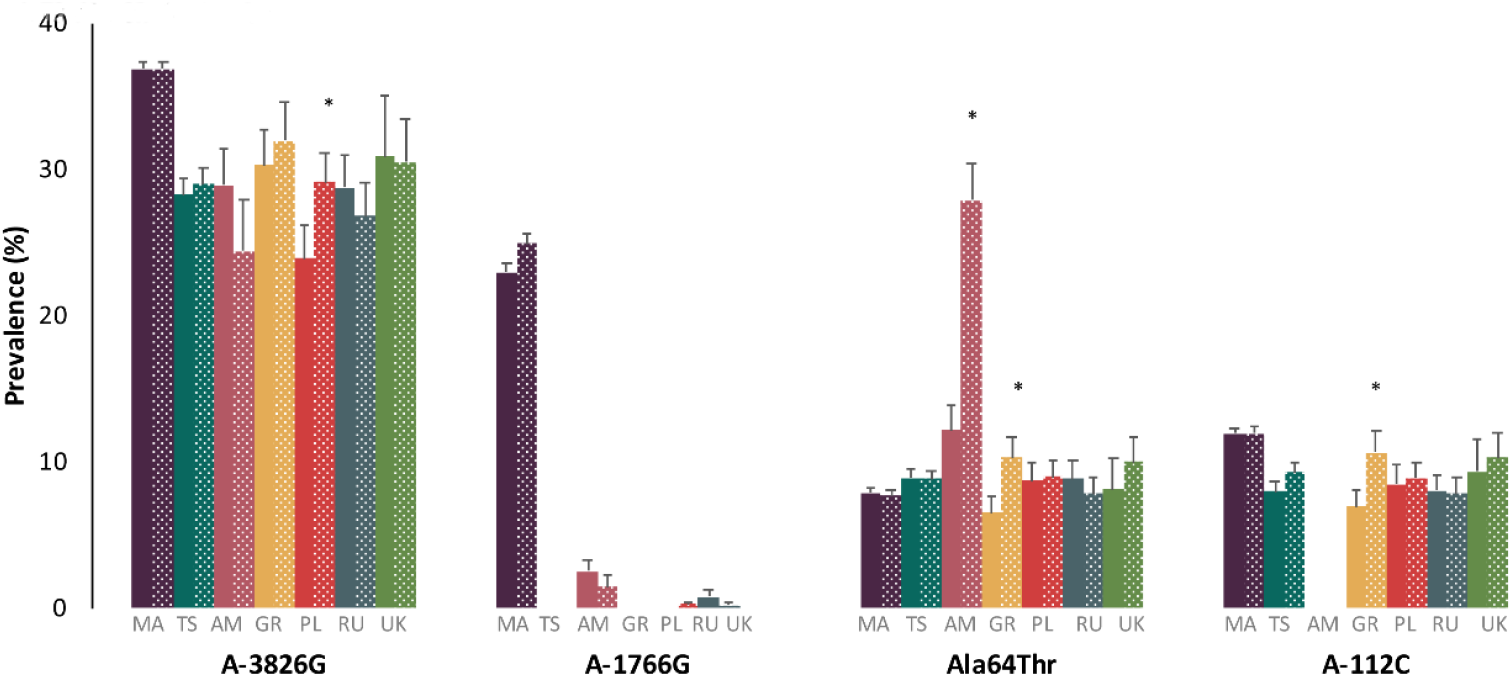
Prevalence of the studied UCP1 SNP alleles. Note: solid color bars indicate results for individuals with CMP; patterned bars indicate results for healthy persons; * indicates differences from CMP persons significant at p<0.05. Key: MA= meta-analysis, TS= total sample, AM= Armenia, GR= Greece, PL= Poland, RU= Russia, UK= United Kingdom

With regard to country-level stratification, allele frequency analysis (Tables S4-S9) in the Greek population showed that individuals carrying the C allele of the A-112C SNP or the A allele of the Ala64Thr SNP are 37% and 39% less likely to develop CMP, respectively (p<0.05; Table S6). Moreover, the G allele of the A-3826G SNP was associated with 23% lower risk to develop CMP in the Polish population (Table S7).

In total, we found no associations between genotype and health status in the overall sample for the studied *UCP1* SNPs (p>0.05). Though, we observed an association between genotype and health status for Ala64Thr within the Armenian population, where the GA genotype was carried by 24.4% of the CMP individuals but only by 13.5% of healthy individuals. Also, the GA genotype of Ala64Thr showed a 2-fold higher risk (p=0.03) for CMP than the GG genotype in the Armenian population (Table 2).

#### 3.1.2. Linkage Disequilibrium

Our analysis for all four SNPs in this study in CMP individuals and healthy controls showed that the A-3826G and Ala64Thr were in strong linkage disequilibrium with a D’ value of 0.831. Similar results were observed for the combinations of A-3826G and A-112C, as well as for the Ala64Thr and A-112C which were in strong linkage disequilibrium with D’ values of 0.917 and 0.924, respectively. However, the r^2^ values for the combinations of A-3826G and Ala64Thr (r^2^=0.165) as well as A-3826G and A-112C (r^2^=0.195) were relatively low, indicating that their effects are independent of each other. In contrast, the r^2^ value for Ala64Thr and A-112C was high (r^2^=0.848), indicating a direct link between these two SNPs. Country-specific analysis of linkage disequilibrium between investigated SNPs can be found in Figures S1-2.

#### 3.1.3. Haplotype analysis

In the overall sample, the haplotype analysis revealed that CMP individuals were 24% less likely to carry the GAC (A-3826G, Ala64Thr, A-112C) haplotype compared to healthy controls (6.5% vs 8.5%; p=0.023; Table S1). Country-specific analysis showed lower CMP risk for this haplotype across countries but this association reached statistical significance only in the Greek population (OR=0.56, CI95%: 0.34-0.91, p=0.017). Additionally, in the Polish population, we found a higher frequency of the AGA haplotype in CMP individuals compared to healthy persons (74.9% vs 70.6%), which indicates the relationship between this haplotype and higher risk of CMP (OR=1.33, CI95%: 1.03-1.73, p=0.032). On the contrary, for GGA haplotype we found a lower frequency in CMP Polish population compared to healthy individuals (15.6% vs 20.3%) indicating a protective effect in healthy individuals (OR=0.74, CI95%: 0.55-0.99, p=0.047). In the Armenian population, the AA haplotype (A-3826G, Ala64Thr) increased the CMP risk more than 4-fold (OR=4.10, CI95%: 1.12-14.98, p=0.02), while the AG haplotype decreased the susceptibility to CMP (OR=0.65, CI95%=0.45-0.95, p=0.025). The AA haplotype differs from the AG in the second position defined by the mutant allele of Ala64Thr confirming the association of A allele of this SNP with CMP risk. Detailed results for haplotype analysis for each country are provided in Tables S1-2.

#### 3.1.4. Association between UCP1 SNPs with specific CMP risk factors

In healthy individuals, we observed significantly higher BMI in the homozygotes GG of A-3826G as compared to AA and AG individuals (p=0.03) as well as in carriers of the mutant allele of A-112C (p=0.015), and Ala64Thr (p=0.004) compared to the wild type homozygotes (Table 3). We also showed that CMP individuals being heterozygotes of A-112C and Ala64Thr had lower WHR than wild type homozygotes (Table 3). Country-specific analysis showed that in the healthy Greek population, heterozygous individuals of A-112C and Ala64Thr displayed higher BMI and fat mass compared to the wild type homozygotes (BMI p=0.005, body fat p=0.008 and BMI p=0.002, body fat p=0.005, respectively; Table S14). In the Polish healthy population, mutant homozygotes of the A-112C SNP presented higher BMI compared to heterozygotes and wild type homozygotes (Table S12; p<0.05). Due to linkage disequilibrium between A-112C and Ala64Thr, the same effect was observed for mutant homozygotes of Ala64Thr. Finally, in Polish healthy individuals, higher WHR was observed in GA heterozygotes (p=0.03) in comparison to wild type homozygous subjects (Table S12).

**Table 3.** Body mass index and waist-to-hip ratio [median (Q1,Q3)] across the different *UCP1* SNPs for the entire sample as well as across healthy controls and individuals with CMP.

| SNP | Genotype | Body mass index |  | Waist-to-hip ratio |  |
| --- | --- | --- | --- | --- | --- |
|  |  | Healthy | CMP | Healthy | CMP |
| A-3826G | AA | 25.6 (23.5,26.6) | 30.3 (27.4,34.1)<sup>1</sup> | 0.87 (0.81,0.93) | 0.97 (0.92,1.04)<sup>1</sup> |
|  | AG | 25.4 (23.6,27.0) | 30.7 (27.5,34.2)<sup>1</sup> | 0.88 (0.81,0.93) | 1.00 (0.92,1.04)<sup>1</sup> |
|  | GG | 26.2 (24.1,28.7)<sup>2,3</sup> | 30.8 (27.2,33.8)<sup>1</sup> | 0.88 (0.80,0.92) | 1.00 (0.92,1.05)<sup>1</sup> |
| A-112C | AA | 25.4 (23.5,26.7) | 30.6 (27.5,34.2)<sup>1</sup> | 0.87 (0.81,0.93) | 0.98 (0.93,1.04)<sup>1</sup> |
|  | AC | 25.9 (23.7,28.3)<sup>2</sup> | 31.2 (27.3,34.2)<sup>1</sup> | 0.88 (0.82,0.94) | 0.96 (0.87,1.02)<sup>1,2</sup> |
|  | CC | 26.3 (25.5,27.2) | 27.9 (27.3,32.5)<sup>1</sup> | 0.87 (0.85,0.89) | 0.94 (0.84,1.00) |
| Ala64Thr | GG | 25.4 (23.4,26.7)<sup>2,3</sup> | 30.5 (27.4,34.1)<sup>1</sup> | 0.87 (0.81,0.93) | 0.98 (0.93,1.04)<sup>1,3</sup> |
|  | GA | 26.0 (23.8,28.3) | 30.5 (27.3,33.7)<sup>1</sup> | 0.88 (0.82,0.93) | 0.97 (0.87,1.03)<sup>1</sup> |
|  | AA | 26.3 (26.1,27.4) | 29.8 (27.2,32.7) | 0.90 (0.84,0.98) | 0.92 (0.80,1.02) |

Note: <sup>1</sup> = difference from healthy significant at p≤0.05; <sup>2</sup> = difference from AA significant at p≤0.05; <sup>3</sup> = difference from AG significant at p≤0.05. Key: CMP = cardio-metabolic pathologies

### 3.2. Systematic review and meta-analysis

#### 3.2.1. Searching procedure

The searching procedure retrieved 817 publications of which 109 were duplicates. We excluded 219 publications being reviews, editorials, and conference proceeding as well as 161 publications which referred to animal studies. From the 328 remaining publications, 276 were excluded as they did not meet the inclusion criteria. In total, 52 eligible publications were included in the analysis. Detailed searching procedure results can be found in a PRISMA flowchart (Figure S3).

#### 3.2.2. Characteristics of included studies and risk of bias assessment

The 52 eligible publications included in the analysis were published between 1998 and 2020 and included data from 24 different countries. The extracted data for all 52 included publications can be found in Table S17. The risk of bias assessment demonstrated low risk for the vast majority of the eligible studies (Online Supplement Section 2.2).

#### 3.2.3. Meta-analysis outcomes

Fifty-one out of the 52 eligible publications were used for prevalence meta-analyses, while 22 eligible publications were used for odds ratios meta-analyses. The results from the meta-analyses are summarized in Figure 1 and Table 4, while the SNP-specific forest and funnel plots for the prevalence (Figures S5-24, S35-44) and the odds ratios (Figures S25-34, S45-S49) can be found in the Online Supplement (Sections 2.2.1 and 2.2.2). On the whole, for the different genotypes and alleles we performed 24 prevalence meta-analyses and 12 odds ratios meta-analyses which included a total of 34,313 cases. No statistically significant differences were observed in the prevalence of the mutant alleles of the four different SNPs (p>0.05; Figure 1). Also, when we considered only case-control studies, we found no statistically significant odds ratios in different alleles across the four studied SNPs (p>0.05).

**Table 4.**
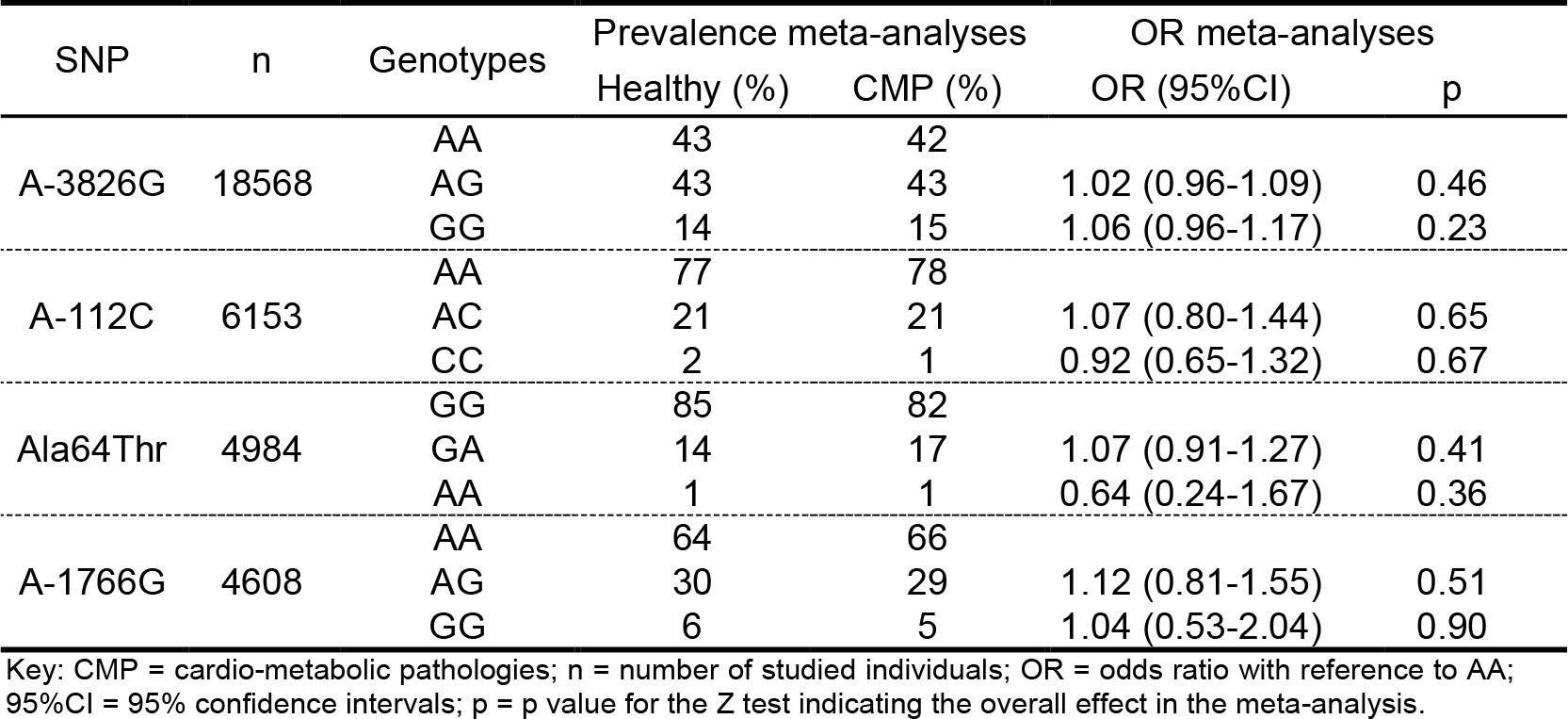
Meta-analysis results for the prevalence and odds ratios of genotypes of the four different SNPs, between healthy and CMP individuals.

## 4. DISCUSSION

Our findings confirm an association between the studied *UCP1* SNPs and cardio-metabolic health in a multi-country sample of 2,283 persons as well as in a systematic review and meta-analysis that considers all previously-studied populations, a total of 34 thousand people across 24 countries. Furthermore, we found that differences in the distribution of genotypes and alleles of the studied SNPs between CMP individuals and healthy controls are associated with the prevalence of one or more of the most common CMP and their risk factors, in some (Armenia, Greece, and Poland) but not all (Russia and United Kingdom) countries.

Within our study population, the A-3826G (AG) was the most prevalent of the four SNPs studied in persons with CMP was 40%, ranging from 34% in the UK to 42% in Armenia and 42% in Russia. This is very similar to the 43% found in our meta-analysis, and mid-way between the 29% reported in Spain (15) and the ≈50% reported in Colombia (10), Japan (19), and Korea (16). Our findings in the case-control study indicate that the A-3826G is not associated with CMP, but that it leads to increased BMI within the healthy population. Thus, it may promote the development of CMP in the presence of environmental factors (39) as well as other genetic traits (40).

Our results for Ala64Thr and A-112C indicate a strong linkage disequilibrium between the two SNPs. In our study the mutant A allele of Ala64Thr was detected in 9% of both healthy individuals and persons with CMP, and this frequency was not very different across the five studied countries. This was similar to the 7% for healthy individuals and 9% for CMP individuals found in our meta-analysis that included data from 4,984 persons across nine countries. Our observed prevalence rates for the C allele of A-112C were 9% in healthy persons and 8% in individuals with CMP. This was somewhat lower than the 12% prevalence found in our meta-analysis that included data from 6,153 persons across eight countries. In terms of health impacts, our results demonstrate that the Ala64Thr and A-112C are associated with opposing effects in healthy individuals and persons with CMP. Specifically, we found that healthy individuals carrying the mutant alleles display higher BMI and, in some countries, body fat percent. On the other hand, persons with CMP who carry the mutant variants have lower WHR. These results partly reflect those reported in previous studies (21, 41). For instance, the presence of mutant alleles Ala64Thr and A-1766G, in combination with A-3826G, can augment the beneficial effects of caloric restriction resulting in greater reductions in WHR (21).

Our findings indicate potential limitations of common analysis of different races, ethnicities, and regions when analyzing our data as an entire sample or via meta-analytic methods. For instance, the frequency of A allele of Ala64Thr across all our studied countries was 9%, similar to the 8% found in our meta-analysis, in both cases suggesting no differences between healthy persons and individuals with CMP. However, our country-specific analysis demonstrated that the prevalence of A allele of Ala64Thr was significantly higher in healthy individuals across the Armenian (27.9%) and the Greek (10.3%) populations, as compared to CMP persons. Considering risk factors, we detected a number of associations with the four studied SNPs across Greece, Armenia and Poland, which were not observed in the other countries. Taken together, these findings suggest that the studied SNPs may be important for promoting risk factors and pathophysiological mechanisms involved in CMP, but that this involvement may be stronger in some races, ethnicities, and/or regions.

We conclude that, in some populations, the A-3826G, A-1766G, Ala64Thr and A-112C SNPs *UCP1* SNPs may be associated with the prevalence of one or more of the most common CMP and their risk factors. Future studies on these SNPs may shed more light on the genetics of CMP and may uncover potential candidates for precision medicine.

## Data Availability

The authors confirm that the data supporting the findings of this study are available within the article [and/or] its supplementary materials

https://www.crd.york.ac.uk/PROSPERO/display_record.php?RecordID=132376

## ACKNOWLEDGMENTS

The authors are grateful to Monika Jasek, Marta Wagner, and Eleftheria Barmpa for their support during the data collection and analysis.

