## Supplementary materials for "Prevalence of uncoupling protein one genetic polymorphisms and their relationship with cardiovascular and metabolic health"

**1**. **CASE-CONTROL STUDY**

**1.1 Materials and Methods**

**1.1.1 *Bioethics approval procedures***

This multicenter, multinational study conducted across Armenia, Greece, Poland, Russia, and United Kingdom, received approval from the relevant Bioethics Review Board in each country:

1. Armenia: Institute of Molecular Biology, National Academy of Sciences of Republic of Armenia, ref. No IRB/IEC: IRB00004079, (IORG 0003427)/3-7-2012;
2. Greece: Department of Physical Education and Sport Science, University of Thessaly, ref. No 610/9-7-2012.
3. Poland: Local Research Bioethics Committee, University of Medical Sciences, Poznan, ref. No KB 215/13 revised KB 85/16;
4. Russia: Institute of Gene Biology, Russian Academy of Sciences, ref. no 12318-308/ 6-7-2012;
5. United Kingdom: National Health Services/Manchester East Research Ethics Committee, ref. No 15/NW/0874/12-1-2016.

**1.1.2*. Blood handling and genotyping***

1.1.2.a Greece and United Kingdom

We collected 4ml of whole blood in EDTA anti-coagulated vacutainers. Genomic DNA was extracted from 4ml whole blood using a NucleoSpin blood QuickPure kit (Macherey-Nagel). Concentration and purity of isolated DNA were evaluated with Qubit 2.0 fluorometer (ThermoFisher Scientific) and Qubit dsDNA BR Assay Kit (ThermoFisher Scientific) and samples were stored at -20 °C until the day of the genotyping analysis. DNA samples (10 ng) were genotyped using the TaqMan SNP genotyping assays (ThermoFisher Scientific) for the *UCP1* A-3826G (rs1800592), A-1766G (rs10011540), A-112C (rs3811791), and Ala64Thr (rs45539933) polymorphisms. Reactions were conducted in 384-well plates in a reaction volume of 10 ul using 1X TaqMan Universal Master Mix II (ThermoFisher Scientific) and 1X TaqMan assay (ThermoFisher Scientific). The plates were then placed in a real-time PCR thermal cycler (ViiA7 Real-Time PCR System; ThermoFisher Scientific), and thermal cycling conditions were as follow: incubation at 60˚C for 1 min and 95 ˚C for 10 min, followed by 40 cycles of 95˚C for 15 seconds and 60˚C for 1 min. Fluorescence data files from each plate were analysed using automated allele-calling software (QuantStudio 7 Real-Time PCR Software v1.3; ThermoFisher Scientific).

1.1.2.b. Armenia

We collected 4ml of whole blood in EDTA anti-coagulated vacutainers. 4 ml of blood sample was added in a 15 ml Falcon tube and centrifuged at 3,000 rpm for 10 min. After removal of supernatant 14 ml of RBC lysis buffer was added and centrifuged at 3,000 rpm for 10 min until a clear white pellet was obtained. 25 µl of Proteinase K (20 mkg/ml) and 5 ml of WBC lysis buffer were added and the pellet was disturbed. The tubes were incubated at 56ºC for 2.5-3 hours. 2 ml of 6 M NaCl was added, the tubes were shaking periodically for 20 minutes and centrifuged at 3,000 rpm for 30 min until a clear supernatant and rigid pellet were obtained. To precipitate DNA aqueous phase was added into the 50 ml transparent clear tubes with 35-40 ml 96% ethanol. The tubes were gently shacked until the DNA medusa was generated. Transferred in 1.5 ml sterile Eppendorf tubes, the DNA medusa was washed in 1 ml 70% ethanol. After drying 400 µl TE buffer (pH=8.0) was added and the tubes with DNAs were put at 37ºC overnight to solve DNAs. Concentration and purity of isolated DNA were evaluated with the ratio of absorbance at 260 nm and 280 nm and DNA samples were stored at -20 °C until the day of the genotyping analysis. Four SNPs in the *UCP1* gene, rs1800592, rs3811791, rs45539933, rs10011540 were genotyped using the TaqManSNP Genotyping Assays, respectively: [C___8866368_20](https://www.thermofisher.com/order/genome-database/details/genotyping/C___8866368_20?CID=&ICID=&subtype=), [C___2052379_10](https://www.thermofisher.com/order/genome-database/details/genotyping/C___2052379_10?CID=&ICID=&subtype=), [C___25619416_30](https://www.thermofisher.com/order/genome-database/details/genotyping/C__25619416_30?CID=&ICID=&subtype=), [C___25761748_10](https://www.thermofisher.com/order/genome-database/details/genotyping/C__25761748_10?CID=&ICID=&subtype=). Amplification reactions (10μl/well) were carried out in 96-well plate with 20 ng of template DNA, 5 μl TaqMan Genotyping Master Mix (ThermoFisher Scientific), 0.2μl of appropriate, for tested SNP, fluorogenic probe and 3.8 μl MiliQ water. An initial denaturing step of 10 min at 95°C was followed by 40 cycles of 15 seconds at 95°C, 1 min at 60°C. Reaction was performer on Viia 7 System (Applied Biosystems), the results were analysed using QuantStudio™ Software V1.2.4, ThermoFisher Scientific. Moreover, accuracy of genotyping of *UCP1* SNPs: rs1800592, rs3811791 and rs45539933 was verified by PCR-restriction fragment length polymorphism (PCR-RFLP) method.

1.1.2.c. Poland

Peripheral blood samples were obtained from 252 individuals with CMP (mean age 59.68 ± 11.37, 163 female/89 male) and 365 healthy, unrelated volunteers (35.19 ± 12.64, 144 female/221 male). Individuals were diagnosed at the Department of Internal Diseases, Poznań University of Medical Science, Poland. Blood samples from healthy volunteers were collected in cooperation with the Regional Center for Blood Donation and Blood Treatment in Wrocław. Genomic DNA was extracted from 3 ml whole blood using Invisorb Spin Blood Midi Kit (*Stratec Molecular GmbH*) according to the manufacturer’s protocol. The DNA concentration and purity (A_260/280_) was determined spectrophotometrically (*Denovix*). All samples were stored at -20°C. Four SNPs in the *UCP1* gene, rs1800592, rs3811791, rs45539933, rs10011540 were genotyped using the TaqManSNP Genotyping Assays, respectively: [C___8866368_20](https://www.thermofisher.com/order/genome-database/details/genotyping/C___8866368_20?CID=&ICID=&subtype=), [C___2052379_10](https://www.thermofisher.com/order/genome-database/details/genotyping/C___2052379_10?CID=&ICID=&subtype=), [C___25619416_30](https://www.thermofisher.com/order/genome-database/details/genotyping/C__25619416_30?CID=&ICID=&subtype=), [C___25761748_10](https://www.thermofisher.com/order/genome-database/details/genotyping/C__25761748_10?CID=&ICID=&subtype=). Amplification reactions (10μl/well) were carried out in 96-well plate with 20 ng of template DNA, 5 μl TaqMan Genotyping Master Mix (ThermoFisher Scientific), 0.2μl of appropriate, for tested SNP, fluorogenic probe and 3.8 μl MiliQ water. An initial denaturing step of 10 min at 95°C was followed by 40 cycles of 15 s at 95°C, 1 min at 60°C. Reaction was performer on Viia 7 System (Applied Biosystems), the results were analysed using QuantStudio™ Software V1.2.4, ThermoFisher Scientific. Moreover, accuracy of genotyping of *UCP1* SNPs: rs1800592, rs3811791 and rs45539933 was verified by PCR-restriction fragment length polymorphism (PCR-RFLP) method.

1.1.2.d. Russia

We collected 4ml of whole blood in EDTA anti-coagulated vacutainers. Genomic DNA was extracted from 0.2 ml whole blood using a GeneJET Genomic DNA Purification Kit (ThermoFisher Scientific). Concentration of isolated DNA was evaluated with Qubit 2.0 fluorometer (ThermoFisher Scientific) and Qubit dsDNA BR Assay Kit (ThermoFisher Scientific). Purity of DNA was assessed based on the ratio of absorbance at 260 nm and 280 nm using an Eppendorf Biospecrometer. Samples were stored at -20 °C until the day of the genotyping analysis. DNA samples (100 ng) were genotyped using polymerase chain reaction with TaqMan probes and primers for the *UCP1* A-3826G (rs1800592), A-1766G (rs3811791), A-112C (rs10011540), and Ala64Thr (rs45539933) polymorphisms. Primers and probes were designed using Primer Express Software (version 3.0; Applied Biosystems). Reactions were conducted in 96-well plates in a reaction volume of 25 ul using 1X Taq Buffer (Evrogen), 1.25 u HS Taq DNA Polymerase (Evrogen), 1 mM dNTPs (ThermoFisher Scientific), 2.5-6 mM (depends on SNP) MgCl_2_, 0.4 μM of each primer, 0.16 μM of each probe, DNA sample, and mQ water. The plates were then placed in a real-time PCR thermal cycler (CFX96 Touch Real-Time PCR Detection System; BIO-RAD), and thermal cycling conditions were as follow: incubation at 60˚C for 1 min and 95 ˚C for 10 min, followed by 40 cycles of 95˚C for 15 seconds and 58-60˚C (depends on SNP) for 1 min. Fluorescence data files from each plate were analyzed using automated allele-calling software (CFX Maestro Software; BIO-RAD). To perform the quality control of the genotyping method, we assessed PCR-products of randomly chosen samples from each genotype by direct sequencing.

**1.1.3*. Statistical analysis***

Prevalence rates for each SNP were calculated for: (1) the overall sample size, (2) each country, c) health status (i.e. CMP and healthy). Prevalence was determined by dividing the presence of genotype/allele of each SNP by the overall sample size. Standard error of the prevalence was calculated with the following formula: a (presence of genotype/allele) / [a (presence of genotype/allele) * b (sample size)]^2^. The odds ratio (OR) for the analyzed genotype/allele was determined with the following equation: OR= [a (sample size of CMP individuals) * b (presence of genotype/allele in CMP individuals)] / [c (sample size of healthy participants) * d (presence of genotype/allele in healthy participants)].

**1.2. Results (Tables and Figures)**

**Figure S1:** Linkage Disequilibrium heat maps for the overall sample size

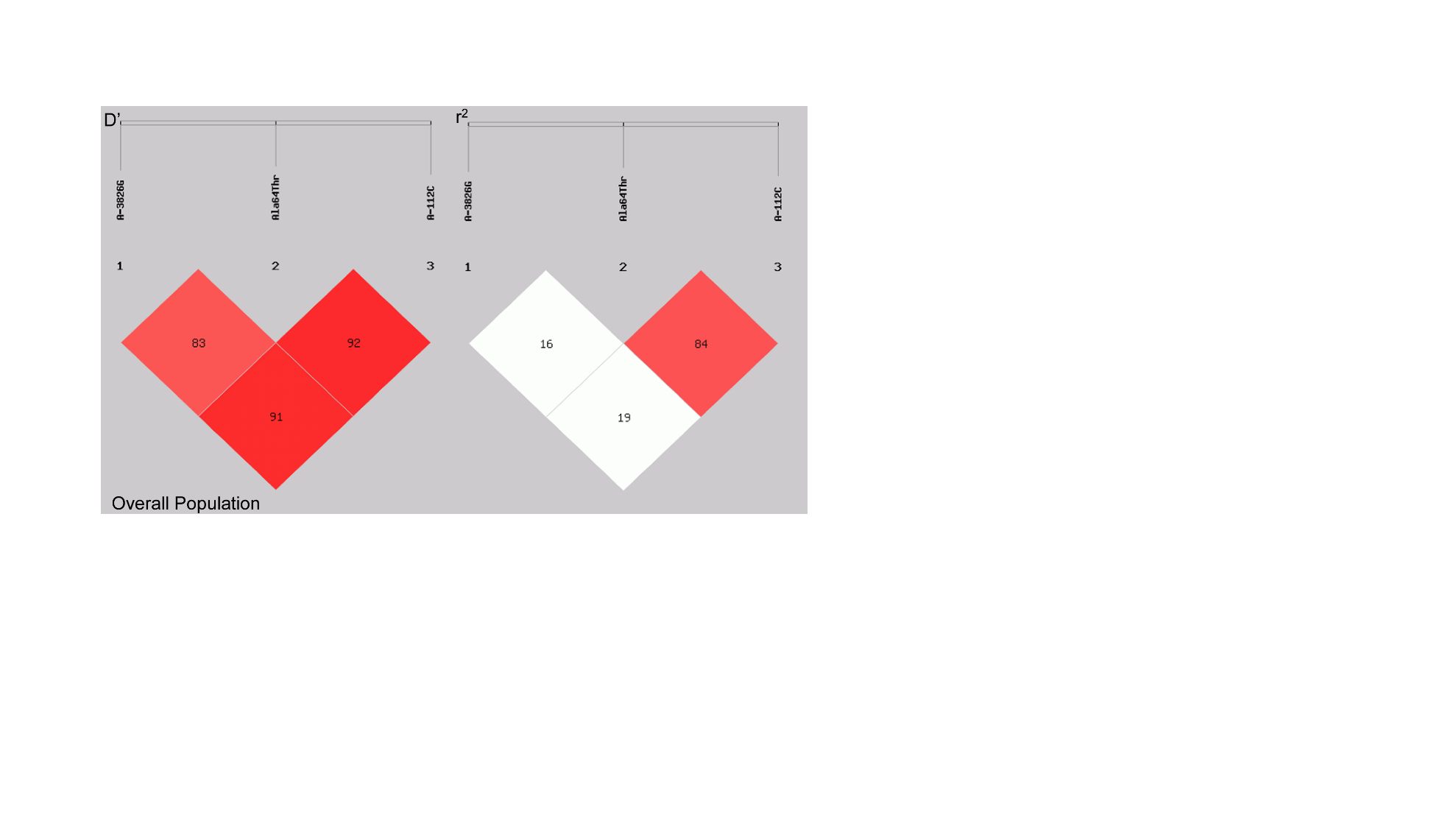

**Figure S2**: Linkage Disequilibrium heat maps per country

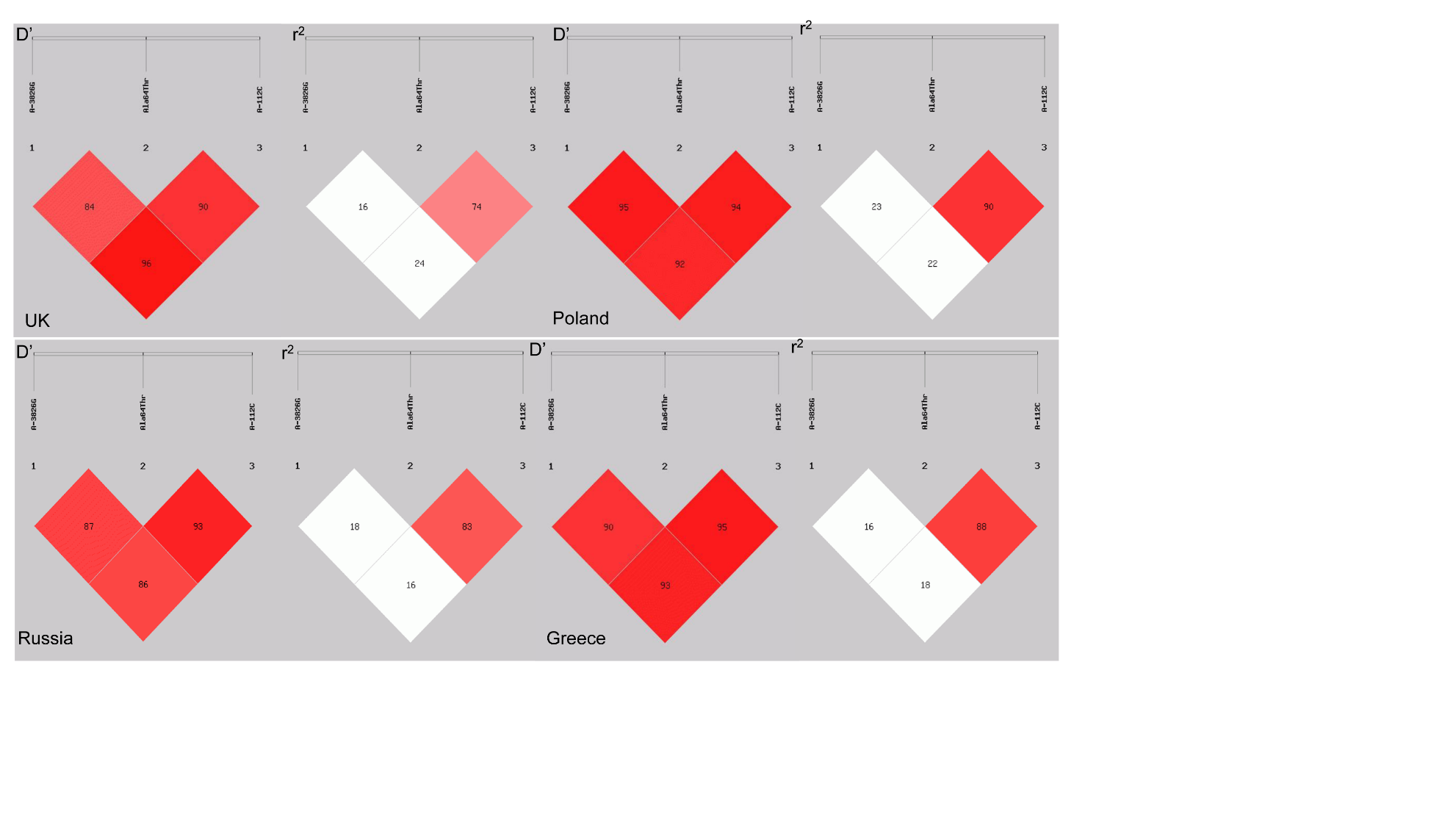

| Table S1: Haplotype frequencies for UK, Greece, Poland and Russia as well as for the overall study population for A-3826G, Ala64Thr and A-112C. | | | | | | | | | | |
| --- | --- | --- | --- | --- | --- | --- | --- | --- | --- | --- |
| Haplotype | **Overall** | | **UK** | | **Greece** | | **Poland** | | **Russia** | |
|  | CMP | Healthy | CMP | Healthy | CMP | Healthy | CMP | Healthy | CMP | Healthy |
| A G A | 1314.17 (0.712) | 1432.11 (0.699) | 125.76 (0.691) | 244.95 (0.684) | 343.10 (0.689) | 304.30 (0.673) | 376.65 (0.750) | 513.99 (0.706) | 435.92 (0.703) | 369.44  (0.724) |
|  | OR: 1.10 (CI95%: 0.96-1.27); x^2^=1.845, p=0.174 | | OR: 1.01 (CI95%: 0.68-1.49); x^2^=0.001, p=0.976 | | OR: 1.10 (CI95%: 0.83-1.45); x^2^=0.427, p=0.513 | | **OR: 1.33 (CI95%: 1.03-1.73); x^2^=4.617, p=0.032** | | OR: 0,96 (CI95%: 0.74-1.25); x^2^=0.09, p=0.764 | |
| G A C | 119.84 (0.065) | 174.03 (0.085) | 12.76 (0.070) | 30.92 (0.086) | 28.60 (0.057) | 44.75 (0.099) | 37.65 (0.075) | 63.00 (0.087) | 40.88 (0.066) | 35.44  (0.069) |
|  | **OR: 0.76 (CI95%: 0.60-0.96); x^2^=5.206, p=0.023** | | OR: 0.79 (CI95%: 0.40-1.56); x^2^=0.463, p=0.496 | | **OR: 0.56 (CI95%: 0.34-0.91); x^2^=5.635, p=0.017** | | OR: 0.87 (CI95%: 0.57-1.33); x^2^=0.424, p=0.52 | | OR: 0.94 (CI95%: 0.61-1.55); x^2^=0.023, p=0.879 | |
| G G A | 372.72 (0.202) | 417.85 (0.204) | 39.24 (0.216) | 70.97 (0.198) | 118.88 (0.239) | 98.70 (0.218) | 78.33 (0.156) | 148.00 (0.203) | 125.03  (0.202) | 99.55  (0.195) |
|  | OR: 0.999 (CI95%: 0.85-1.17); x^2^=0.001, p=0.991 | | OR: 1.10 (CI95%: 0.71-1.71); x^2^=0.184, p=0.668 | | OR: 1.13 (CI95%: 0.83-1.53); x^2^=0.630, p=0.428 | | **OR: 0.74 (CI95%: 0.55-0.99); x^2^=3.948, p=0.047** | | OR: 1.07 (CI95%: 0.79-1.43); x^2^=0.183, p=0.669 | |
| Note: Haplotypes with frequencies lower than 3 % were omitted.  Key: CMP = individuals with cardio-metabolic pathologies; OR = odds ratio; CI95% = 95% confidence interval. | | | | | | | | | | |

| Table S2: Haplotype frequencies in Armenia for the A-3826G and Ala64Thr SNPs. | | |
| --- | --- | --- |
| Haplotypes | **CMP** | **Healthy** |
| A A | 22.02 (0.052) | 2.60 (0.013) |
|  | **OR: 4.10 (CI95%: 1.12-14.98); x^2^ = 5.3, p = 0.021** | |
| A G | 278.98 (0.655) | 147.40 (0.744) |
|  | **OR: 0.65 (CI95%: 0.45-0.95); x^2^ = 5.011, p = 0.025** | |
| G A | 27.98 (0.066) | 10.40 (0.053) |
|  | OR: 1.27 (CI95%: 0.61-2.64); x^2^ = 0.405, p = 0.524 | |
| G G | 97.02 (0.228) | 37.60 (0.190) |
|  | OR: 1.26 (CI95%: 0.83-1.92); x^2^ = 1.144, p = 0.285 | |
| Key: CMP = individuals with cardio-metabolic pathologies; OR = odds ratio; CI95% = 95% confidence interval. | | |

**Table S3:** Prevalence rates for *UCP1* polymorphisms for the overall sample size and per country. SE: standard error; CMP: cardio-metabolic pathologies risk factors; UK: United Kingdom. Data for A-112C *UCP1* polymorphism are not available for Armenia.

| **Polymorphism** | **Group / Genotype** | **Events** | **Sample size** | **Prevalence** | **SE** |
| --- | --- | --- | --- | --- | --- |
| **A-1766G** | A-1766G overall AA | 2216 | 2283 | 0.97 | 0.02 |
|  | A-1766G overall AG | 40 | 2283 | 0.02 | 0.003 |
|  | A-1766G overall GG | 3 | 2283 | 0.001 | 0.001 |
|  | A-1766G healthy AA | 1116 | 1139 | 0.98 | 0.03 |
|  | A-1766G healthy AG | 17 | 1139 | 0.01 | 0.004 |
|  | A-1766G healthy GG | 2 | 1139 | 0.002 | 0.001 |
|  | A-1766G CMP AA | 1079 | 1144 | 0.94 | 0.03 |
|  | A-1766G CMP AG | 23 | 1144 | 0.02 | 0.004 |
|  | A-1766G CMP GG | 1 | 1144 | 0.001 | 0.001 |
|  | A-1766G UK overall AA | 264 | 273 | 0.97 | 0.06 |
|  | A-1766G UK overall AG | 6 | 273 | 0.02 | 0.01 |
|  | A-1766G UK overall GG | 2 | 273 | 0.01 | 0.01 |
|  | A-1766G UK healthy AA | 173 | 181 | 0.96 | 0.07 |
|  | A-1766G UK healthy AG | 6 | 181 | 0.03 | 0.01 |
|  | A-1766G UK healthy GG | 1 | 181 | 0.01 | 0.01 |
|  | A-1766G UK CMP AA | 91 | 92 | 0.99 | 0.10 |
|  | A-1766G UK CMP AG | 0 | 92 | 0.00 | 0 |
|  | A-1766G UK CMP GG | 1 | 92 | 0.01 | 0.01 |
|  | A-1766G Armenia overall AA | 289 | 331 | 0.87 | 0.05 |
|  | A-1766G Armenia overall AG | 13 | 331 | 0.04 | 0.01 |
|  | A-1766G Armenia overall GG | 0 | 331 | 0.00 | 0 |
|  | A-1766G Armenia healthy AA | 102 | 105 | 0.97 | 0.10 |
|  | A-1766G Armenia healthy AG | 3 | 105 | 0.03 | 0.02 |
|  | A-1766G Armenia healthy GG | 0 | 105 | 0.00 | 0 |
|  | A-1766G Armenia CMP AA | 187 | 226 | 0.83 | 0.06 |
|  | A-1766G Armenia CMP AG | 10 | 226 | 0.04 | 0.01 |
|  | A-1766G Armenia CMP GG | 0 | 226 | 0.00 | 0 |
|  | A-1766G Poland overall AA | 615 | 617 | 0.997 | 0.04 |
|  | A-1766G Poland overall AG | 2 | 617 | 0.003 | 0.002 |
|  | A-1766G Poland overall GG | 0 | 617 | 0.00 | 0 |
|  | A-1766G Poland healthy AA | 363 | 365 | 0.99 | 0.05 |
|  | A-1766G Poland healthy AG | 2 | 365 | 0.01 | 0.004 |
|  | A-1766G Poland healthy GG | 0 | 365 | 0.00 | 0 |
|  | A-1766G Poland CMP AA | 0 | 252 | 0.00 | 0 |
|  | A-1766G Poland CMP AG | 0 | 252 | 0.00 | 0 |
|  | A-1766G Poland CMP GG | 0 | 252 | 0.00 | 0 |
|  | A-1766G Russia overall AA | 559 | 565 | 0.99 | 0.04 |
|  | A-1766G Russia overall AG | 6 | 565 | 0.01 | 0.004 |
|  | A-1766G Russia overall GG | 0 | 565 | 0.00 | 0 |
|  | A-1766G Russia healthy AA | 254 | 255 | 0.996 | 0.06 |
|  | A-1766G Russia healthy AG | 1 | 255 | 0.004 | 0.004 |
|  | A-1766G Russia healthy GG | 0 | 255 | 0.00 | 0 |
|  | A-1766G Russia CMP AA | 305 | 310 | 0.98 | 0.06 |
|  | A-1766G Russia CMP AG | 5 | 310 | 0.02 | 0.01 |
|  | A-1766G Russia CMP GG | 0 | 310 | 0.00 | 0 |
|  | A-1766G Greece overall AA | 489 | 529 | 0.92 | 0.04 |
|  | A-1766G Greece overall AG | 13 | 529 | 0.02 | 0.01 |
|  | A-1766G Greece overall GG | 1 | 529 | 0.002 | 0.002 |
|  | A-1766G Greece healthy AA | 224 | 233 | 0.96 | 0.06 |
|  | A-1766G Greece healthy AG | 5 | 233 | 0.02 | 0.01 |
|  | A-1766G Greece healthy GG | 1 | 233 | 0.004 | 0.004 |
|  | A-1766G Greece CMP AA | 244 | 264 | 0.92 | 0.06 |
|  | A-1766G Greece CMP AG | 8 | 264 | 0.03 | 0.01 |
|  | A-1766G Greece CMP GG | 0 | 264 | 0.00 | 0 |
| **A-3826G** | A-3826G Overall AA | 1167 | 2283 | 0.51 | 0.01 |
|  | A-3826G Overall AG | 919 | 2283 | 0.40 | 0.01 |
|  | A-3826G Overall GG | 193 | 2283 | 0.08 | 0.01 |
|  | A-3826G Healthy AA | 571 | 1139 | 0.50 | 0.02 |
|  | A-3826G Healthy AG | 463 | 1139 | 0.41 | 0.02 |
|  | A-3826G Healthy GG | 97 | 1139 | 0.09 | 0.01 |
|  | A-3826G CMP AA | 584 | 1144 | 0.51 | 0.02 |
|  | A-3826G CMP AG | 448 | 1144 | 0.39 | 0.02 |
|  | A-3826G CMP GG | 95 | 1144 | 0.08 | 0.01 |
|  | A-3826G UK overall AA | 136 | 273 | 0.50 | 0.04 |
|  | A-3826G UK overall AG | 105 | 273 | 0.38 | 0.04 |
|  | A-3826G UK overall GG | 31 | 273 | 0.11 | 0.02 |
|  | A-3826G UK healthy AA | 88 | 181 | 0.49 | 0.05 |
|  | A-3826G UK healthy AG | 74 | 181 | 0.41 | 0.05 |
|  | A-3826G UK healthy homozygous GG | 18 | 181 | 0.10 | 0.02 |
|  | A-3826G UK CMP AA | 48 | 92 | 0.52 | 0.08 |
|  | A-3826G UK CMP AG | 31 | 92 | 0.34 | 0.06 |
|  | A-3826G UK CMP GG | 13 | 92 | 0.14 | 0.04 |
|  | A-3826G Armenia overall AA | 168 | 331 | 0.51 | 0.04 |
|  | A-3826G Armenia overall AG | 129 | 331 | 0.39 | 0.03 |
|  | A-3826G Armenia overall GG | 24 | 331 | 0.07 | 0.01 |
|  | A-3826G Armenia healthy AA | 57 | 105 | 0.54 | 0.07 |
|  | A-3826G Armenia healthy AG | 37 | 105 | 0.35 | 0.06 |
|  | A-3826G Armenia healthy GG | 6 | 105 | 0.06 | 0.02 |
|  | A-3826G Armenia CMP AA | 111 | 226 | 0.49 | 0.05 |
|  | A-3826G Armenia CMP AG | 92 | 226 | 0.41 | 0.04 |
|  | A-3826G Armenia CMP GG | 18 | 226 | 0.08 | 0.02 |
|  | A-3826G Poland overall AA | 323 | 617 | 0.52 | 0.03 |
|  | A-3826G Poland overall AG | 254 | 617 | 0.41 | 0.03 |
|  | A-3826G Poland overall GG | 40 | 617 | 0.06 | 0.01 |
|  | A-3826G Poland healthy AA | 179 | 365 | 0.49 | 0.04 |
|  | A-3826G Poland healthy AG | 159 | 365 | 0.44 | 0.03 |
|  | A-3826G Poland healthy GG | 27 | 365 | 0.07 | 0.01 |
|  | A-3826G Poland CMP AA | 144 | 252 | 0.57 | 0.05 |
|  | A-3826G Poland CMP AG | 95 | 252 | 0.38 | 0.04 |
|  | A-3826G Poland CMP GG | 13 | 252 | 0.05 | 0.01 |
|  | A-3826G Russia overall AA | 296 | 565 | 0.52 | 0.03 |
|  | A-3826G Russia overall AG | 222 | 565 | 0.39 | 0.03 |
|  | A-3826G Russia overall GG | 47 | 565 | 0.08 | 0.01 |
|  | A-3826G Russia healthy AA | 140 | 255 | 0.55 | 0.05 |
|  | A-3826G Russia healthy AG | 93 | 255 | 0.36 | 0.04 |
|  | A-3826G Russia healthy GG | 22 | 255 | 0.09 | 0.02 |
|  | A-3826G Russia CMP AA | 156 | 310 | 0.50 | 0.04 |
|  | A-3826G Russia CMP AG | 129 | 310 | 0.42 | 0.04 |
|  | A-3826G Russia CMP GG | 25 | 310 | 0.08 | 0.02 |
|  | A-3826G Greece overall AA | 244 | 529 | 0.46 | 0.03 |
|  | A-3826G Greece overall AG | 209 | 529 | 0.40 | 0.03 |
|  | A-3826G Greece overall GG | 51 | 529 | 0.10 | 0.01 |
|  | A-3826G Greece healthy AA | 107 | 233 | 0.46 | 0.04 |
|  | A-3826G Greece healthy AG | 100 | 233 | 0.43 | 0.04 |
|  | A-3826G Greece healthy GG | 24 | 233 | 0.10 | 0.02 |
|  | A-3826G Greece CMP AA | 125 | 264 | 0.47 | 0.04 |
|  | A-3826G Greece CMP AG | 101 | 264 | 0.38 | 0.04 |
|  | A-3826G Greece CMP GG | 26 | 264 | 0.10 | 0.02 |
| **Ala64Thr** | Ala64Thr Overall GG | 1893 | 2283 | 0.83 | 0.02 |
|  | Ala64Thr Overall GA | 363 | 2283 | 0.16 | 0.01 |
|  | Ala64Thr Overall AA | 19 | 2283 | 0.01 | 0.002 |
|  | Ala64Thr Healthy GG | 944 | 1139 | 0.83 | 0.03 |
|  | Ala64Thr Healthy GA | 175 | 1139 | 0.15 | 0.01 |
|  | Ala64Thr Healthy AA | 13 | 1139 | 0.01 | 0.003 |
|  | Ala64Thr CMP GG | 928 | 1144 | 0.81 | 0.03 |
|  | Ala64Thr CMP GA | 188 | 1144 | 0.16 | 0.01 |
|  | Ala64Thr CMP AA | 6 | 1144 | 0.01 | 0.002 |
|  | Ala64Thr UK overall GG | 225 | 273 | 0.82 | 0.05 |
|  | Ala64Thr UK overall GA | 43 | 273 | 0.16 | 0.02 |
|  | Ala64Thr UK overall AA | 4 | 273 | 0.01 | 0.01 |
|  | Ala64Thr UK healthy GG | 148 | 181 | 0.82 | 0.07 |
|  | Ala64Thr UK healthy GA | 28 | 181 | 0.15 | 0.03 |
|  | Ala64Thr UK healthy AA | 4 | 181 | 0.02 | 0.01 |
|  | Ala64Thr UK CMP GG | 77 | 92 | 0.84 | 0.10 |
|  | Ala64Thr UK CMP GA | 15 | 92 | 0.16 | 0.04 |
|  | Ala64Thr UK CMP AA | 0 | 92 | 0.00 | 0 |
|  | Ala64Thr Armenia overall GG | 254 | 331 | 0.77 | 0.05 |
|  | Ala64Thr Armenia overall GA | 67 | 331 | 0.20 | 0.02 |
|  | Ala64Thr Armenia overall AA | 0 | 331 | 0.00 | 0 |
|  | Ala64Thr Armenia healthy GG | 90 | 105 | 0.86 | 0.09 |
|  | Ala64Thr Armenia healthy GA | 14 | 105 | 0.13 | 0.04 |
|  | Ala64Thr Armenia healthy AA | 0 | 105 | 0.00 | 0 |
|  | Ala64Thr Armenia CMP GG | 164 | 226 | 0.73 | 0.06 |
|  | Ala64Thr Armenia CMP GA | 53 | 226 | 0.23 | 0.03 |
|  | Ala64Thr Armenia CMP AA | 0 | 226 | 0.00 | 0 |
|  | Ala64Thr Poland overall GG | 515 | 617 | 0.83 | 0.04 |
|  | Ala64Thr Poland overall GA | 96 | 617 | 0.16 | 0.02 |
|  | Ala64Thr Poland overall AA | 6 | 617 | 0.01 | 0.004 |
|  | Ala64Thr Poland healthy GG | 304 | 365 | 0.83 | 0.05 |
|  | Ala64Thr Poland healthy GA | 58 | 365 | 0.16 | 0.02 |
|  | Ala64Thr Poland healthy AA | 3 | 365 | 0.01 | 0.005 |
|  | Ala64Thr Poland CMP GG | 211 | 252 | 0.84 | 0.06 |
|  | Ala64Thr Poland CMP GA | 38 | 252 | 0.15 | 0.02 |
|  | Ala64Thr Poland CMP AA | 3 | 252 | 0.01 | 0.01 |
|  | Ala64Thr Russia overall GG | 475 | 565 | 0.84 | 0.04 |
|  | Ala64Thr Russia overall GA | 85 | 565 | 0.15 | 0.02 |
|  | Ala64Thr Russia overall AA | 5 | 565 | 0.01 | 0.004 |
|  | Ala64Thr Russia healthy GG | 218 | 255 | 0.85 | 0.06 |
|  | Ala64Thr Russia healthy GA | 34 | 255 | 0.13 | 0.02 |
|  | Ala64Thr Russia healthy AA | 3 | 255 | 0.01 | 0.01 |
|  | Ala64Thr Russia CMP GG | 257 | 310 | 0.83 | 0.05 |
|  | Ala64Thr Russia CMP GA | 51 | 310 | 0.16 | 0.02 |
|  | Ala64Thr Russia CMP AA | 2 | 310 | 0.01 | 0.005 |
|  | Ala64Thr Greece overall GG | 424 | 529 | 0.80 | 0.04 |
|  | Ala64Thr Greece overall GA | 72 | 529 | 0.14 | 0.02 |
|  | Ala64Thr Greece overall AA | 4 | 529 | 0.01 | 0.004 |
|  | Ala64Thr Greece healthy GG | 184 | 233 | 0.79 | 0.06 |
|  | Ala64Thr Greece healthy GA | 41 | 233 | 0.18 | 0.03 |
|  | Ala64Thr Greece healthy AA | 3 | 233 | 0.01 | 0.01 |
|  | Ala64Thr Greece CMP GG | 219 | 264 | 0.83 | 0.06 |
|  | Ala64Thr Greece CMP GA | 31 | 264 | 0.12 | 0.02 |
|  | Ala64Thr Greece CMP AA | 1 | 264 | 0.004 | 0.004 |
| **A-112C** | A-112C Overall AA | 1634 | 2283 | 0.72 | 0.02 |
|  | A-112C Overall AC | 300 | 2283 | 0.13 | 0.01 |
|  | A-112C Overall CC | 18 | 2283 | 0.01 | 0.002 |
|  | A-112C Healthy AA | 847 | 1139 | 0.74 | 0.03 |
|  | A-112C Healthy AC | 169 | 1139 | 0.15 | 0.01 |
|  | A-112C Healthy CC | 11 | 1139 | 0.01 | 0.003 |
|  | A-112C CMP AA | 766 | 1144 | 0.67 | 0.02 |
|  | A-112C CMP AC | 131 | 1144 | 0.11 | 0.01 |
|  | A-112C CMP CC | 7 | 1144 | 0.01 | 0.002 |
|  | A-112C UK overall AA | 218 | 273 | 0.80 | 0.05 |
|  | A-112C UK overall AC | 50 | 273 | 0.18 | 0.03 |
|  | A-112C UK overall CC | 2 | 273 | 0.01 | 0.01 |
|  | A-112C UK healthy AA | 144 | 181 | 0.80 | 0.07 |
|  | A-112C UK healthy AC | 33 | 181 | 0.18 | 0.03 |
|  | A-112C UK healthy CC | 2 | 181 | 0.01 | 0.01 |
|  | A-112C UK CMP AA | 74 | 92 | 0.80 | 0.09 |
|  | A-112C UK CMP AC | 17 | 92 | 0.18 | 0.04 |
|  | A-112C UK CMP CC | 0 | 92 | 0.00 | 0 |
|  | A-112C Poland overall AA | 515 | 617 | 0.83 | 0.04 |
|  | A-112C Poland overall AC | 94 | 617 | 0.15 | 0.02 |
|  | A-112C Poland overall CC | 7 | 617 | 0.01 | 0.004 |
|  | A-112C Poland healthy AA | 303 | 365 | 0.83 | 0.05 |
|  | A-112C Poland healthy AC | 57 | 365 | 0.16 | 0.02 |
|  | A-112C Poland healthy CC | 4 | 365 | 0.01 | 0.01 |
|  | A-112C Poland CMP AA | 212 | 252 | 0.84 | 0.06 |
|  | A-112C Poland CMP AC | 37 | 252 | 0.15 | 0.02 |
|  | A-112C Poland CMP CC | 3 | 252 | 0.01 | 0.01 |
|  | A-112C Russia overall AA | 480 | 565 | 0.85 | 0.04 |
|  | A-112C Russia overall AC | 80 | 565 | 0.14 | 0.02 |
|  | A-112C Russia overall CC | 5 | 565 | 0.01 | 0.004 |
|  | A-112C Russia healthy AA | 218 | 255 | 0.85 | 0.06 |
|  | A-112C Russia healthy AC | 34 | 255 | 0.13 | 0.02 |
|  | A-112C Russia healthy CC | 3 | 255 | 0.01 | 0.01 |
|  | A-112C Russia CMP AA | 262 | 310 | 0.85 | 0.05 |
|  | A-112C Russia CMP AC | 46 | 310 | 0.15 | 0.02 |
|  | A-112C Russia CMP CC | 2 | 310 | 0.01 | 0.005 |
|  | A-112C Greece overall AA | 421 | 529 | 0.80 | 0.04 |
|  | A-112C Greece overall AC | 72 | 529 | 0.14 | 0.02 |
|  | A-112C Greece overall CC | 4 | 529 | 0.01 | 0.004 |
|  | A-112C Greece healthy AA | 182 | 233 | 0.78 | 0.06 |
|  | A-112C Greece healthy AC | 45 | 233 | 0.19 | 0.03 |
|  | A-112C Greece healthy CC | 2 | 233 | 0.01 | 0.01 |
|  | A-112C Greece CMP AA | 218 | 264 | 0.83 | 0.06 |
|  | A-112C Greece CMP AC | 31 | 264 | 0.12 | 0.02 |
|  | A-112C Greece CMP CC | 2 | 264 | 0.01 | 0.01 |

**Table S4**: Allele frequencies in the overall study population

| SNP | Allele | CMP individuals | | Healthy | | OR | %95 CI | p |
| --- | --- | --- | --- | --- | --- | --- | --- | --- |
|  |  | **no** | **freq.** | **no** | **freq.** |  |  |  |
| A-1766G | A  G | 2181  25 | 0.99  0.01 | 2249  21 | 0.99  0.01 | 1.23 | 0.69-2.20 | p=0.49 |
| A-3826G | A  G | 1616  638 | 0.72  0.28 | 1605  657 | 0.71  0.29 | 0.96 | 0.85-1.10 | p=0.58 |
| Ala64Thr | G  A | 2044  200 | 0.91  0.09 | 2063  201 | 0.91  0.09 | 1.00 | 0.82-1.23 | p=0.97 |
| A-112C | A  C | 1663  145 | 0.92  0.08 | 1863  191 | 0.91  0.09 | 0.85 | 0.68 -1.07 | p=0.16 |

**Table S5:** Allele frequencies in the Armenian population

| SNP | Allele | CMP individuals | | Healthy | | OR | %95 CI | p |
| --- | --- | --- | --- | --- | --- | --- | --- | --- |
|  |  | **no** | **freq.** | **no** | **freq.** |  |  |  |
| A-1766G | A  G | 384  10 | 0.97  0.03 | 207  3 | 0.99  0.01 | 1.80 | 0.49-6.60 | p=0.37 |
| A-3826G | A  G | 314  128 | 0.71  0.29 | 151  49 | 0.75  0.25 | 1.26 | 0.86-1.84 | p=0.24 |
| Ala64Thr | G  A | 381  53 | 0.88  0.12 | 294  114 | 0.72  0.28 | 0.36 | 0.25- 0.51 | p<0.001 |
| A-112C | A  C | -  - | -  - | -  - | -  - | - | - |  |

**Table S6**: Allele frequencies in the Greek population

| SNP | Allele | CMP individuals | | Healthy | | OR | %95 CI | p |
| --- | --- | --- | --- | --- | --- | --- | --- | --- |
|  |  | **no** | **freq.** | **no** | **freq.** |  |  |  |
| A-1766G | A  G | 496  8 | 0.99  0.01 | 453  5 | 0.99  0.01 | 1.46 | 0.47 - 4.50 | p=0.51 |
| A-3826G | A  G | 351  153 | 0.70  0.30 | 314  148 | 0.68  0.32 | 0.92 | 0.70 - 1.21 | p=0.57 |
| Ala64Thr | G  A | 469  33 | 0.93  0.07 | 409  47 | 0.9  0.1 | 0.61 | 0.38 - 0.97 | p=0.04 |
| A-112C | A  C | 467  35 | 0.93  0.07 | 409  49 | 0.89  0.11 | 0.63 | 0.40 - 0.98 | p=0.04 |

**Table S7:** Allele frequencies in the Polish population

| SNP | Allele | CMP individuals | | Healthy | | OR | %95 CI | p |
| --- | --- | --- | --- | --- | --- | --- | --- | --- |
|  |  | **no** | **freq.** | **no** | **freq.** |  |  |  |
| A-1766G | A  G | 504  0 | 100  0.0 | 728  2 | 0.99  0.01 | - | - |  |
| A-3826G | A  G | 383  121 | 0.76  0.24 | 517  213 | 0.71  0.29 | 0.77 | 0.59 - 0.99 | p=0.045 |
| Ala64Thr | G  A | 460  44 | 0.91  0.09 | 666  66 | 0.91  0.09 | 0.97 | 0.65 - 1.44 | p=0.86 |
| A-112C | A  C | 461  43 | 0.91  0.01 | 663  65 | 0.91  0.09 | 0.95 | 0.64 - 1.42 | p=0.81 |

**Table S8:** Allele frequencies in the UK population

| SNP | Allele | CMP individuals | | Healthy | | OR | %95 CI | p |
| --- | --- | --- | --- | --- | --- | --- | --- | --- |
|  |  | **no** | **freq.** | **no** | **freq.** |  |  |  |
| A-1766G | A  G | 182  2 | 0.99  0.01 | 352  8 | 0.98  0.02 | 0.48 | 0.10 - 2.30 | p=0.35 |
| A-3826G | A  G | 127  57 | 0.69  0.31 | 250  110 | 0.69  0.31 | 1.02 | 0.60 - 1.50 | p=0.92 |
| Ala64Thr | G  A | 169  15 | 0.92  0.08 | 324  36 | 0.9  0.1 | 0.80 | 0.43 - 1.50 | p=0.48 |
| A-112C | A  C | 165  17 | 0.91  0.09 | 321  37 | 0.90  0.10 | 0.89 | 0.49- 1.64 | p=0.72 |

**Table S9:** Allele frequencies in the Russian population

| SNP | Allele | CMP individuals | | Healthy | | OR | %95 CI | p |
| --- | --- | --- | --- | --- | --- | --- | --- | --- |
|  |  | **no** | **freq.** | **no** | **freq.** |  |  |  |
| A-1766G | A  G | 615  5 | 0.99  0.01 | 509  1 | 0.99  0.1 | 4.14 | 0.48 - 35.54 | p=0.16 |
| A-3826G | A  G | 441  179 | 0.71  0.29 | 373  137 | 0.73  0.27 | 1.11 | 0.85 - 1.44 | p=0.45 |
| Ala64Thr | G  A | 565  55 | 0.91  0.09 | 470  40 | 0.92  0.08 | 1.14 | 0.75 - 1.75 | p=0.54 |
| A-112C | A  C | 570  50 | 0.92  0.08 | 470  40 | 0.92  0.08 | 1.03 | 0.67 - 1.59 | p=0.89 |

| **Table 10.** Frequency of genotypes for A-3826G in CMP and healthy individuals. \| \| \| \| \| \| \|; \| \| Healthy \| \| CMP \| \| OR (95% CI) \| F-test; \| \| (n) \| (%) \| (n) \| (%) \| \|; Total sample \| AA \| 571 \| 50.49 \| 584 \| 51.82 \| \| 5.29 p=0.153; \| AG \| 463 \| 40.94 \| 448 \| 39.75 \| 0.95 (0.80-1.13) \|; \| GG \| 97 \| 8.57 \| 95 \| 8.43 \| 0.96 (0.71-1.30) \|; \| HWE \| 0.490 \| \| 0.819 \| \| \|; Armenia \| AA \| 57 \| 57.00 \| 111 \| 50.23 \| \| 3.13 p=0.354; \| AG \| 37 \| 37.00 \| 92 \| 41.63 \| 1.27 (0.78-2.09) \|; \| GG \| 6 \| 6.00 \| 18 \| 8.14 \| 1.47 (0.57-3.79) \|; \| HWE \| 0.998 \| \| 0.861 \| \| \|; Greece \| AA \| 107 \| 46.32 \| 125 \| 49.60 \| \| 0.58 p=0.747; \| AG \| 100 \| 43.29 \| 101 \| 40.08 \| 0.87 (0.59-1.26) \|; \| GG \| 24 \| 10.39 \| 26 \| 10.32 \| 0.93 (0.51-1.70) \|; \| HWE \| 0.929 \| \| 0.408 \| \| \|; Poland \| AA \| 179 \| 49.04 \| 144 \| 57.14 \| \| 4.21 p=0.120; \| AG \| 159 \| 43.56 \| 95 \| 37.70 \| 0.74 (0.53-1.04) \|; \| GG \| 27 \| 7.40 \| 13 \| 5.16 \| 0.61 (0.31-1.21) \|; \| HWE \| 0.302 \| \| 0.599 \| \| \|; Russia \| AA \| 140 \| 54.90 \| 156 \| 50.32 \| \| 1.57 p=0.463; \| AG \| 93 \| 36.47 \| 129 \| 41.61 \| 1.24 (0.88-1.76) \|; \| GG \| 22 \| 8.63 \| 25 \| 8.07 \| 1.02 (0.55-1.87) \|; \| HWE \| 0.251 \| \| 0.816 \| \| \|; UK \| AA \| 88 \| 48.89 \| 48 \| 52.17 \| \| 1.96 p=0.396; \| AG \| 74 \| 41.11 \| 31 \| 33.70 \| 0.77 (0.45-1.33) \|; \| GG \| 18 \| 10.00 \| 13 \| 14.13 \| 1.33 (0.61-2.92) \|; \| HWE \| 0.675 \| \| 0.042 \| \| \|; Key: CMP = cardio-metabolic pathologies; OR = odds ratio; HWE = p value for the Hardy-Weinberg equilibrium. \| \| \| \| \| \| \| **Table 11.** Frequency of genotypes for A-112C in CMP and healthy individuals. \| \| \| \| \| \| \|; \| \| Healthy \| \| CMP \| \| OR (95% CI) \| F-test; \| \| (n) \| (%) \| (n) \| (%) \| \|; Total sample \| AA \| 847 \| 82.47 \| 766 \| 84.74 \| \| 1.93 p=0.367; \| AC \| 169 \| 16.46 \| 131 \| 14.49 \| 0.86 (0.67-1.10) \|; \| CC \| 11 \| 1.07 \| 7 \| 0.77 \| 0.72 (0.29-1.82) \|; \| HWE \| 0.433 \| \| 0.593 \| \| \|; Greece \| AA \| 182 \| 79.48 \| 218 \| 86.85 \| \| 4.92 p=0.73; \| AC \| 45 \| 19.65 \| 31 \| 12.35 \| 0.58 (0.35-0.95) \|; \| CC \| 2 \| 0.87 \| 2 \| 0.80 \| 0.58 (0.35-0.95) \|; \| HWE \| 0.668 \| \| 0.448 \| \| \|; Poland \| AA \| 303 \| 83.24 \| 212 \| 84.13 \| \| 0.20 p=0.947; \| AC \| 57 \| 15.66 \| 37 \| 14.68 \| 0.93 (0.60-1.46) \|; \| CC \| 4 \| 1.10 \| 3 \| 1.19 \| 1.11 (0.27-4.54) \|; \| HWE \| 0.479 \| \| 0.347 \| \| \|; Russia \| AA \| 218 \| 85.49 \| 262 \| 84.52 \| \| 0.75 p=0.717; \| AC \| 34 \| 13.33 \| 46 \| 14.84 \| 1.12 (0.70-1.81) \|; \| CC \| 3 \| 1.18 \| 2 \| 0.65 \| 0.59 (0.12-3.04) \|; \| HWE \| 0.215 \| \| 0.990 \| \| \|; UK \| AA \| 144 \| 81.32 \| 74 \| 81.32 \| \| 0.64 p=0.941; \| AC \| 33 \| 18.44 \| 17 \| 18.68 \| 1.01 (0.53-1.93) \|; \| CC \| 2 \| 1.12 \| 0 \| 0.00 \| 0.39 (0.02-8.18) \|; \| HWE \| 0.943 \| \| 0.326 \| \| \|; Key: CMP = cardio-metabolic pathologies; OR = odds ratio; HWE = p value for the Hardy-Weinberg equilibrium. \| \| \| \| \| \| \| **Table S12.** Body mass index, waist-to-hip ratio and body fat percent [median (Q1,Q3)] across the different genotypes of *UCP1* SNPs across healthy controls and individuals with CMP in Poland. | | | | | | | | |
| --- | --- | --- | --- | --- | --- | --- | --- | --- |
|  |  | | BMI | | WHR | | Body fat % | |
| SNP | Genotype | | Healthy | CMP | Healthy | CMP | Healthy | CMP |
| A-3826G | | AA | 23.7 (21.7,25.6) | 31.3 (29.1,33.8) | 0.84 (0.79,0.90) | 0.96 (0.87,1.05) | 22.6 (18.3,28.6) | - |
|  |  | AG | 23.9 (22.5,25.6) | 31.2 (29.9,33.9) | 0.86 (0.81,0.90)^1^ | 0.95 (0.87,1.04) | 23.8 (18.0,27.6) | - |
|  |  | GG | 24.6 (22.3,26.7) | 30.1 (28.5,32.2) | 0.86 (0.77,0.90) | 0.95 (0.84,1.06) | 24.8 (22.4,28.1) | - |
| A-112C | | AA | 23.7 (22.0,25.6) | 31.2 (29.4,33.8) | 0.85 (0.79,0.90) | 0.96 (0.88,1.06) | 22.9 (18.3,28.3) | - |
|  |  | AC | 23.9 (22.1,25.9) | 31.3 (29.5,34.0) | 0.85 (0.79,0.90) | 0.94 (0.85,1.03) | 24.5 (19.4,28.4) | - |
|  |  | CC | 26.4 (25.5,27.3)^,2^ | 27.3 (27.3, 29.8) | 0.86 (0.84, 0.88) | 0.84 (0.76,0.92) | 22.7 (21.9,23.0) | - |
| Ala64Thr | | GG | 23.7 (22.0,25.6) | 31.3 (29.4,33.8) | 0.85 (0.78,0.89) | 0.96 (0.88, 1.06) | 22.9 (18.3,28.2) | - |
|  |  | GA | 24.0 (22.2,25.9)^1^ | 30.9 (29.0,33.8) | 0.85 (0.79,0.90) | 0.95 (0.85,1.03) | 24.3 (19.1,28.4) | - |
|  |  | AA | 27.5 (27.3,27.6) | 27.3 (27.3,29.8) | 0.83 (0.82,0.84) | 0.84 (0.76,0.92) | 22.1 (21.6,22.7) | - |
| Note: 1 = difference from AA significant at p≤0.05; 2 = difference from AC significant at p≤0.05;  Key: CMP = cardio-metabolic pathologies, BMI= body mass index, WHR= waist-to-hip ratio, Q=quartile. | | | | | | | | |

| **Table S13.** Body mass index, waist-to-hip ratio and body fat percent [median (Q1,Q3)] across the different genotypes of *UCP1* SNPs across healthy controls and individuals with CMP in Russia. | | | | | | | | |
| --- | --- | --- | --- | --- | --- | --- | --- | --- |
|  |  | | BMI | | WHR |  | Body fat % |  |
| SNP | Genotype | | Healthy | CMP | Healthy | CMP | Healthy | CMP |
| A-3826G | | AA | 25.9 (25.4,26.3) | 28.4 (26.1,34.1) | - | - | 29.0 (26.8,32.0) | 49.0 (45.8,50.6) |
|  |  | AG | 25.6 (25.2,26.2) | 29.5 (26.0,35.5) | - | - | 29.0 (27.0,32.0) | 48.3 (44.3,50.3) |
|  |  | GG | 25.8 (25.4,26.4) | 30.2 (26.9,38.9) | - | - | 28.0 (26.0,32.0) | 49.9 (46.8,51.0) |
| A-112C | | AA | 25.8 (25.2,26.3) | 28.9 (26.1,34.6) | - | - | 29.0 (27.0,32.0) | 48.6 (45.5,51.0) |
|  |  | AC | 26.0 (25.4,26.5) | 30.3 (26.4,36.0) | - | - | 30.0 (26.0,33.0) | 48.6 (45.6,50.1) |
|  |  | CC | 25.9 (25.5,26.1) | 29.8 (28.0,31.5) | - | - | 31.0 (29.0,33.0) | 50.4 (50.1, 50.5) |
| Ala64Thr | | GG | 25.8 (25.2,26.3) | 28.9 (26.0,34.7) | - | - | 29.0 (27.0,32.0) | 48.6 (44.2,51.0) |
|  |  | GA | 26.0 (25.5,26.5) | 29.9 (26.1,35.9) | - | - | 29.5 (26.0,33.0) | 48.4 (45.9,50.1) |
|  |  | AA | 25.9 (25.5,26.1) | 29.8 (28.0,31.5) | - | - | 31.0 (29.0,33.0) | 50.4 (50.2,50.5) |
| Key: CMP = cardio-metabolic pathologies, BMI= body mass index, WHR= waist-to-hip ratio, Q=quartile. | | | | | | | | |

| **Table S14.** Body mass index, waist-to-hip ratio and body fat percent [median (Q1,Q3)] across the different genotypes of UCP1 SNPs across healthy controls and individuals with CMP in Greece. | | | | | | | | |
| --- | --- | --- | --- | --- | --- | --- | --- | --- |
|  |  | | BMI | | WHR | | Body fat % | |
| SNP | Genotype | | Healthy | CMP | Healthy | CMP | Healthy | CMP |
| A-3826G | | AA | 26.4 (24.2,29.5) | 31.7 (28.9,34.4) | 0.94 (0.88,1.00) | 1.02 (0.96,1.05) | 29.4 (17.3,36.6) | 40.4 (34.3,43.4) |
|  |  | AG | 27.2 (23.8,29.2) | 31.7 (29.2,33.9) | 0.94 (0.88,1.02) | 1.02 (0.97,1.04) | 29.6 (19.8,36.6) | 39.0 (34.5,42.9) |
|  |  | GG | 28.7 (26.2,30.7) | 30.9 (28.3,33.5) | 0.94 (0.88,1.02) | 1.02 (0.97,1.06) | 26.5 (19.5,40.5) | 37.9 (36.6,44.0) |
| A-112C | | AA | 26.5 (23.9,29.2) | 31.6 (28.8,34.3) | 0.94 (0.88,1.00) | 1.03 (0.96,1.05) | 28.9 (17.5,36.4) | 39.1 (34.5,43.3) |
|  |  | AC | 28.8 (26.0,31.4)^1^ | 32.3 (29.3,33.6) | 0.95 (0.89,1.01) | 1.00 (0.96,1.03) | 31.4 (25.1,41.7)^1^ | 39.1 (33.5,44.5) |
|  |  | CC | 26.3 | 30.4 (29.1,31.6) | 1.03 | 1.00 (0.97,1.03) | 19.6 | 35.7 (30.1, 41.2) |
| Ala64Thr | | GG | 26.5 (23.9,29.1) | 31.5 (28.8,34.4) | 0.94 (0.88,1.00) | 1.03 (0.96,1.05) | 28.9 (17.6,36.3) | 39.1 (34.5,43.4) |
|  |  | GA | 29.0 (26.0,31.8)^2^ | 32.0 (28.9,33.6) | 0.95 (0.90,1.02) | 1.00 (0.96,1.04) | 34.5 (25.5,41.9)^2^ | 39.1 (31.4,44.5) |
|  |  | AA | 26.3 | 32.8 | 1.00 | 1.05 | 19.6 | 46.7 |
| Note: 1 = difference from AA significant at p≤0.05; 2 = difference from GG significant at p≤0.05; Q1 and Q3 values are reported only where more than one case was detected for a specific genotype.  Key: CMP = cardio-metabolic pathologies, BMI= body mass index, WHR= waist-to-hip ratio, Q=quartile. | | | | | | | | |

| **Table S15.** Body mass index, waist-to-hip ratio and body fat percent [median (Q1, Q3)] across the different genotypes of UCP1 SNPs across healthy controls and individuals with CMP in UK. | | | | | | | | |
| --- | --- | --- | --- | --- | --- | --- | --- | --- |
|  |  | | BMI | | WHR | | Body fat % | |
| SNP | Genotype | | Healthy | CMP | Healthy | CMP | Healthy | CMP |
| A-3826G | | AA | 25.6 (23.0,29.5) | 29.2 (26.9,34.5) | 0.86 (0.81,0.91) | 0.95 (0.92,0.97) | 29.6 (24.1,35.1) | 33.3 (27.4,38.4) |
|  |  | AG | 25.8 (23.3,29.4) | 30.1 (25.7,34.8) | 0.83 (0.78,0.90) | 0.94 (0.92,0.98) | 29.5(24.5,38.0) | 33.0 (28.6,38.4) |
|  |  | GG | 27.7 (24.1,31.1) | 29.8 (25.9,31.4) | 0.84 (0.80,0.89) | 0.94 (0.83,1.00) | 35.4 (28.2, 40.2) | 29.0 (26.9,37.0) |
| A-112C | | AA | 25.7 (23.1,29.6) | 29.2 (26.5,34.6) | 0.85 (0.79,0.91) | 0.95 (0.72,0.97) | 30.1 (25.1,37.1) | 33.0 (27.7,38.2) |
|  |  | AC | 26.0 (23.5,31.1)^1^ | 30.1 (25.3,33.4) | 0.84 (0.80,0.89) | 0.93 (0.84,0.97) | 29.8 (23.0,39.5)^1^ | 32.1 (26.9,39.5) |
|  |  | CC | 28.5 (26.6,30.5) | - | 0.88 (0.88,0.89) | - | 27.8 (19.8,35.8) | - |
| Ala64Thr | | GG | 25.7 (22.6,29.6) | 29.0 (25.9,34.5) | 0.85 (0.79,0.91) | 0.95 (0.92,0.97) | 30.1 (24.8,37.2) | 33.7 (27.1,38.1) |
|  |  | GA | 26.4 (23.7,30.6)^2^ | 31.4 (28.4,33.6) | 0.83 (0.79,0.90) | 0.93 (0.87, 0.97) | 30.0 (22.6,38.4)^2^ | 35.7 (28.2,40.3) |
|  |  | AA | 28.6 (24.7,35.3) | - | 0.88 (0.86,0.89) | - | 39.4 (29.1,46.3) | - |
| Key: CMP = cardio-metabolic pathologies, BMI= body mass index, WHR= waist-to-hip ratio, Q=quartile. | | | | | | | | |

**2. SYSTEMATIC REVIEW**

**2.1. Materials and Methods**

**2.1.1. Search strategy, selection criteria and meta-analysis process**

The searching procedure, screening of the titles, abstracts and full texts for eligibility as well as the selection of the included studies were conducted independently by two investigators (PS and AEP) and any conflicts were resolved through discussion with a third investigator (PCD). We excluded reviews, conference proceedings and magazine articles and we also searched the reference lists of the included studies to identify potential eligible publications.

Two independent investigators (PS and AEP) evaluated the risk of bias (ROB) of the included studies in the systematic review, via the 13-item of Research Triangle Institute item bank,[1] which is designed for observational studies and has previously shown median interrater agreement of 75%[2] and 93.5%.[3] Conflicts in the risk of bias assessment were resolved by an independent referee investigator (PCD). Data extraction was performed independently by two investigators (PS and AW) and conflicts were resolved through consensus and supervision by a third researcher (PCD). For all studies, we extracted the first author’s name, year of publication, methodological design, genotyping method, participants’ characteristics and main outcomes (i.e. means±standard deviations/standard error, percentages, confidence intervals, frequencies, etc.).

We conducted prevalence meta-analyses by dividing the incidence of CMP by the overall sample size [a (incidence of genotype)/b (sample size)] of each study for *UCP1* A-3826G, A-1766G, Ala64Thr and A-112C SNPs. These meta-analyses were conducted for each one of the *UCP1* homozygous and heterozygous genotypes as well as for the mutant alleles of each studied SNP. Standard errors for these meta-analyses were calculated using the following formula: a (incidence of genotype/allele) /[a (incidence of genotype/allele) *b (sample size)]^2^. Standard errors were then used for weighted proportions and the RevMan 5.3 software[4] to generate forest and funnel plots. We also conducted odds ratio meta-analyses, using a dichotomous, inverse variance, random-effect model, via the RevMan 5.3 software. Incidence of each one of the *UCP1* homozygous and heterozygous genotypes and mutant alleles were calculated between a group of CMP individuals and a group of healthy participants, while weighted proportions were calculated based on each study’s sample size. For all meta-analyses, we evaluated the 95% confidence interval (CI) and heterogeneity between studies using the I² statistic. We considered a statistically significant result for heterogeneity when p<0.10, while interpretation of I^2^ index was made based on previous guidelines.[5] Where pertinent, standard error (SE) was converted to standard deviation (SD) using the following formula: SD= SE*√n.[5]

*2.1.1.a. Searching algorithm used in PubMed*

(((UCP1 variant*[Title/Abstract]) OR (UCP-1 variant*[Title/Abstract]) OR (uncoupling protein-1 variant*[Title/Abstract]) OR (uncoupling protein 1 variant*[Title/Abstract]) OR (thermogenin variant*[Title/Abstract]) OR (UCP-1 polymorphism*[Title/Abstract]) OR (UCP1 polymorphism*[Title/Abstract]) OR (uncoupling protein-1 polymorphism*[Title/Abstract]) OR (uncoupling protein 1 polymorphism*[Title/Abstract]) OR (thermogenin polymorphism*[Title/Abstract]) OR (UCP-1 gen*[Title/Abstract]) OR (UCP1 gen*[Title/Abstract]) OR (uncoupling protein-1 gen*[Title/Abstract]) OR (uncoupling protein 1 gen*[Title/Abstract]) OR (thermogenin gen*[Title/Abstract]) OR (UCP-1 single nucleotide polymorphism*[Title/Abstract]) OR (UCP1 single nucleotide polymorphism*[Title/Abstract]) OR (uncoupling protein-1 single nucleotide polymorphism*[Title/Abstract]) OR (uncoupling protein 1 single nucleotide polymorphism*[Title/Abstract]) OR (thermogenin single nucleotide polymorphism*[Title/Abstract]) OR (UCP-1 SNP*[Title/Abstract]) OR (UCP1 SNP*[Title/Abstract]) OR (uncoupling protein-1 SNP*[Title/Abstract]) OR (uncoupling protein 1 SNP*[Title/Abstract]) OR (thermogenin SNP*[Title/Abstract]) OR (UCP-1 mut*[Title/Abstract]) OR (UCP1 mut*[Title/Abstract]) OR (uncoupling protein-1 mut*[Title/Abstract]) OR (uncoupling protein 1 mut*[Title/Abstract]) OR (thermogenin mut*[Title/Abstract]) OR (UCP1 haplotype*[Title/Abstract]) OR (UCP-1 haplotype*[Title/Abstract]) OR (A-3826G[Title/Abstract]) OR (A-112C[Title/Abstract]) OR (Ala64Thr[Title/Abstract]) OR (A-3826G[Title/Abstract]) OR (A-1766G[Title/Abstract]) OR (A-112C[Title/Abstract]) OR (+1068G/A[Title/Abstract]) OR (rs10011540[Title/Abstract]) OR (rs45539933[Title/Abstract]) OR (rs3811791[Title/Abstract]) OR (rs1800592[Title/Abstract]) OR (-3826A>G[Title/Abstract]) OR (-112A>C[Title/Abstract]) OR (A-1766G[Title/Abstract]) OR (-1766A>G[Title/Abstract]) OR (uncoupling protein 1[Title/Abstract]) OR (uncoupling protein one[Title/Abstract])) AND ((metabolic syndrome[Title/Abstract]) OR (metabolic dis*[Title/Abstract]) OR (cardiometabolic dis*[Title/Abstract]) OR (CMD*[Title/Abstract]) OR (cardiometabolic disease[MeSH Terms]) OR (obesity[Title/Abstract]) OR (diabetes[Title/Abstract]) OR (T2DM[Title/Abstract]) OR (T2D[Title/Abstract]) OR (type 2 diabetes[Title/Abstract]) OR (type 2 diabetes mellitus[Title/Abstract]) OR (type II diabetes mellitus) OR (cardiovascular dis*[Title/Abstract]) OR (CVD*[Title/Abstract]) OR (cardiovascular disease[MeSH Terms]))) NOT ((animals[MeSH Terms]) NOT (humans[MeSH Terms]))

**Figure S3:** PRISMA flowchart

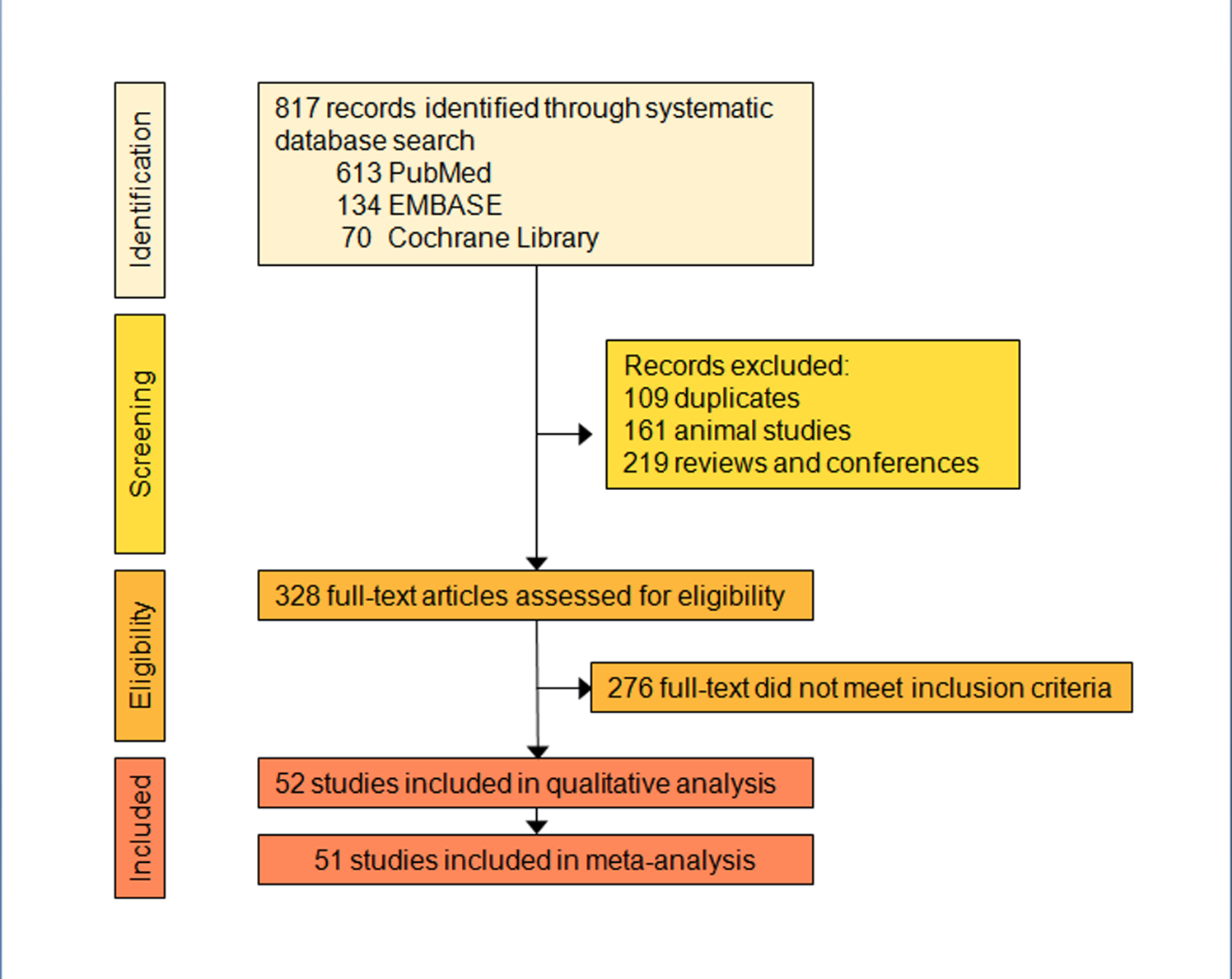

**2.2. Results (Tables and Figures)**

The risk of bias assessment revealed that 58.97% of the studies displayed low selection bias, 2.54% displayed unclear ROB and 38.49% not applicable ROB. In performance and selective bias all studies displayed low ROB, while in detection bias 2.54% of the studies showed high ROB and 97.44% unclear ROB. In attrition bias, 2.54% of the studies displayed low ROB and 97.44% not applicable ROB. Finally, in confounding bias 89.75% of the studies displayed low ROB, 2.54% displayed high ROB and 7.69% unclear ROB.

**Table S16:** Risk of bias assessment results for the included studies in the systematic review.

| **First author** | **Selection bias** | **Performance bias** | **Detection bias** | **Attrition bias** | **Selective outcome reporting** | **Confounding** |
| --- | --- | --- | --- | --- | --- | --- |
| Bracale et al, 2012 | N | + | ? | N | + | + |
| Brondan et al i, 2012 | + | + | ? | N | + | + |
| Brondani et al, 2014a | N | + | ? | N | + | + |
| Brondani et al, 2014b | + | + | ? | N | + | + |
| Cha et al, 2008 | N | + | - | N | + | + |
| Chathoth et al, 2018 | + | + | ? | N | + | + |
| Chen et al, 2015 | + | + | ? | N | + | + |
| Csernus et al, 2014 | + | + | ? | N | + | + |
| de Souza et al, 2013 | + | + | ? | N | + | + |
| Dhall et al, 2012 | N | + | ? | N | + | - |
| Dong et al, 2020 | + | + | ? | N | + | + |
| Elfasakhany, 2020 | - | + | ? | N | + | + |
| Esterbauer, 1998 | + | + | ? | N | + | + |
| Forga, 2003 | + | + | ? | N | + | + |
| Franco-Hincapie, 2009 | + | + | ? | N | + | + |
| Fukuyama, 2006 | N | + | ? | N | + | + |
| Gagnon 1998 | + | + | ? | N | + | + |
| Hamada, 2009 | + | + | ? | - | + | + |
| Heilbronn, 2000 | N | + | ? | N | + | + |
| Jin 2020 | - | + | ? | N | + | + |
| Kiec-Wilk, 2002 | N | + | ? | N | + | + |
| Kotani, 2008 | + | + | ? | N | + | + |
| Kotani, 2011 | + | + | ? | N | + | + |
| Labruna, 2009 | + | + | ? | N | + | + |
| Lim, 2012 | + | + | ? | N | + | + |
| Lin, 2009 | + | + | ? | N | + | + |
| Lindholm, 2004 | + | + | ? | N | + | + |
| Malczewska-Malec, 2004 | N | + | ? | N | + | + |
| Montesanto, 2018 | + | + | ? | N | + | + |
| Mori, 2001 | + | + | ? | N | + | ? |
| Mottagui-Tabar, 2008 | + | + | ? | N | + | ? |
| Nakatochi, 2015 | + | + | ? | N | + | + |
| Nicoletti, 2016 | N | + | ? | N | + | + |
| Nieters, 2002 | + | + | ? | N | + | + |
| Oh, 2004 | N | + | ? | N | + | + |
| Pei, 2017 | + | + | ? | N | + | + |
| Proenza, 2000 | + | + | ? | N | + | + |
| Rudofsky, 2006 | + | + | ? | N | + | + |
| Rudofsky, 2007 | N | + | ? | N | + | + |
| Sale, 2007 | N | + | ? | N | + | + |
| Samano, 2012 | + | + | ? | N | + | + |
| Schaffler, 1999 | N | + | ? | N | + | ? |
| Sivenius, 2000 | ? | + | ? | + | + | + |
| Sramkova, 2007 | + | + | ? | N | + | + |
| Sun, 2018 | + | + | ? | N | + | + |
| Tiwari, 2009 | + | + | ? | N | + | + |
| Verdi, 2020 | + | + | ? | N | + | + |
| Vimaleswaran, 2007 | + | + | ? | N | + | + |
| Vimaleswaran, 2010 | + | + | ? | N | + | + |
| Yiew, 2010 | + | + | ? | N | + | - |
| Zhang, 2015 | + | + | ? | N | + | + |
| Zietz, 2001 | N | + | ? | N | + | + |
| Key: + = low risk of bias; – = high risk of bias; ? = unclear risk of bias; N = not-applicable. | | | | | | |

**Figure S4:** Summary of risk of bias assessment**.**

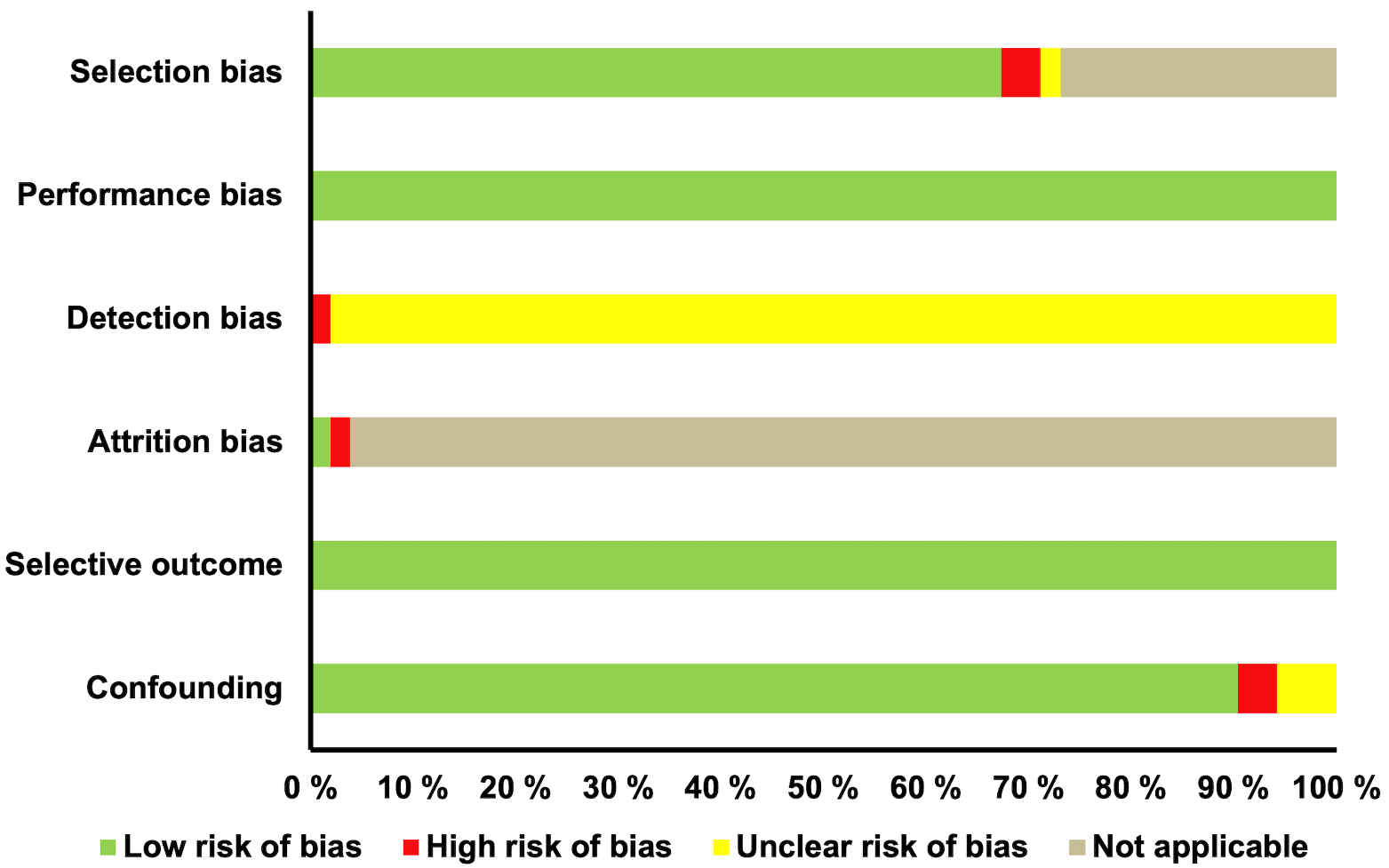

**Table S17:** Data extraction.

| **First Author-Year** | **Design** | **Methods** | **Participants** | **Outcomes** |
| --- | --- | --- | --- | --- |
| 1.Bracale et al,2012 [6] | Case-only | Genotyping analysis by TaqMan assay by Real-Time PCR; A-3826G *UCP1* SNP. | Italians (n=112; m=40, f=72; age=32.7±10.5 years; BMI=48.5±7.5 kg/m^2^.  **Group 1:** severely obese non-diabetic individuals, IR+ (n=50)  **Group 2:** severely obese non-diabetic individuals, IR- (n=62). | 1. The A-3826G (rs1800592) genotypes were reported more in the IR+ positive (88%) than in IR- (63%) obese individuals (OR= 4.3, 95% CI= 1.6-11.7 p=0.003). 2. Absence of A-3826G UCP1 polymorphism displayed high negative predictive value (100%) for IR. |
| 2.Brondani et al,2012 [7] | Case-control | Genotyping analysis by PCR-RFLPs; A-3826G *UCP1* SNP. | European ancestry, Type I diabetics (n=257)  **Group 1:** Patients with Diabetic retinopathy (n=154), age=39.2±12.1 years, BMI= 23.6±4.9kg/m^2^  **Group 2:** Patients without Diabetic retinopathy (n=103), age=33.32±13.8 years; BMI= 22.9±5.1 kg/m^2^  **Group 3:** Healthy controls (n=29), age=44±7.8 years | 1. Genotype frequencies Group1: AA=37%, AG=43.5%, GG=19.5% 2. Genotype frequencies Group 2: AA= 43.7%, AG=49.5%, GG=6.8% 3. G allele frequency Group 3 =33% 4. *UCP1* A-3826G GG genotype is associated with an increased risk of DR in type 1 DM patients. |
| 3.Brondani et al, 2014a [8] | Case-control | Genotyping analysis by TaqMan assay by Real-Time PCR; A-3826G *UCP1* SNP. | Brazilians (n=765)  **Group 1:** non-obese+T2DM (n=483); m=53.1%, f=46.9%; age=59.2±10.7 years; BMI= 25.8±2.8 kg/m^2^  **Group 2:** obese+T2DM (n=282); m=37.4%, f=62.6%; age=57.3±10.0 years; BMI= 34.4±4.3 kg/m^2^ | 1. A-3826G genotype frequencies in group 1 were for AA 46.7%, for AG 42.7%, and GG 10.6%; in group 2 were 49.4%, 38.6%, and 12%, respectively (p=0.529). 2. -3826G allele frequency in group 1 and group 2 was 0.319 and 0.313, respectively (p=0.839). 3. No difference in the allelic and genotypic distributions between group 1 and group 2 (p>0.05). |
| 4.Brondani et al, 2014b [9] | Case-Control | Genotyping analysis by PCR-fast real system; A-3826G *UCP1* SNP. | Brazilians with Caucasian ancestry (n=1576)  **Group 1:** non-diabetic (n=561)  **Group 2:** T2DM individuals (n=1015); m=456, f=559; age=59.5±10.5 years; BMI=28.7±5.3 kg/m^2^. Obesity was present in 35.9% of participants. | 1. Among T2DM individuals, A-3826G genotype frequencies in group 1 were for AA 49.3%, AG 39.5%, and GG 11.2%; in group 2 were 49.8%, 37.7%, and 12.4%, respectively (p= 0.694). 2. -3826G allele frequency in group 1 and group 2 was 0.310 and 0.314, respectively (p = 0.851). 3. No difference in SBP and DBP, BMI, waist circumference, TC, HDL-C, LDL-C, HbA1c, FPG, TG levels between A-3826G genotypes (p>0.05). |
| 5.Cha et al, 2008 [10] | Case-only | Genotyping analysis by TaqMan assay by Real-Time PCR; A-1766G, A-3826G and Ala64Thr (+1068G/A) *UCP1* SNPs. | Koreans (n=832)  Obese females (n=832); age=27.88±7.80 years; BMI= 25.89±4.27 kg/m^2^. | 1. A-1766G (rs3811791) genotype distribution were for AA n=458, AG n=307, GG n=63. 2. Ala64Thr (rs45539933) genotype distribution were AA n=706, AG n=111, GG n=5. 3. A-3826G (rs1800592) genotype distribution were AA n= 216, AG n=406, GG=209. 4. -1766G allele frequency was 0.27. 5. Ala64ThrT allele frequency was 0.07. 6. -3826G allele frequency was 0.49 7. AG genotype of A-3826G SNP is associated with SBP in dominant model (p=0.042) and DBP in co-dominant model (p=0.035). 8. No association between A-1766G and Ala64Thr genotypes and SBP and DBP in any of the inheritance models studied (Co-dominant, Dominant and Recessive). |
| 6.Chen et al, 2015 [11] | Case-control | Genotyping analysis by PCR-fast real system; A-3826G *UCP1* SNP. | Chinese (n=418)  **Group 1:** non-obese (n=169); m=93, f=76; age=53.35±11.78 years; BMI=20.74±1.6 kg/m^2^.  **Group 2:** overweight/obese (n=249); m=149, f=100; age=55.04±10 years; BMI= 25.72±2.25 kg/m^2^. | 1. A-3826G genotype frequencies in group 1 were for AA 0.225, AG 0.479, GG 0.296; in group 2 were: 0.249, 0.454, and 0.297, respectively (p>0.05). 2. -3826G allele frequency in group 1 and group 2 was 0.524 and 0.536, respectively (p = 0.746). 3. Among overweight/obese individuals, -3826AG heterozygotes showed higher serum mean concentration of TG, apo C-II and apo C-III compared with t-3826AA homozygotes (p < 0.05). 4. No association was found of the A-3826G polymorphism and low HDL-cholesterolemia in overweight/obese individuals (p>0.05). 5. No association was found between UCP1 A-3826G polymorphism and overweight/obesity. |
| 7.Chathoth et al, 2018 [12] | Case-Control | Genotyping analysis by TaqMan assay by Real-Time PCR; A-3826G, A-1766G, A-112C, *UCP1* SNPs | Saudi Arabians (n=492)  **Group 1:** non-obese (n=155); m=76, f=79; age=43.86±14.54 years; BMI=24.09±2.6 kg/m^2^. T2DM was present in 36.12% of participants.  **Group 2:** obese T2DM (n=235) + obese hypertensive (n=85). The group was subdivided into: a. moderate-obese BMI≥30-39.9 kg/m^2^); m=4.96%, f=56.03%; age=50.45±11.17 years; BMI=34.15±2.6 kg/m^2^.  b. extreme-obese, BMI≥40 kg/m^2^; m=36.15%, f=63.84%; age=42.57±13.72 years; BMI= 48.26±11.94 kg/m^2^.  T2DM was present in 69.73% of participants, and hypertension was present in 25.22% of participants. | 1. Among the obese (n=231), the -3826G allele frequency was higher as compared with non-obese (83) (OR=1.52, 95% CI= 1.10-2.08; p=0.009) (adjusted for age, gender and BMI). 2. -1766G allele was associated with the moderate-obesity (OR=2.89, CI=1.33–6.25; p=0.007), but not with extreme obesity (adjusted for age, gender and BMI). 3. -3826G allele frequency was higher in moderate-obese cohort with abnormal HDL, LDL, and hypertriglyceridemia; for hypercholesterolemia, -3826G allele frequency was higher in the extreme-obese cohort. 4. -1766G allele frequency was higher in the moderate-obese with high LDL. 5. -3826G allele frequency was higher in the moderate-obese with T2DM and hypertension. 6. -3826G allele frequency was higher in the extreme-obese males ≤35 years, and higher in the moderate-obese males > 35 years. 7. -1766G allele frequency was higher in females aged ≤35 years with extreme obesity, and in males aged ≤35 years with moderate obesity. |
| 8.Csernus K. et al 2014 [13] | Case only | Genotyping analysis by PCR/PCR-RFLP; A-3826G, *UCP1* SNP. | Obese Hungarian children (n=528),  Age:13.2±2.6 years,  BMI: 30.6±4.6kg/m^2^, m=297 and f=231. | 1. No significant differences in measures of obesity adjusted BMR, obesity related metabolic parameters or blood pressure values according to *UCP1* A-3826G. 2. Genotype frequencies: AA=51.1%, AG= 40.5% and GG=8.3%. 3. Minor allele frequency= 0.29 |
| 9.de Souza 2013 [14] | Case-Control | Genotyping analysis by TaqMan assay by Real-Time PCR; A-3826G *UCP1* SNP. | Brazilians with European ancestry (n=1515)  **Group 1:** non-diabetic (n=534); m=55%, f=45%; age=44.0±7.8 years.  **Group 2:** T2DM (n=981); m= 47.4%, f=52.6%; age=59.52±10.63 years; BMI=28.84±5.39 kg/m^2^. | 1. A-3826G genotype distributions in group 1 were: for AA 49.3%, AG 39.5%, GG 11.2%; in group 2 were: 49.9%, 37.7%, 12.4%, respectively (p=0.694). 2. -3826G allele frequency in group 1 and group 2 was 0.310 and 0.313, respectively (p=0.510). 3. A-3826G allele frequencies did not differ between diabetic and non-diabetic cohorts even when assuming dominant, recessive, additive or co-dominant models of inheritance (p>0.05). |
| 10.Dhall et al, 2012 [15] | Case-only | Genotyping analysis by PCR-fast real system; A-3826G *UCP1* SNP | Indians (n= 96)  Individuals with metabolic syndrome; m= 49, age=44±17 years; f=47, age=48±17 years. | 1. A-3826G genotype frequencies were: for AA 39.9%, for AG 46.5%, and for GG 13.5%. 2. Among females, the -3826GG homozygotes showed higher BMI, SBP and DBP (p<0.001); among males, homozygotes for -3826A allele showed higher DBP (p<0.001). 3. In female -3826GG homozygotes DBP was correlated with waist circumference and WHtR (p<0.05); in female -3826 AG heterozygotes SBP and DBP were correlated with WC, fat %, BMI, WHtR and WHR (p<0.001). 4. Among males, no association with obesity markers and blood pressure between A-3826G genotypes. |
| 11.Dong et al, 2020 [16] | Case-Control | Mass ARRAY genotyping system, A-1766G *UCP1* SNP | T2D individuals(n=928), 39.2% male and 60.8% female subjects, 60.9±10 years, 25.1±3.6 kg/m^2^  Healthy Controls (n=1034), 37.7% male and 62.3% female subjects, 60.0±9.5 years, 24 ±3.2 kg/m^2^ | 1. rs3811791 CC variant genotype conferred a   significantly increased risk of T2DM and a higher level of TG   2. rs3811791 of UCP1 may be associated with   T2DM and TG.   3. SB interacted with rs3811791 of UCP1 was   associated with T2DM,   4. PA interacted with rs3811791 of UCP1 was   associated with the level of HOMA-IR, HDL-C, and TG, suggesting that it is of paramount importance for people to take regular physical exercise |
| 12.Elfasakhany et al, 2020 | Case control | Genotyping analysis by PCR-RFLP, A-3826G *UCP1* SNP | Saudi Arabians (n=218)  **Group 1:** control healthy (n=110), age=39.1 ± 6.07 years, BMI=23.47±1.44 kg/m^2^  **Group 2** (n=108) Type 2 diabetes patients, age= 41.2±6.88 years, BMI=23.61± 1.23 kg/m^2^ | 1. Control group🡪AA=50.9%, AG=40.9%, GG=8.2% 2. Type 2 diabetes group🡪 AA=47.22%, AG=39.82%, GG=12.96% 3. No significant difference in the genotype frequency between subjects with T2DM and healthy controls (P > 0.05). 4. No significant difference in the allele frequency between both T2DM subjects and healthy controls (P > 0.05) 5. They suggest that UCP1 A/G polymorphism at −3826 promoter region may not contribute to higher susceptibility to the T2DM in the Saudi population of Makkah region. |
| 13.Esterbauer et al, 1998 [17] | Case- only | Genotyping analysis by PCR-RFLPs, A-3826G *UCP1* SNP | Caucasians (n=153) | 1. Genotype frequencies: AA=70, AG=66, GG=8 2. -3826G polymorphism is probably a marker for expressional differences, but not the causative mutation. 3. UCP-1 gene locus is identified as a common cause of reduced UCP-1 gene activity in obese subjects. |
| 14.Forga, et al, 2003 [18] | Case-Control | Genotyping analysis by PCR-fast real system; A-3826G *UCP1* SNP. | Spanish (n=313)  **Group 1:** non-obese (n=154); BMI=22.3±1.8 kg/m^2^  **Group 2:** obese (n=159); BMI=37.6±5.7 kg/m^2^  Age=20-60 years. | 1. A-3826G genotype distribution in group 1 was 63.6% for AA, 31.2% for AG, 5.2% for GG; in group 2 was 66.7%, 28.9%, and 4.4%, respectively (p=0.574). 2. No differences in UCP1 -3826G allele frequency between group 1 (0.21) and group 2 (0.19) individuals (p=0.574) 3. In obese group, -3826G allele carriers had higher BMI (p<0.05), fat % (p<0.05), SBP (p< 0.01), DBP (p<0.05). |
| 15.Franco-Hincapie et al, 2009 [19] | Case-Control | Genotyping analysis by PCR-fast real system; A-3826G and Ala64Thr UCP1 SNPs. | Colombians (n=994)  **Group 1:** non-diabetic (n=449); m=126, f=323; BMI=25.2±3.8 kg/m^2^; age>40 years.  **Group 2:** T2DM (n=545); m=190, f= 355; BMI=27±4.6 kg/m^2^. | 1. Association between A-3826G allele and T2DM (OR=0.78; 95% CI: 0.63-0.97; p=0.02). |
| 16.Fukuyama et al, 2006 [20] | Case only | Genotyping analysis by TaqMan assay by Real-Time PCR; A-3826G and A-112C, UCP1 SNPs. | Japanese (n=93)  T2DM; m=55, f=38; age=56.6±13.5 years; BMI=25.6±4 kg/m^2^. | 1. A-3826G genotype frequencies were: for AA 32.3%, for AG 48.4%, and for GG 19.3%. 2. A-112C genotype frequencies were: for AA 88.2%, for AG 10.7%, and for GG1.1%. 3. Carriers of -112C allele showed higher levels of fasting plasma immune-reactive insulin concentration (p=0.0085), HOMA-IR (p= 0.0089), and hepatic lipid content (p= 0.012). 4. No association was found of the A-3826G UCP1 polymorphism and any measured clinical parameters. |
| 17.Gagnon et al, 1998 [21] | Case control | Genotyping analysis by PCR-RFLP, 3826A/G UCP1 SNP | Swedish (n=985)  **Group 1:** Obese (n=684)  **Group 2:** control subjects (n=311) | **Allele frequencies**   1. Control Group: A allele=473 G allele=149 2. Obese group: A allele= 1013, G allele=335   **Genotype frequencies**   3. Control group: AA=185, AG=103, GG=23 4. Obese group: AA=384, AG=245, GG=45 5. In both genders, there was no difference between carriers and non-carriers for variables pertaining to weight history. |
| 18.Hamada et al. 2009 [22] | Case only | Genotyping analysis by PCR, 3826A/G UCP1 SNP | Japanese obese healthy women (n=32), mean ± S.D.; age 49.9 ± 8.45 years; BMI 28.4 ± 3.3 kg/m^2^ | 1. The distribution of the A/A, A/G, and G/G genotypes was 18%, 49%, and 33%, respectively. 2. No difference in the changes of any physiological and metabolic parameters between the subjects with and without the A allele 3. No difference in the changes of any physiological and metabolic parameters between the subjects with the A/G genotype and those with the G/G genotype 4. |
| 19.Heilbronn et al 2000 [23] | Case-only | Genotyping analysis by PCR-fast real system; A-3826G UCP1 SNP. | Australians (n=526)  Female overweight/obese; BMI=34.1 kg/m^2^. | 1. A-3826G genotype frequencies were: for AA 0.307, for AG 0.190, and for GG 0.29. The -3826G allele frequency was 0.23. 2. -3826G allele was associated with higher BMI (p=0.02), insulin (p=0.03), and higher fasting glucose concentrations (p = 0.01). 3. Among T2DM females, -3826G allele carriers were more frequent (p=0.02) and was related with higher fasting glucose concentrations (p = 0.02). |
| 20.Jin P. et al 2020 [24] | Case-control | Genotyping analysis: GWAS, TaqMan assay by Real-Time PCR; A-112C and A-3826G UCP1 SNPs | Han Chinese (n=3107)  **Case**= 662 T2D patients with diabetic retinopathy (DR),  **Control**=2445 T2D patients without diabetic retinopathy | 1. Genotype frequencies: A112C: AA 🡪 DR=523, NDR=2046, AC🡪 DR=127, NDR=342, CC🡪 DR=4, NDR=10 2. rs10011540 (A-3826G) of the UCP1 gene is marginally significantly associated with DR |
| 21.Kiec-Wilk et al 2002 [25] | Case-only | Genotyping analysis by PCR-fast real system; A-3826G UCP1 SNP. | Polish (n=118)  Overweight/obese; m=38, f=80; age=43.4±19.3 years; BMI=33.21±7.73 kg/m^2^. | 1. A-3826G 1 genotype frequencies were: for AA 51.38%, for AG 33.94%, and for GG 14.68%. 2. -3826G allele frequency was 30.5%. 3. No association was found of -3826G allele with BMI and glucose tolerance. 4. -3526GG homozygotes showed higher fasting levels of TG (p=0.04) and those recorder at 6hrs of OLTT (p=0.058), and the lower HDL levels compared with -3826AA homozygotes (p=0.004). 5. Free fatty acids increased in -3826GG homozygotes, especially at 8 hrs post-OLTT (p=0.031). 6. Carriers of the -3826G allele showed increased LDL levels as compared with -3826AA homozygotes (p=0.027). 7. -3826G allele carriers showed higher beta-tromboglobulin levels compared with -3826AA homozygotes (pAA:A/G= 0.012; pAA:GG=0.055). |
| 22.Kotani et al,2008 [26] | Case only | Genotyping analysis by PCR-RFLPs, 3826A/G UCP1 SNP. | Japanese (n=298), age= 45.2±7.2 years, male=144, female=154 | 1. Genotype frequencies of UCP-1 genotypes were 26.5, 51.3, and 22.2% for AA, AG, and GG, respec tively. 2. Allelic frequency of 0.48 for the G allele. 3. In males, HDL-C levels increased in the order of AA AG GG genotypes, and the levels in GG genotypes (1.75 ± 0.49 mmol/L) were significantly higher than those in the AA genotype (1.45 ± 0.34 mmol/L, p 5 0.015), whereas this trend was non-significantly detected in females. 4. GG genotype may be an independent protective factor associated with low HDL-cholesterolemia in healthy Japanese individuals. |
| 23.Kotani et al,2011 [27] | Case-Control | Genotyping analysis by an intercalater-mediated  fluorescent allele-specific PCR method; A-3826G UCP1 SNP. | Japanese (n=294)  **Group** **1:** non-obese (n=192).  **Group 2:** obese (n=102).  Age=65±13 years. | 5. A-3826G genotype distributions in group 1 were: for AA 31%, AG 46%, and GG 23%; in group 2 were: 31%, 27%, and 27%, respectively (p=0.79). 6. The frequency of the -3826G allele was 0.47. 7. Among obese, -3826 GG homozygotes were more frequent (OR: 6.85, 95% CI: 1.65-28.49; p<0.01). 8. Obese carriers of -3826GG genotype showed higher prevalence of low HDL-cholesterolemia (37%) than those with the AA and AG genotypes (13%) (p<00.1). 9. Obese -3826GG homozygotes showed lower HDLC levels (1.20±0.30) than those carriers of the -3826 AA and AG genotypes (1.39±0.36; p=0.01). |
| 24.Labruna et al,2009 [28] | Case-Control | Genotyping analysis by TaqMan assay by Real-Time PCR; A-3826G UCP1 SNP. | Italians (n= 197)  **Group 1:** non-obese (n= 95); m=29, f=66; BMI>20 and <25 kg/m^2^, respectively.  **Group 2:** obese (n=102); m=41, f=61; age 34.5 and 31 years, respectively; BMI=47.9 and 47.7 kg/m2, respectively.  Metabolic syndrome was present in 53% males and 66% females, and hypertension was present in 73% males and 31% females. | 1. A-3826G frequencies in group 1 were: for AA 54.8%, for AG 34.7%, and for GG 10.5%; in group 2 were: 50%, 41.2%, and 8.8%, respectively. 2. -3826G allele frequency in group 1 and group 2 was 0.28 and 0.29, respectively. 3. In participants with severe liver steatosis -3826 AG and GG genotypes were more frequent than in those with mild/moderate liver steatosis (21/31; 65% vs 30/70; 43%, p=0.0003). 4. 3826 AG + GG genotypes did not differ among metabolic syndrome+ and metabolic syndrome- obese (46% vs 56%) (p>0.05). |
| 25.Lim et al,2012 [29] | Case-Control | Genotyping analysis by TaqMan assay by Real-Time PCR; A-3826G, A-1766G and Ala64Thr UCP1 SNPs. | Koreans (n= 2180)  **Group 1:** heathy (n=587); m=273, f=314; age=64 years; BMI=24.17 kg/m^2^.  **Group 2:** obese with CIN and DP+ (n= 583); m=314, f=331; age=69 years; BMI=24.6 kg/m^2^.  **Group 3:** obese with CIN and DP- (n=1010); m=619, f=523; age=70 years; BMI=23.28 kg/m^2^. | 1. A-3826G minor frequency allele for groups 1, 2 and 3 were 46.73, 48.66, and 50.43, respectively. 2. A-1766G minor frequency allele for groups 1, 2, and 3 were 24.78, 24.4, and 24.09, respectively. 3. Ala64Thr minor frequency allele for groups 1, 2, and 3 were 6.58, 6.45, and 7.84, respectively. 4. Carriers of the -1766AG + GG genotypes were more frequent in DP+ group compared with the normal group in the dominant model (77.76% in DP+ vs.71.77% in normal, OR=1.508, p=0.006, power=85.3%). 5. -1766G allele frequency was lower in the DP- group compared with the normal group in the recessive model (4.77% in DP- vs. 5.10% in normal, OR = 0.606, p =0.0423, power=56.9%). 6. -1766GG homozygotes were less frequent in DP- compared with the normal group in the recessive model (OR=0.606, p=0.042). 7. Carriers of the -3826G allele showed higher serum HLC-C levels in the dominant models (p=0.032). 8. Serum TGs and HDL-C levels were associated with the -1766G allele in the recessive model (p=0.002; p=0.046, respectively). |
| 26.Lin et al,2009 [30] | Case-Control | Genotyping analysis by TaqMan assay by Real-Time PCR; A-3826G UCP1 SNP. | Taiwanese (n=575)  **Group 1:** Non-obese T2DM (n=191); m= 95, f=96; age=57.8±9 years; BMI=22.4±2 kg/m^2^.  **Group 2:** Non-obese controls (n=135); m=56, f=79; age=57.1±10.8 years; BMI= 22.1±1.8 kg/m^2^.  **Group 3:** Obese with T2DM (n=198); m=100, f=98; age=56.9±10.1 years; BMI=28.2±3.1 kg/m^2^.  **Group 4:** Obese controls (n=51); m=21, f=31; age=57.4±10 years; BMI=27.4±2 kg/m^2^. | 1. A-3826G genotype frequencies in group 1 were: for AA, 42%, for AG 79%, for GG 57%; in group 2 were 24%, 54%,30%, respectively (p=0.449). 2. In non-obese - as determined by BMI, the A-3826G polymorphism was not associated with T2DM. 3. A-3826G genotype frequencies in group 3 were for AA 44%, for AG 91%, and for GG 49%; in group 4 were 10%, 15%, and 12%, respectively (p=0.277). 4. In obese- as determined by BMI, no association was found with T2DM for A-3826G polymorphism. |
| 27.Lindholm et al, 2004 [31] | Case-Control | Genotyping analysis by PCR- fast real system; A-3826G UCP1 SNP. | Scandinavians (n=540)  **Group 1:** non-diabetic (n=106); m= 61, f=45; age=55.0±14.1 years; BMI=26.2±4.6 kg/m^2^. **Group 2:** diabetic + normoalbuminuria (n=218); m=118, f= 100; age=54.3±14.7 years; BMI=25.1±3.9 kg/m^2^.  **Group 3:** diabetic + micro- or macroalbuminuria (n=216); m=117, f=99; age=55.9±14.6 years; BMI=26.6±4.3 kg/m^2^. | 1. A-3826G genotype frequencies in group 1 were for AA 64.2% and for AG/GG 35.8%, in group 2 were 55.5% and 44.5%, respectively; in group 3 were 61.1% and 38.9%, respectively. 2. No differences in allele and genotype frequencies in the 3826A/G between the groups were found. 3. Carriers of the -3826G allele showed lower HDL-C levels (p=0.01). |
| 28.Malczewska-Malec et al,2004 [32] | Case-only | Genotyping analysis by PCR- fast real system; A-3826G UCP1 SNP. | Southern Polish (n=122)  Members of obese families; m=38, f=84; age=43±19 years.  Obese (BMI≥30 kg/m^2^), Overweight  (BMI≥25 kg/m^2^) were 44% and 25%, respectively. | 1. No differences in glucose tolerance parameters between the A-3826G genotypes 2. No association between A-3826G polymorphism (i.e. AA and C allele carriers) and BMI and insulin resistance |
| 29.Montesanto et al, 2018 [33] | Case-Control | Genotyping analysis by SEQUENOM  MassArray iPLEX technology; Ala64Thr, A-1766G and A-3826G UCP1 SNPs. | Italians (n=940)  **Group 1:** non-diabetic (n=505); m=41.2%, f= 58.8%; age=58.59±12.2 years; BMI= 27.1kg/m^2^.  **Group 2:** T2DM (n=435); m= 56.6%, f=43.4%; age=65.71±7.9 years; BMI=28.7 kg/m^2^. The group was subdivided based on the presence and absence of retinopathy (yes:111/no:324) and nephropathy (yes:54/no:381). | 1. No association of Ala64Thr (C/T) polymorphism with T2DM (p=0.969). 2. Ala64ThrT allele was less frequent in individuals with coexisting diabetic retinopathy compared to those without (OR=0.31, 95% CI=0.12-0.82; p=0.010). 3. -3826G allele was less frequent in individuals with nephropathy compared with those without (OR=0.55, 95% CI=0.33-0.98; p=0.031). |
| 30.Mori et al, 2001 [34] | Case-Control | Genotyping analysis by PCR- fast real system; A-112C UCP1 SNP. | Japanese (n=570)  **Group 1:** healthy controls (n=250); m=145, f=105; age=76.4±7.9 years; BMI=20.9±3.4 kg/m^2^.  **Group 2:** T2DM (n=320); m=180, f=140; age=62.9±11.8 years; BMI=23.1±3.5 kg/m^2^. | 1. - 112A/C genotype distributions in group 1 were for AA 220, for AC 29, and for CC 1; in group 2 were 257, 61, and 2, respectively. 2. -112C allele frequency was higher in T2DM (10.2%) than in controls (6.2%) (p= 0.017). 3. A-3826G genotype distributions in Grp1 were for AA 58, for AG 116 and for GG 76; in Grp2 were 83,156, and 81, respectively. 4. The frequency of the -3826G allele did not differ between T2DM (49,7%) and controls (53,6%) (p=0.190). |
| 31.Motaggui-Tabar et al, 2008 [35] | Case-only | Genotyping analysis by Dynamic allele specific hybridization and TaqMan assay by Real-Time PCR; A-3826G UCP1 SNP. | Swedish (n=773)  **Group 1:** healthy controls (n=481); Females; BMI=23±3 kg/m^2^.  **Group 2:** obese (n=292); Females; BMI=39±5 kg/m^2^. | 1. A-3826G genotype frequencies in group 1 were for AA 59%, for AG 36%, and for GG 5%; in group 2 were 55%, 38%, and 7%, respectively (p=0.53). 2. No allele and genotype differences between the obese and controls. 3. A-3826G polymorphism was not associated with BMI, waist circumference, serum insulin or insulin sensitivity (p>0.05). |
| 32.Nakatochi et al,2015 [36] | Case-Control | Genotyping analysis by DigiTag2 assay; A-3826G *UCP1* SNP. | Japanese (n=2343)  **Group 1:** Controls  with no metabolic syndrome in 2001 and 2009 (n= 1983); Males; BMI=21.6 kg/m^2^.  **Group 2:** no metabolic syndrome in 2001, metabolic syndrome in 2009 (n= 360); BMI=24.6 kg/m^2^. | 1. -3826G allele frequency in groups 1 and 2 were 0.502 and 0.456, respectively (p= 0.022). 2. A-3826G polymorphism was associated with metabolic syndrome (OR=0.83; 95%CI=0.70-0.97; p=0.022). |
| 33.Nicoletti et al 2016 [37] | Case-only | Genotyping analysis by TaqMan assay by Real-Time PCR; A-3826G *UCP1* SNP. | Mixed ethnicity (n=150)  Obese; m=20%, f= 80%; age=47.2±10.5 years; BMI ≥35 kg/m^2^. | 1. A-3826G genotype frequencies were for AA 41.3%, for AG 45.3%, and for GG 13.4%. 2. -3826G allele frequency was 0.36. 3. Carriers of the -3826G allele had lower weight, body fat, and fat free mass for the dominant model (p<0.05). 4. -3826GG homozygotes showed lower frequency of T2DM compared with those carriers of -3826AA + GG genotypes. 5. A-3826G polymorphism was associated with weight [r^2^:0.417, 95% CI: (-20.020 to -2.259); p=0.015] and with FM [r^2^: 0.339, 95% CI: (-15.314 to –2.077); p=0.011] following a multiple regression model adjusted for sex, age, height, physical activity, and energy intake. 6. A-3826G polymorphism was associated with FFM following a simple linear regression model [r^2^=0.288, 95% CI: (-8.110 to -1.238); p=0.008]. 7. A-3826G polymorphism was associated with weight [r^2^: 0.094, 95%CI: (-25.421 to -4.752); p= 0.005], and FFM [r^2^: 0.228, 95%CI: (-8.110 to -1.238); p= 0.008] following a linear regression model. |
| 34.Nieters et al 2002 [38] | Case- control | Genotyping analysis by PCR-RFLPs, 3826A/G *UCP1* SNP. | Germans (n=308)  **Group 1**: normal weight (n=154) age=51.3±8.5 years; BMI= 23±1.6 kg/m^2^  **Group 2**: grade II and III obese (n=154) age=51.2±8.4 years; BMI= 38.2±2.8 kg/m^2^ | 1. Genotype percent in Group 1: AA=54.9%, AG=41.2%, GG=3.9% 2. Genotype percent in Group 2: AA=54.5%, AG 41.6%, GG=3.9%/ |
| 35.Oh et al 2004 [39] |  | Genotyping analysis by PCR- fast real system; A-3826G *UCP1* SNP. | Koreans (n=190)  Obese; m=44, f=146; age=28.38±0.72 years; BMI= 33.88±0.28 kg/m^2^. | 1. A-3826G UCP1 genotype distribution was for AA 22.1%, for AG 53.7%, and for GG 24.2%. 2. The frequency of the -3826G allele was 0.51. 3. Carriers of the -3826AG+GG genotypes showed higher DBP compared with those carriers of the -3826AA genotype (p=0.023). 4. LDL cholesterol levels were higher in obese carriers of the -3826G allele compared with -3826AA homozygotes type (p=0.011) 5. HDL cholesterol levels were lower in -3826GG homozygotes compared with those carriers of the -3826AA+ AG genotypes (p=0.042). 6. The atherogenic index was 22.8% higher in -3826GG homozygotes compared with those carriers of the -3826AA genotype (p=0.027). 7. Obese -3826GG homozygotes showed higher LDL/HDL compared with those carriers of the -3826AA genotype (p=0.001). 8. When obese group was further divided into a normal group and a hyper-LDL cholesterolemia group, the frequency of -3826GG genotype was higher in the hyper 9. LDL cholesterolemia group (71.4%) than in normal group (43.9%) (p=0.05). 10. The frequency of hyper-LDL cholesterolemia was higher in -3826GG genotype carriers (25.6%) compared with those in -3826AA genotype carriers (9.8%) (p=0.05). 11. -3826GG genotype (OR= 4.115; p=0.03) and body fat mass (OR= 1.079; p=0.03), were risk factors of hyper-LDL cholesterolemia. |
| 36.Pei et al 2017 [40] | Case-only | Genotyping analysis by ligase detection reaction; A-3826G and Ala64Thr *UCP1* SNPs. | Chinese (n=528)  **Group 1:** normal fasting plasma glucose (n=445); m=174, f=271; age=51.57±13.13 years; BMI=24.00±3.54 kg/m^2^.  **Group 2:** impaired fasting glucose + T2DM (n=83); m=24, f=59; age=56.71±11.96 years; BMI=26.06±3.56 kg/m^2^. | 1. Ala64Thr genotype frequencies in group 1 were for CC 87.2%, for CT 12.6, and for TT 0.2%; in group 2 were 84.3%, 15.7%, and 0%, respectively (p>0.05). 2. Ala64Thr T allele frequency for groups 1 and 2 was 6.5% and 7.8%, respectively (p>0.05). 3. A-3826G genotype frequencies in group 1 were for AA 25.8%, for AG 52.6%, and for GG 21.6; in group 2 were 24.1%, 49.4%, and 26.5%, respectively (p>0.05). 4. -3826G allele frequency for groups 1 and 2 was 47.9% and 51.2%, respectively (p>0.05). 5. A-3826G and Ala64Thr genotype distributions and allele frequencies didn’t differ between the normal fasting plasma glucose group and the impaired fasting glucose + T2DM group using codominant, dominant, and recessive genetic models- even after adjusting for age, sex, drinking status, and BMI (p>0.05). |
| 37.Proenza et al, 2000 [41] | Case-Control | Genotyping analysis by PCR- fast real system; A-3826G *UCP1* SNP was studied. | Turkish (n=240)  **Group 1:** lean healthy (n=94); m=77, f=17; age=30±1years; BMI=22.3±0.2 kg/m^2^.  **Group 2:** obese (n=146); m=83, f=63; age=35±1years; BMI=37.8±0.5 kg/m^2^. | 1. A-3826G genotype frequencies in group1 were for AA 52.1%, for AG 35.1%, and for GG 12.8%; in group 2 were 47.8%, 43.4%, and 8.8%, respectively (p=0.370). 2. -3826G allele frequency in groups 1 and 2 were 0.30 and 0.31, respectively. 3. -3826GG homozygotes showed higher cholesterol levels associated with BMI compared with carriers of the -3826AA (p=0.027) and -3826AG (p=0.039) genotypes. |
| 38.Rudofsky et al,2007 [42] | Case-only | Genotyping analysis by PCR- fast real system; A-3826G *UCP1* SNP. | Caucasian with T2DM (n=517) | 1. A-3826G genotype frequencies were for AA 49.9%, for AG 45.6%, and for GG was, and 4.5%. 2. -3826G allele frequency was 0.27 for G. 3. Genotypic distribution did not differ with respect to the baseline clinical characteristics except of age (p = 0.03). 4. A-3826G genotypes were not associated with diabetes-related microvascular complications: [neuropathy: p = 0.79); (retinopathy: p = 0.48); (nephropathy: p = 0.93)]. |
| 39.Rudofsky et al, 2006 [43] | Case only | Genotyping analysis by PCR-RFLPs, A-3826G *UCP1* SNP. | Type 1 diabetes (n=227) | 1. 130 patients (57.3%) were (AA), 85 patients (37.4%) were (AG), and 12 (5.3%) were ho- mozygous for the polymorphism (GG) 2. No difference in genotype frequencies was found with respect to diabetes complications 3. No association of the A-3826G polymorphism in the UCP1 gene with diabetic neuropa- thy was observed, |
| 40. Sale et al, 2007 [44] | Case-only | Genotyping analysis by MassARRAY system; A-3826G and Ala64Thr *UCP1* SNPs. | **Group 1:** African Americans (n=287); m=43.5%, f= 56.5%; age=43.8±14.8 years; BMI=28.8±6.5 kg/m^2^.  **Group 2**: Hispanics (n=811), subdivided into:  Group 2a: Hispanics from San Antonio (n=493); m= 40%, f=60%, age=43.6±14.8; BMI= 30.1±6.3 kg/m^2^. Group 2b Hispanic  individuals from San Luis Valley (n= 318); m= 47.8%, f= 52.2%; age=40.3±14 years; BMI=27.5±5.6 kg/m^2^.  Diabetes prevalence for group 1, group 2a, and group 2b was 11.6%, 17.7%, and 12.8%, respectively. | 1. A-3826G polymorphism was associated with acute insulin response to glucose in African Americans-adjusted for age, sex, and BMI (p=0.017). 2. A-3826G polymorphism was associated with HDL-C levels in Hispanic families from San Antonio-adjusted for age, sex, BMI- (p=0.001). |
| 41.Samano et al, 2012 [45] | Case control | Genotyping analysis by Taq-Man PCR, A-3826G *UCP1* SNP | Mexican children (n=270)  m=173, f=142  **Group 1**= Normal weight n=159, age=16.6±1 year, BMI=21.1±1.9 kg/m^2^  **Group 2**= obesity, n=111, age=16.7±1.1 years, BMI=27.8±3.9 kg/m^2^ | 1. The UCP1A-3826G (rs 1800592) polymorphism was associated with high percentage of fat (p = 0.002) and muscle weight (p = 0.019) in a recessive model. 2. Normal weight group: AA=32.1%, AG=50.9%, GG=17% 3. Obesity Group: AA=27.9%, AG=55.9%, GG=16.2% |
| 42.Schaffler et al, 1999 [46] | Case-only | Genotyping analysis by PCR- fast real system; A-3826G *UCP1* SNP. | Germans (n=1020)  m=534, f=486; age=51.3±14.6 years; BMI=25.5±4.4 kg/m^2^. | 1. A-3826G UCP1 genotype frequencies for AA, AG, and GG were 57.0%, 35.4%, and 7.6%, respectively. 2. -3826G allele frequency 0.25. 3. No significant differences between the genotypes and age, gender, BMI, leptin, glucose, fasting insulin, C-peptide, HbA1c, diabetes, TC, and HDL-C (p>0.05). |
| 43.Sivenius et al, 2000 [47] | Case-Control | Genotyping analysis by PCR- fast real system; A-3826G *UCP1* SNP. | Finish (n=203)  **Group 1:** non-diabetic (n= 123); m=55, f=68; age=58.4±5.3 years; BMI=27.1±4.4 kg/m^2^.  **Group 2:** T2DM (n=70); m=38, f=32; age=60.1±5.8 years; BMI=30.5±5.2kg/m^2^. | 1. -3826G allele frequency in groups 1 and 2 was 34.1% and 38.6%, respectively. 2. No difference in the A-3826G polymorphism frequency between T2DM individuals and healthy controls. |
| 44. Sramkova et al, 2007 [48] | Case-Control | Genotyping analysis by PCR- fast real system; A-3826G *UCP1* SNP. | Czech (n= 415)  **Group 1:** healthy controls (n=120); m=42, f=78; age=32.5±11.0 years; BMI=23.3±3.8 kg/ m^2^.  **Group 2:** T2DM (n=295); m=112, f=183; age=58.8±7.0 years; BMI=30.5±5.5 kg/m^2^.  **Group 3**: healthy offspring of T2DM (n=113); m=41, f=72; age=38.2±10.4 years; BMI=25.5±4.2 kg/m^2^. | 1. -3826G allele frequency was 0.26 and not associated with increased risk of T2DM. 2. A-3826G genotype frequencies in group 1 were for AA 50.83%, for AG 40.83%, and for GG 8.33%; in group 2 were 53.22%, 42.03%, and 4.75%, respectively; in group 3 were 53.10%, 45.13%, and 1.77%, respectively. 3. Genotypic distribution did not differ between diabetics and controls (χ^2^ = 2.02; p = 0.36). 4. Among diabetic women, -3826AG +GG genotype carriers showed lower WHR (p=0.000) and WHeR (p=0.049) compared with diabetic women carriers of the -3826AA genotype. |
| 45. Sun et al, 2018 [49] | Case-Control | Genotyping analysis by Sequenom MassArray System; A-3826G UCP1 SNP. | Chinese (n=2207)  **Group 1:** normotensives (n=1045); m=373, f= 672; age=50.28±8.70 years; BMI=23.73±3.41 kg/m^2^  **Group 2:** hypertensives (n=1162); m=573, f=589; age=57.22±11.10; BMI=26.07±5.53 kg/m^2^. | 1. A-3826G genotype frequencies in group 1 were for AA 25.12%, for AG 50.82%, and for GG 24.06%; in group 2 were 25.2%, 52.01%, and 22.77%, respectively. 2. -3826G allele frequency in groups 1and 2 was 49.47% and 48.78%, respectively. 3. No association between A-3826G genotype and allele distributions and essential hypertension in co-dominant, dominant, and recessive models (p>0.05). |
| 46.Tiwari et al, 2009 [50] | Case-Control | Genotyping analysis by PCR- fast real system; A-3826G, A-112C, and Ala64Thr UCP1 SNPs. | Asian Indians (n=420)  **Group 1:**  T2DM and no history of kidney disease;  Group 1a: from south India T2DM cases (n=149); m=102, f=47; age=60.45±11.5 years; Group 1b: from north India T2DM cases (n=75); m=40, f=35; age=61.03±8.88 years.  **Group 2:** T2DM and chronic renal insufficiency. Group 2a: from south India T2DM cases with chronic renal insufficiency (n=106); m=81, f=25; age=55.97±11.5 years; Group 2b: from north India T2DM cases with chronic renal insufficiency (n=90); m=78, f=12; age=53.56±10.99 years. | 1. Among south-Indians A-112C and Ala64Thr polymorphisms were associated with the development of chronic renal insufficiency; In the north-Indians no association for the UCP1 polymorphisms studied was found. 2. Among south-Indians, 112AA homozygotes showed higher percentage of T2DM with chronic renal insufficiency compared with -3826AG and GG carriers (OR=2.076, 95%CI=1.1893.625; p=0.0089) 3. Among south-Indians, -112A allele was more frequent in T2DM with chronic renal insufficiency (OR=1.849, 95% CI=1.1422.994; p=0.012) 4. Among south-Indians, Ala64Thr CC homozygotes showed a higher percentage of T2DM with chronic renal insufficiency compared with carriers of Ala64Thr TC and TT genotypes (OR=2.585, 95%CI=1.318–5.072; p=0.0048). 5. Among south-Indians, Ala64Thr C allele was more frequent in T2DM with chronic renal insufficiency individuals (OR=2.099 95%CI=1.146–3.844; p=0.015). 6. A-3826G polymorphism was not associated with the development of chronic renal insufficiency. 7. A-112C genotype in south-Indians and C allele frequencies in group 1 were for AA 59.7%, for AC 35.3%, and for CC 5%, and for C allele 0.227; in group 2 were 75.5%, 21.7%, and 2.8%, and 0.137, respectively. 8. A-112C in north-Indians and C allele frequencies in group 1 were for AA 60%, for AC 36%, and for CC 4%, and for C allele 0.220; in group 2 were 65.5%, 28.9%, and 5.6%, and 0.137, respectively. 9. Ala64Thr T/C in south-Indians and T allele frequencies in group 1 were for TT 1.5%, for TC 29.4%, and for CC 69.1%, and for T allele 0.162; in group 2 were 2.1%, 12.6%, and 85.3%, and 0.084%, respectively. 10. Ala64Thr T/C in north-Indians and T allele frequencies in group 1 were for TT 1.3%, for TC 25.3% and for CC 73.3%, and for T allele 0.140; in group 2 were 2.3%, 21.2%, and 76.5%, and 0.130, respectively. |
| 47. Verdi H et al. 2020 [51] | Case-Control | Genotyping analysis by Real time PCR, melting curve; 3826A/G UCP1 SNP | Turkish (n=189)  **Group 1:** obese children n=102 (f=54, f=48), age=12.3±2.8 years, BMI z score=2.6±0.5  **Group 2:** control n=87 (f=48, m=39), age= 11.9±3.2 years, BMI z score=-0.7±0.8 | 1. A-3826G allele frequencies in obese group G allele=27% and 22% in control group 2. A-3826G genotype frequencies in obese group AA=53%, AG=39%, GG=8%. 3. In control group AA=66%, AG=25% and GG=9%. 4. UCP1 A-3826G genotype is not associated with obesity, metabolic disorders, gender and glucose- insulin responses during oral glucose tolerance test. 11. |
| 48. Vimaleswaran et al, 2010 [52] | Case-Control | Genotyping analysis by PCR- fast real system; A-3826G and A-112C UCP1 SNPs. | Asian Indians (n=1800)  **Group 1:** normal glucose tolerant (n=990); m=374, f=616; age=49±12 years; BMI=24±4.7 kg/m^2^.  **Group 2:** T2DM (n=810); m=353, f=457; age=43±13 years; BMI=26.1±4.2 kg/m^2^. | 1. A-3826G genotype frequencies in group 1 were for AA 40%, for AG, 45%, and for GG 15%; in group 2 were 36%, 46%, and 18%, respectively (p=0.11). 2. -3826G allele frequency for groups1 and 2 was 0.38 and o.41, respectively (p=0.11). 3. A-112C genotype frequencies in group 1 were for AA 62%, for AC 34%, and for CC 4%; in group 2 were 63%, 33%, and 4%, respectively (p=0.87). 4. -112C allele frequency for groups 1 and 2 was 0.21 (p=0.87).   A-3826G and A-112C UCP1 genotype and allele frequencies were not associated with T2DM. |
| 49. Vimaleswaran et al, 2007 [53] |  | Genotyping analysis by PCR- fast real system; A-3826G and A-112C UCP1 SNPs. | Asian Indians (n=1500)  **Group 1:** normal glucose tolerant (n=950); Subdivided into:  Group 1a: metabolic syndrome (n=211); m=78, f=133; age=43±11 years; BMI=27.1±4.2 kg/m^2^.  Group 1b: no metabolic syndrome (n= 739); m=292, f=447; age=37±12 years; BMI=22.4±4.3 kg/m^2^.  **Group 2:** T2DM (n= 550); Subdivided into:  Group 2a: metabolic syndrome (n=402); m=179, f=223; age=51±11 years; BMI=25.8±4.2 kg/m^2^.  Group 2b: no metabolic syndrome (n= 148); m=70, f=78; age=51±12 years; BMI=23.6±3 kg/m^2^. | 1. A-3826G genotype frequencies based in no metabolic syndrome groups (n=887) and in metabolic syndrome groups (n=613) were for AA 58% and 56%, for AG 36% and 39%, and for GG 6% and 5%, respectively. 2. -3826G allele frequency in no metabolic syndrome and metabolic syndrome groups was 0.24, respectively. 3. A-112C genotype frequencies in no metabolic syndrome and metabolic syndrome groups were for AA 74% and 70%, for AC 24% and 28%, and for CC 2%, respectively. 4. -112C allele frequency in no metabolic syndrome and metabolic syndrome groups was 0.14 and 0.16, respectively. 5. A-3826G allelic (p=0.89) and genotypic (p=0.26) were not associated with metabolic syndrome. 5. A-112C allelic (p=0.16) and genotypic (p=0.21) distributions were not associated with metabolic syndrome. |
| 50. Yiew et al 2010 [54] | Case only | Genotyping analysis by PCR-RFLPs, A-3826G UCP1 SNP. | Malaysian Chinese (n=256) healthy and unrelated students, age=21.7 ± 1.7 years | 1. G allele frequency = 0.58 2. In lean subjects: AA=10%, AG=61, 8%, GG=28,2% 3. In overweight subjects: AA=12.8%, AG=60.5%, GG=26.7% 4. UCP1 −3826A/G SNP is not associated with obesity and its related anthropometric indicators among the Malaysian Chinese university students |
| 51. Zhang et al, 2015 [55] | Case-Control | Genotyping analysis by PCR-ligase detection reactions; A-3826G UCP1 SNP. | Chinese (n=792)  **Group 1:** diabetic retinopathy (n=448); m=196, f=252; age=62.35±11.92 years; BMI=25.58±4.18 kg/m^2^.  Subdivided into:  Group 1a**:** diabetic retinopathy proliferative (n= 220); m= 91, f= 119; age=60.36±11.66 years; BMI=27.33±4.06 kg/m^2^.  Group 1b: diabetic retinopathy non-proliferative (n= 228); m=95, f=133; age=63.03±11.57 years; BMI=25.20±4.13 kg/m^2^.  **Group 2:** diabetic retinopathy proliferative -no signs of diabetic retinopathy (n=334); m=163, f= 181; age=60.16±11.67 years; BMI=26.16±4.75 kg/m^2^. | 1. A-3826G genotype frequencies in diabetic retinopathy were for AA 23.6%, AG 48.9%, GG 27.5%; in diabetic non-retinopathy were 28.1%, 48.2%, 3.7%, respectively; in diabetic retinopathy proliferative were 20.7%, 49.3%, 30%, respectively; in diabetic retinopathy non-proliferative 26.4%, 48.5%, 25.1%, respectively. 2. -3826G allele frequency in diabetic retinopathy, diabetic retinopathy proliferative, proliferative diabetic and diabetic retinopathy non-proliferative was 51.9%, 47.8%, 54.6%, and 49.3%, respectively. 3. The frequency of the -3826GG genotype was higher in the diabetic retinopathy proliferative than in the diabetic retinopathy non-proliferative group in the additive model (OR=1.72, 95%CI=1.06–2.79, p=0.03). 4. The frequency of the -3826G allele in the additive model was higher in the diabetic retinopathy proliferative than in the diabetic retinopathy non-proliferative (OR=1.32, 95% CI=1.03–1.68; p=0.03). 5. No differences were found for the A-3826G allele frequencies and genotype distributions between the diabetic retinopathy and diabetic retinopathy proliferative or diabetic retinopathy non-proliferative and diabetic proliferative (p>0.05). 6. The A-3826G polymorphism is associated with increased risk of diabetic retinopathy proliferative in T2DM individuals. |
| 52. Zietz et al, 2001 [56] | Case-only | Genotyping analysis by PCR- fast real system; A-3826G UCP1 SNP. | Germans (n=549)  T2DM; m=312, f=237. | 1. A-3826G genotype frequencies were for AA 58.3%, for AG 37.3%, and for GG 4.4%. 2. -3826G allele frequency 0.23. 3. No differences in grade of retinopathy were found among A-3826G genotypes. 4. Serum levels of dehydroepiandrosterone sulfate were lowest in -3826GG homozygotes with no retinopathy compared with those carriers of the -3826AG and AA genotypes (p<0.05). 5. Among female T2DM, dehydroepiandrosterone sulfate was negatively correlated to cholesterol and positively to SBP (p<0.05). 6. No differences in sex, age, BMI known duration of diabetes, cholesterol, glycemic control (HbA1c), SBP, serum levels of C-peptide, cortisol and leptin between A-3826G genotypes were found. |

**Key:** PCR= polymerase chain reaction; UCP1= uncoupling protein one; SNP= single nucleotide polymorphism; m= male; f=female; BMI=body mass index; IR= insulin resistance; OR=odds ratio; T2DM= type 2 diabetes mellitus; SBP= systolic blood pressure; DBP= diastolic blood pressure; TC= total cholesterol; HDL-C= high density lipoprotein-cholesterolemia; LDL-C= low density lipoprotein-cholesterolemia; HbA1c= glycated haemoglobin; FPG= fasting plasma glucose; TG= triglycerides; WHR= waist-to-hip ratio; HOMA-IR= homeostatic Model Assessment of Insulin Resistance; OLTT= oral lipid tolerance test; OGTT= oral glucose tolerance test; CIN= cerebral infarction; DP= Dampness-phlegm; CI= confidence interval; FFM= free fat mass.

**2.2.1** The results from the SNP-specific forest and funnel plots for the prevalence (Figures S7-26) and the odds ratio (Figures S27-36) for different genotypes are shown below. Funnel plots were only produced for those meta-analyses that included >10 studies [5].

**Figure S5:** Forest plot for prevalence of UCP1 A-1766G / AG in the CMP population.

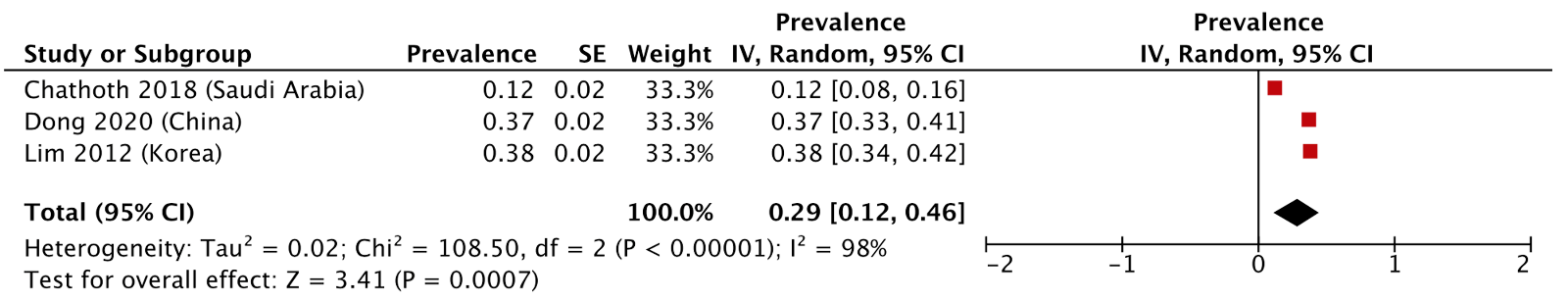

**Figure S6:** Forest plot for prevalence of UCP1 A-1766G / AG in healthy individuals.

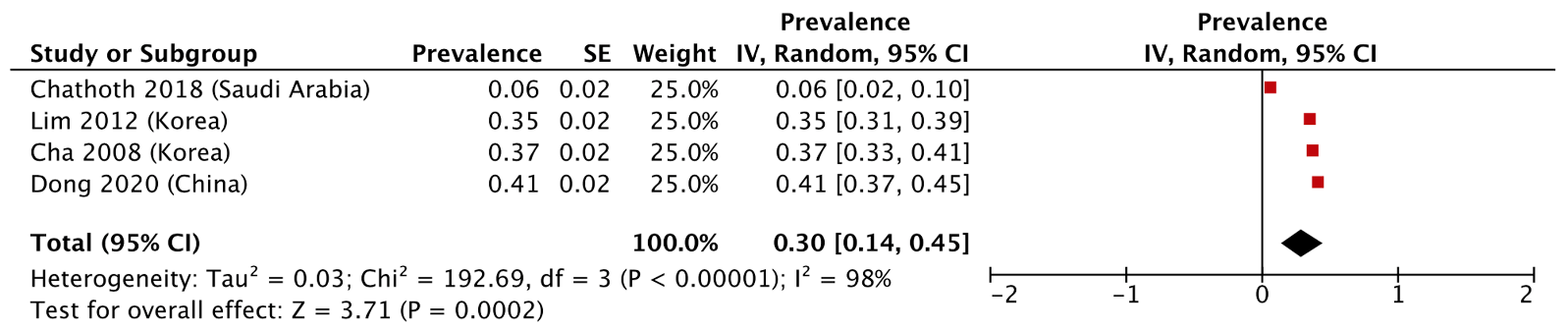

**Figure S7:** Forest plot for prevalence of *UCP1* A-3826G / AG in CMP individuals.

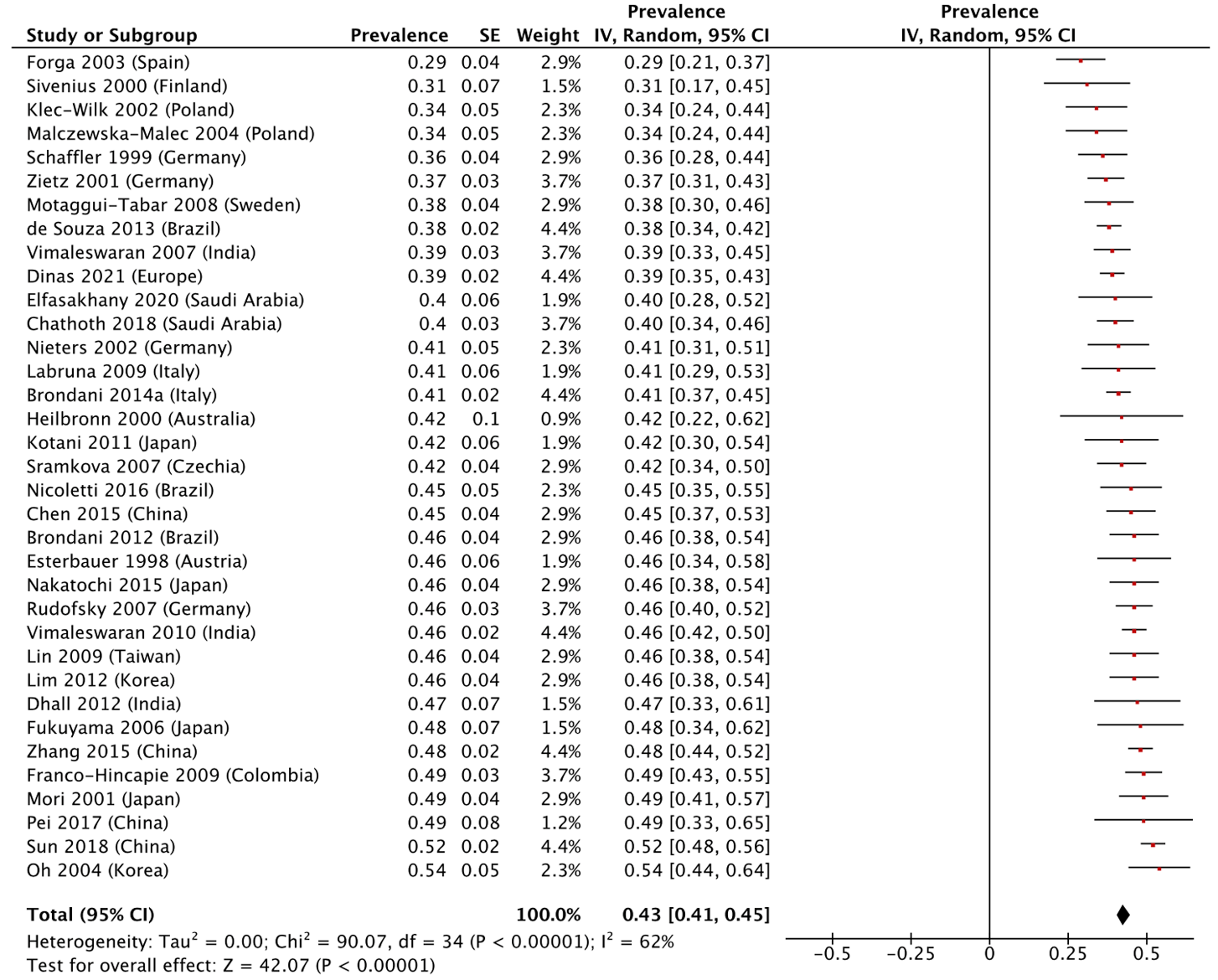
**Figure S8:** Funnel plot for prevalence of UCP1 A-3826G / AG in CMP individuals.

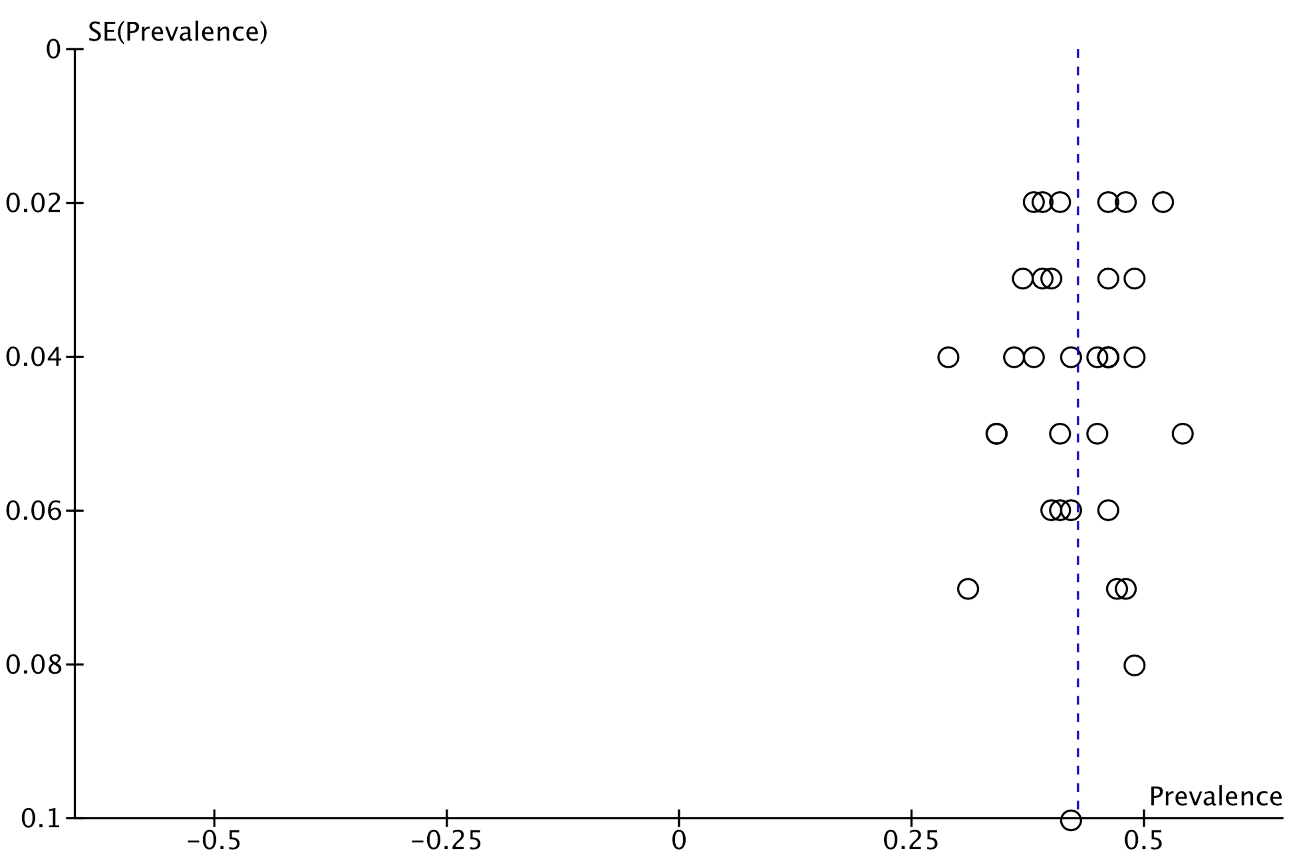

**Figure S9:** Forest plot for prevalence of UCP1 A-3826G / AG in healthy individuals.
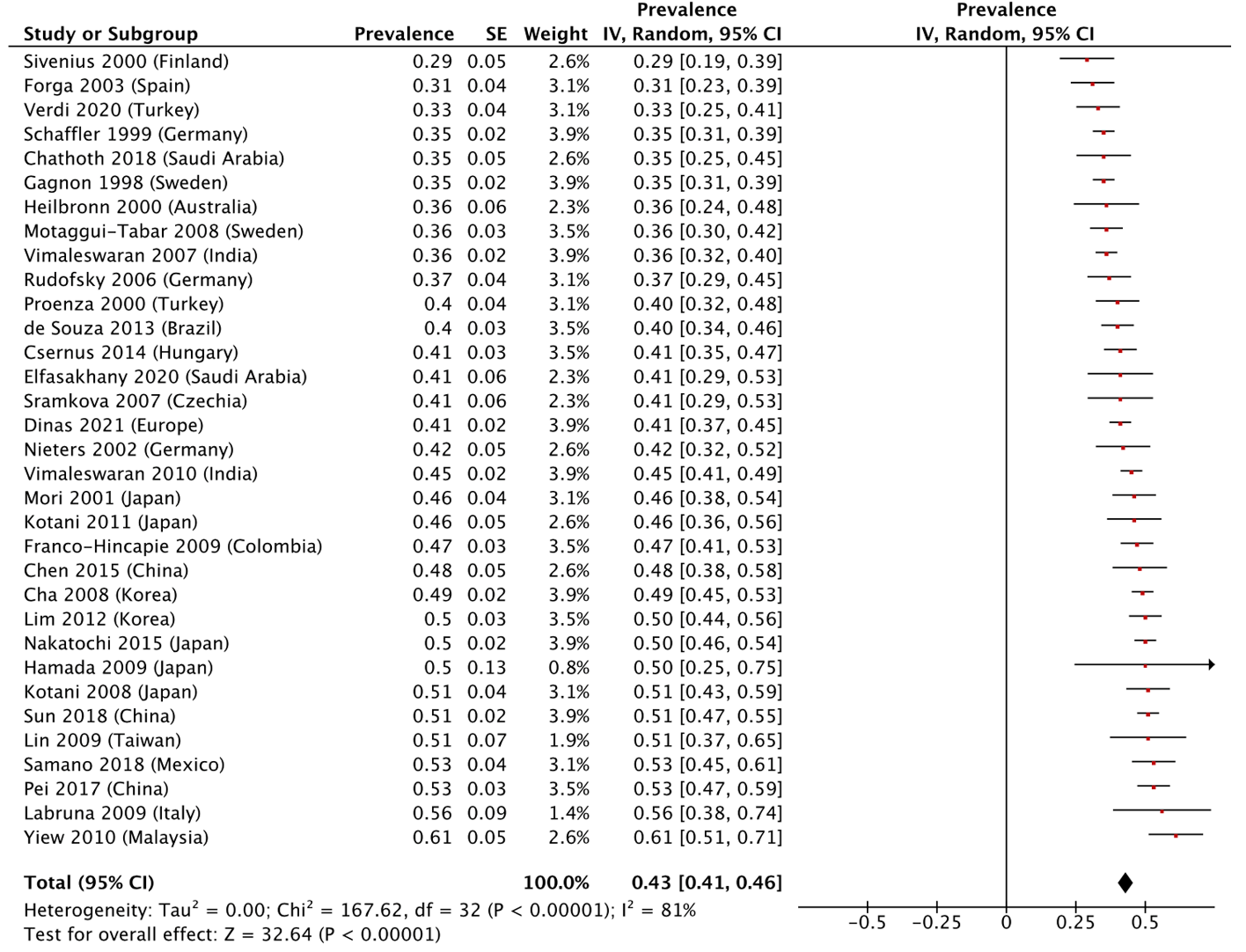

**Figure S10:** Funnel plot for prevalence of UCP1 A-3826G / AG in healthy individual
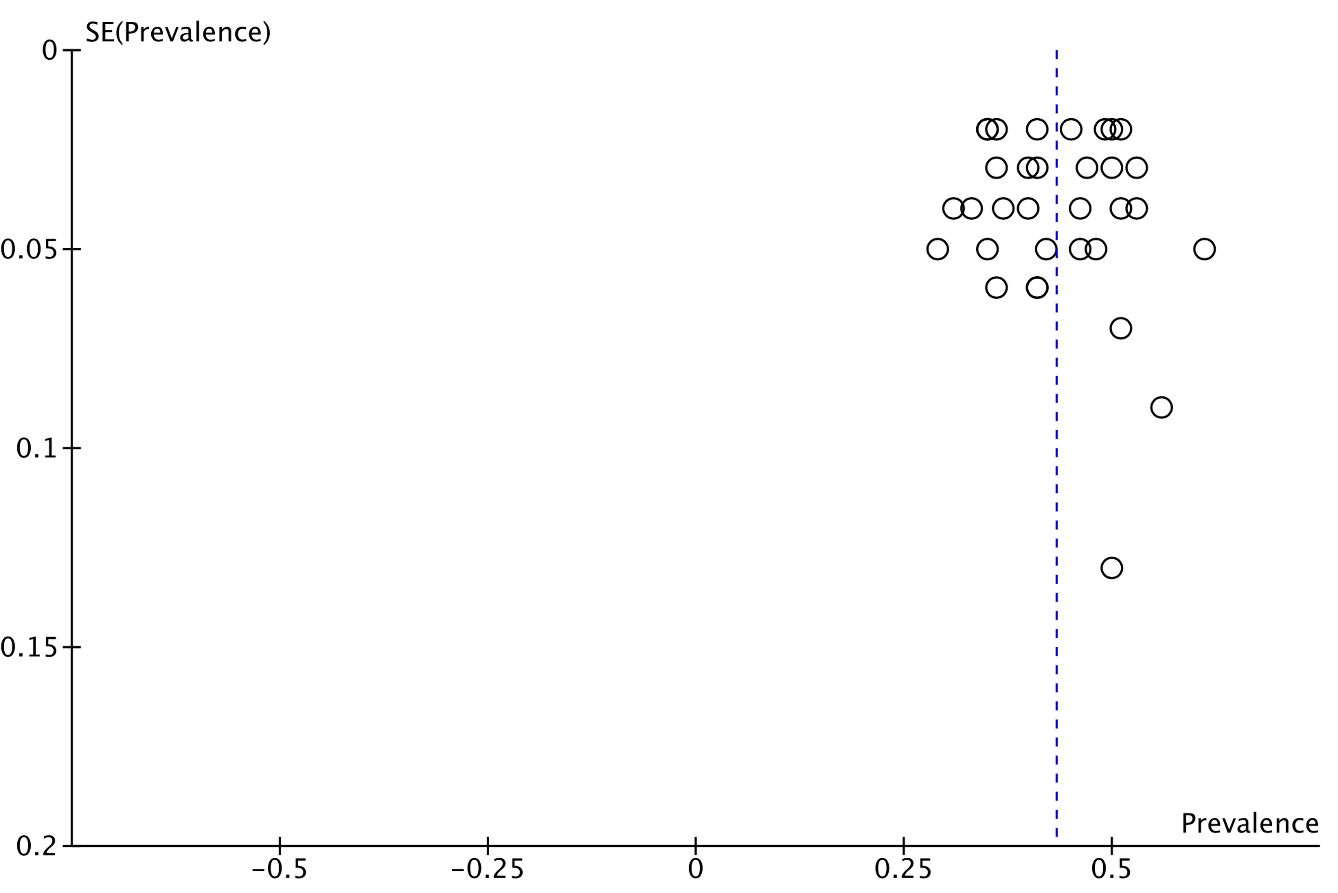

**Figure S11:** Forest plot for prevalence of UCP1 Ala64Thr / GA in CMP individuals
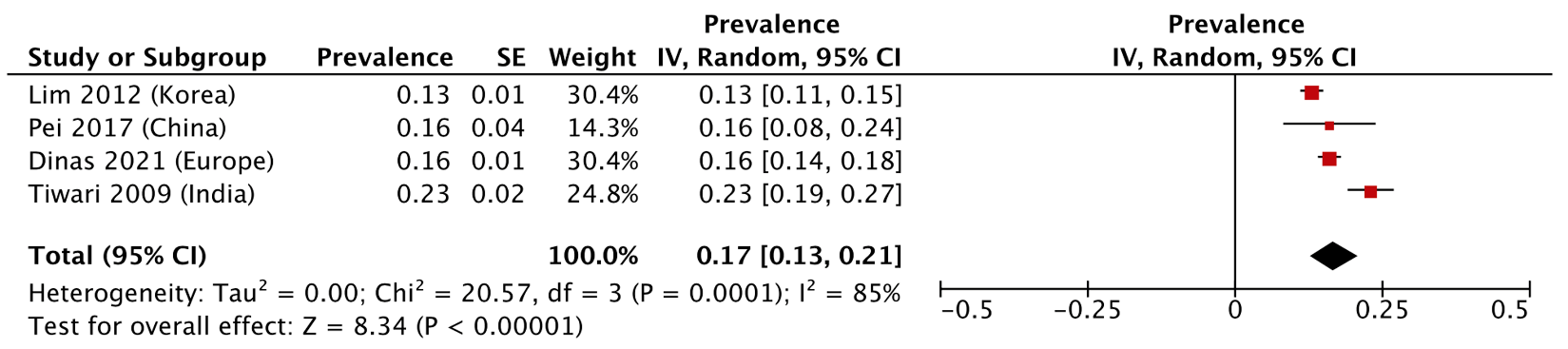

**Figure S12:** Forest plot for prevalence of *UCP1* Ala64Thr / GA in healthy individuals.
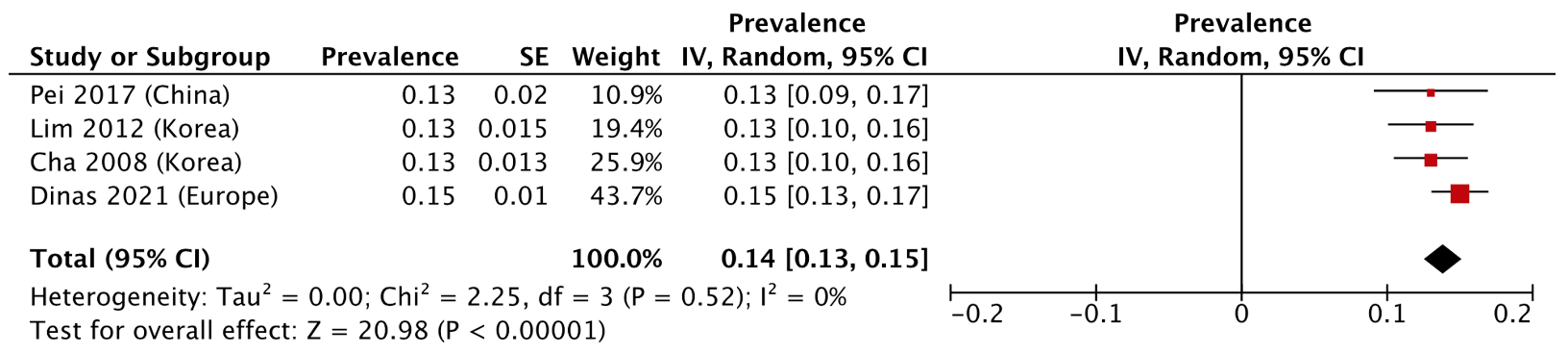

**Figure S13:** Forest plot for prevalence of *UCP1* A-112C / AC in CMP individuals.

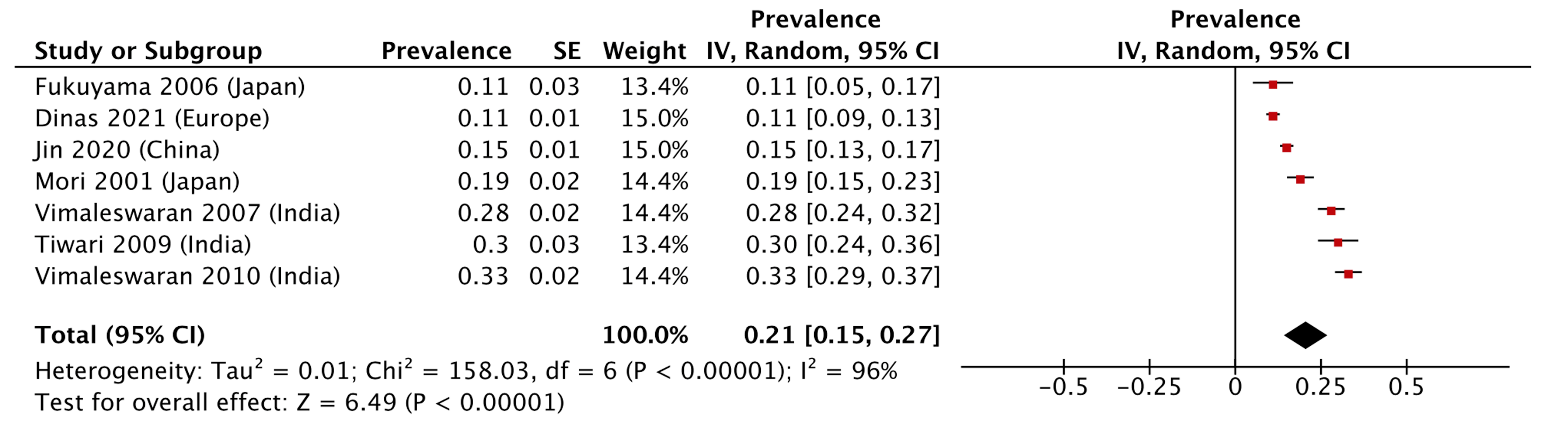

**Figure S14:** Forest plot for prevalence of *UCP1* A-112C / AC in healthy individuals.

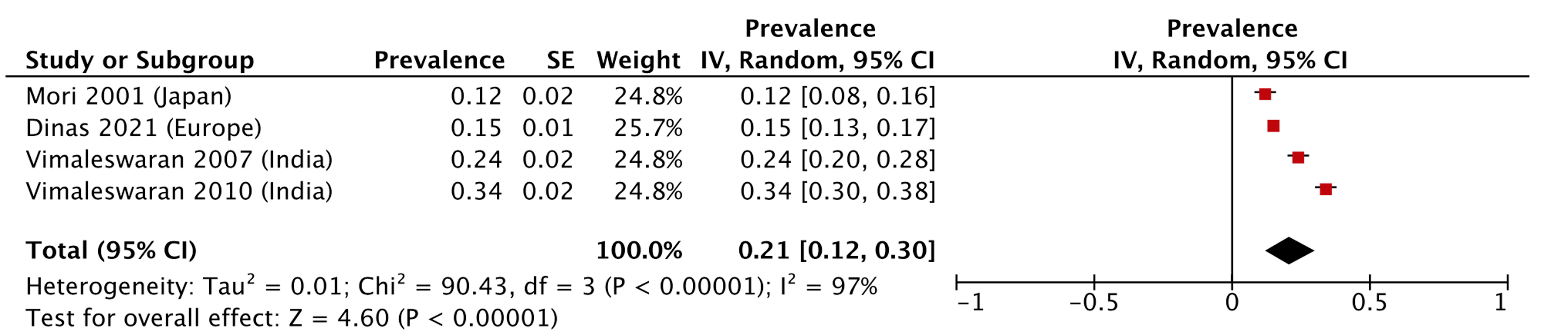

**Figure S15:** Forest plot for prevalence of *UCP1* A-1766G / GG in CMP individuals.

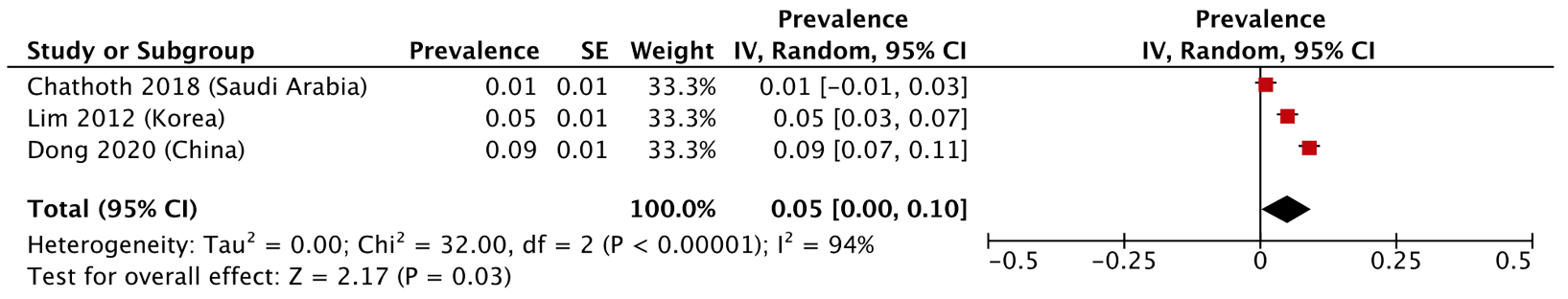

**Figure S16:** Forest plot for prevalence of *UCP1* A-1766G / GG in healthy individuals.

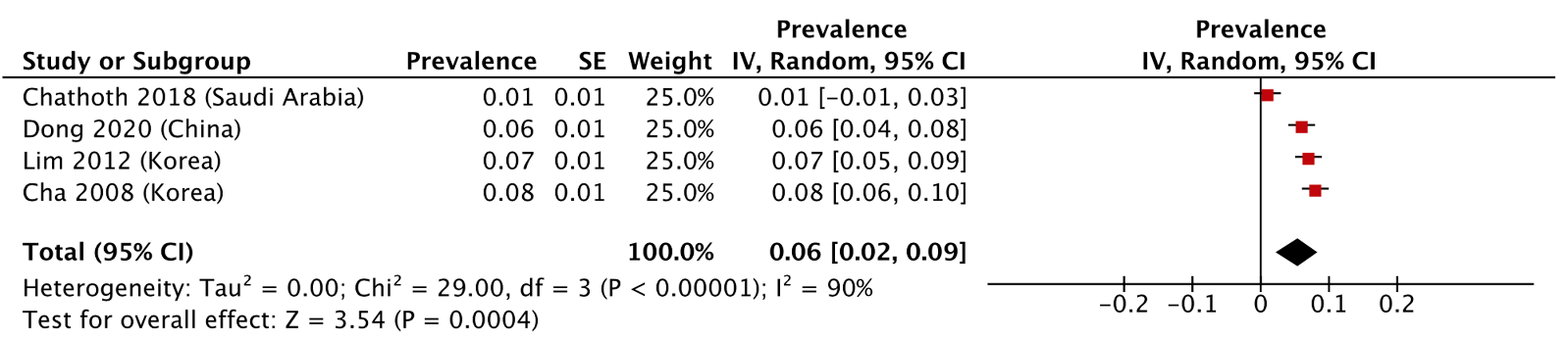
**Figure S17:** Forest plot for prevalence of *UCP1* A-3826G / GG in CMP individuals.

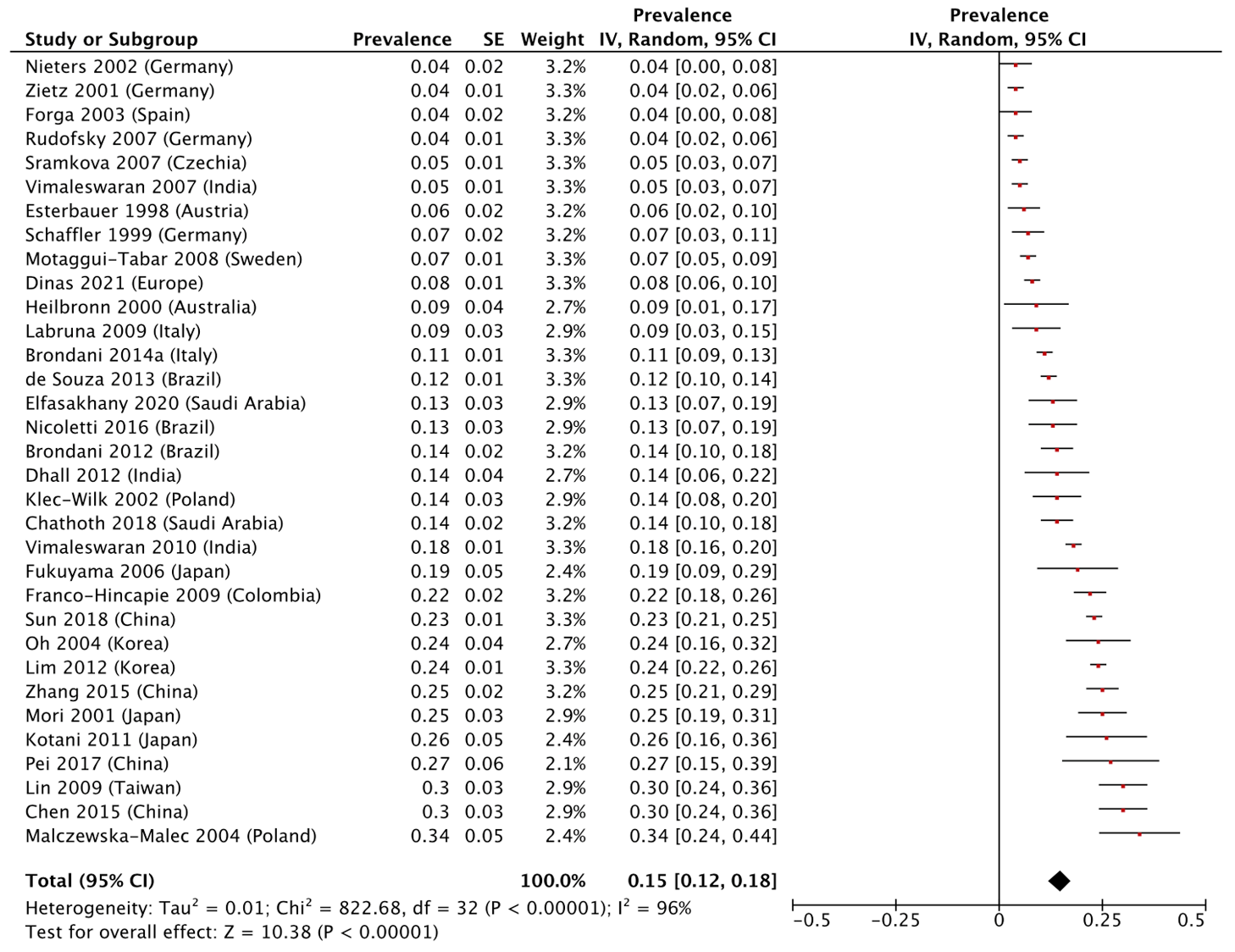

**Figure S18:** Funnel plot for prevalence of *UCP1* A-3826G / GG in CMP individuals.

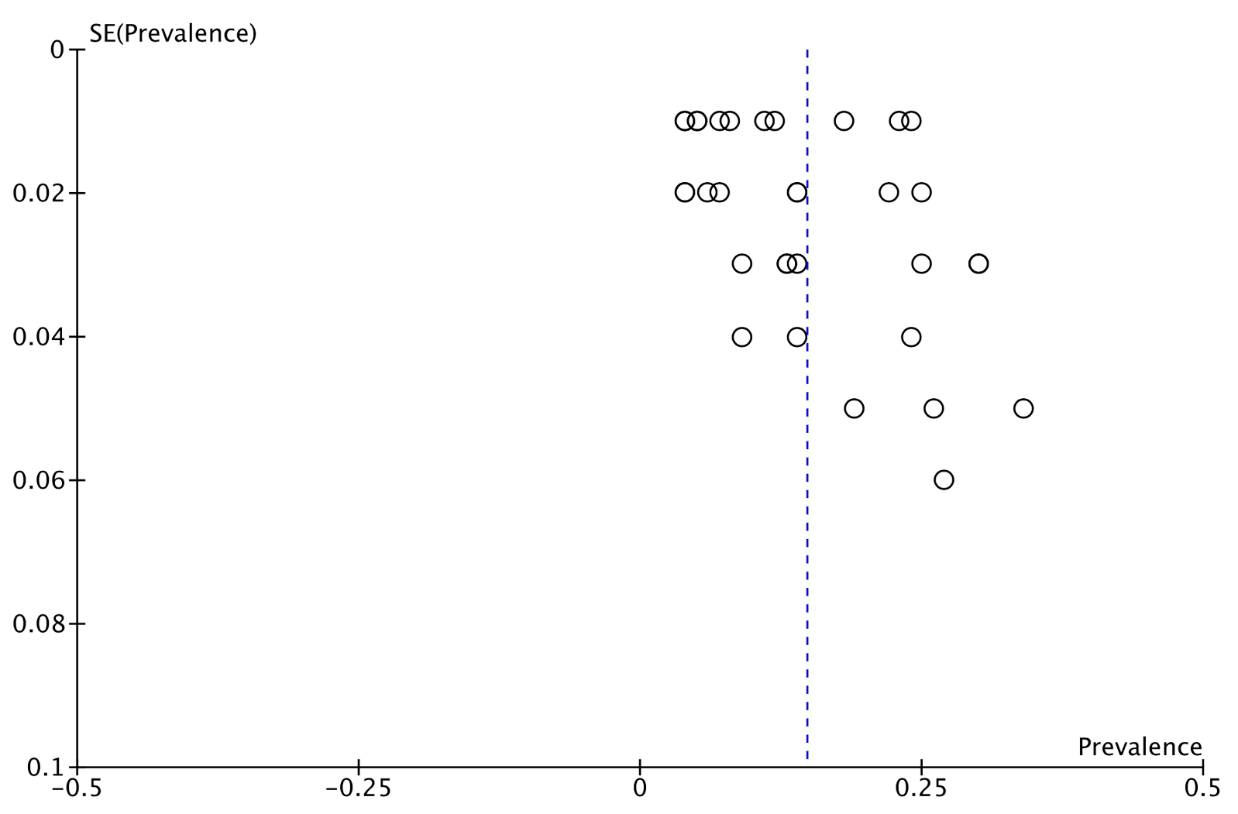

**Figure S19:** Forest plot for prevalence of *UCP1* A-3826G / GG in healthy individuals.

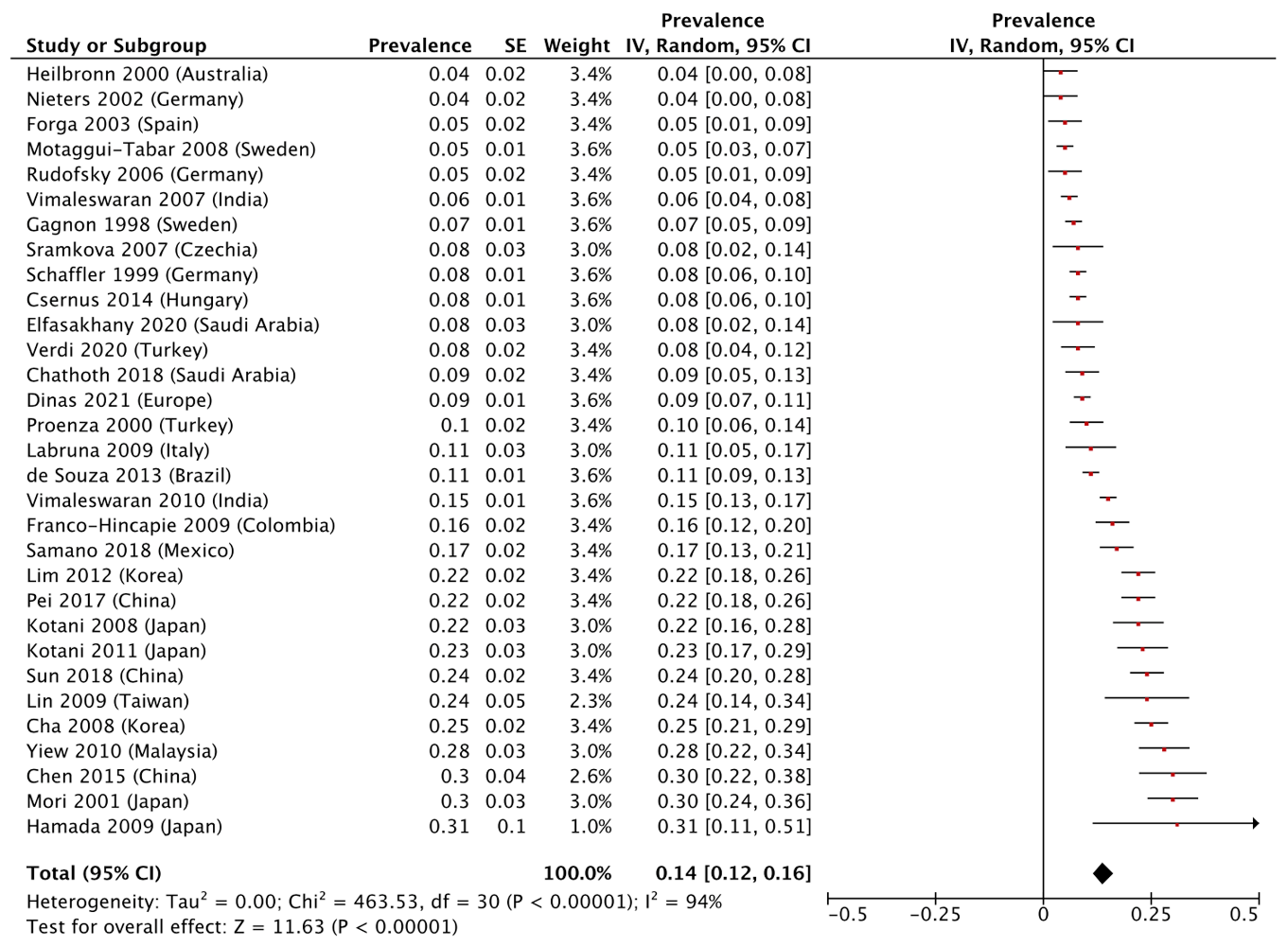

**Figure S20:** Funnel plot for prevalence of *UCP1* A-3826G / GG in healthy individuals.

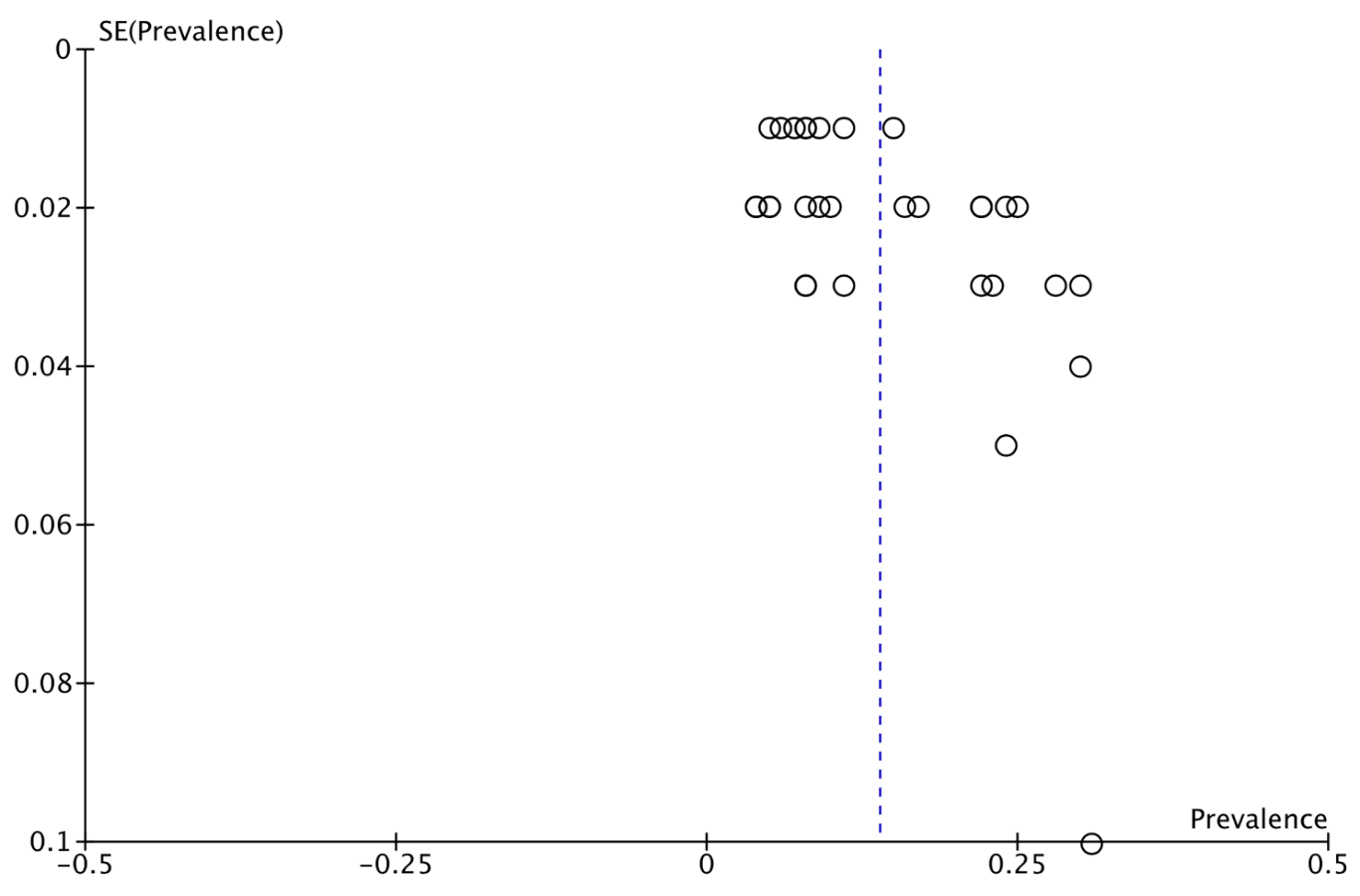

**Figure S21:** Forest plot for prevalence of *UCP1* Ala64Thr / AA in CMP individuals.

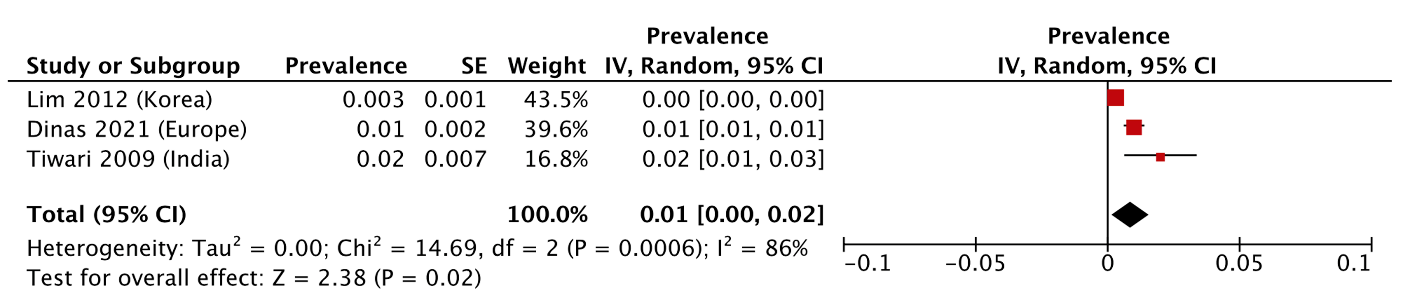

**Figure S22:** Forest plot for prevalence of *UCP1* Ala64Thr / AA in healthy individuals.

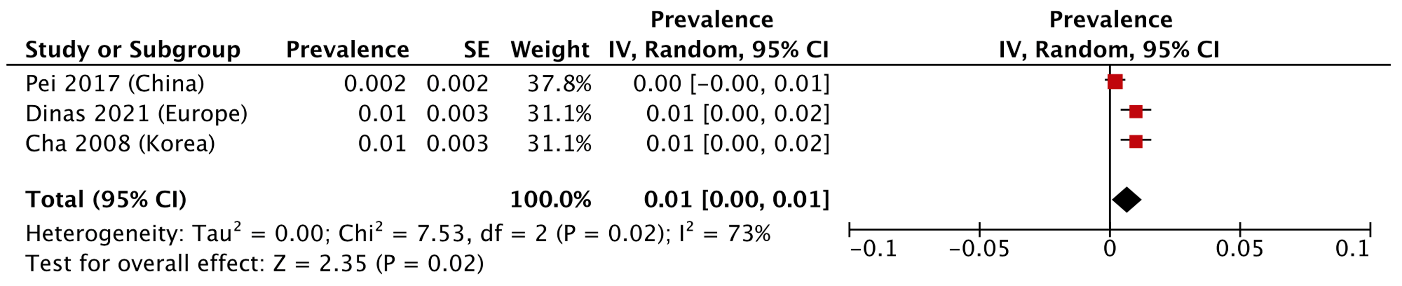

**Figure S23:** Forest plot for prevalence of *UCP1* A-112C / CC in CMP individuals.

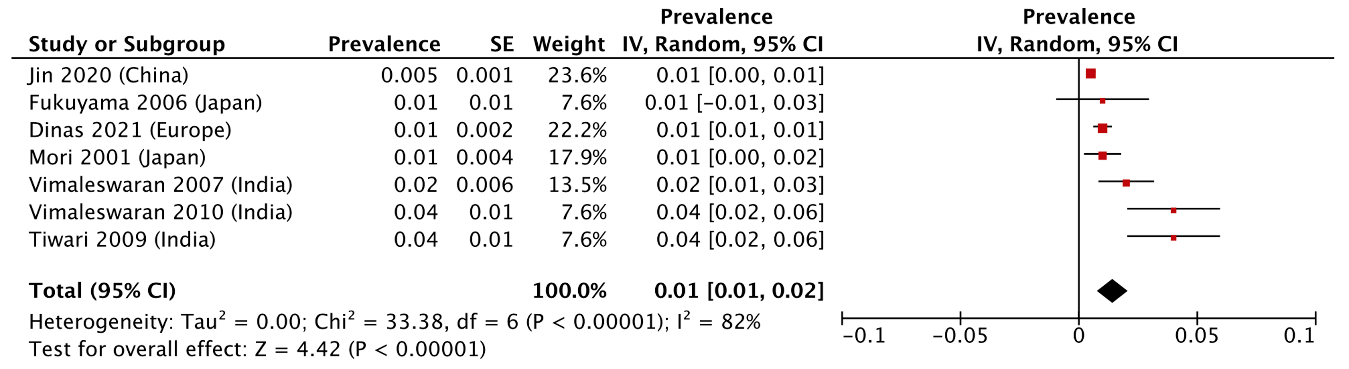

**Figure S24:** Forest plot for prevalence of *UCP1* A-112C / CC in healthy individuals.

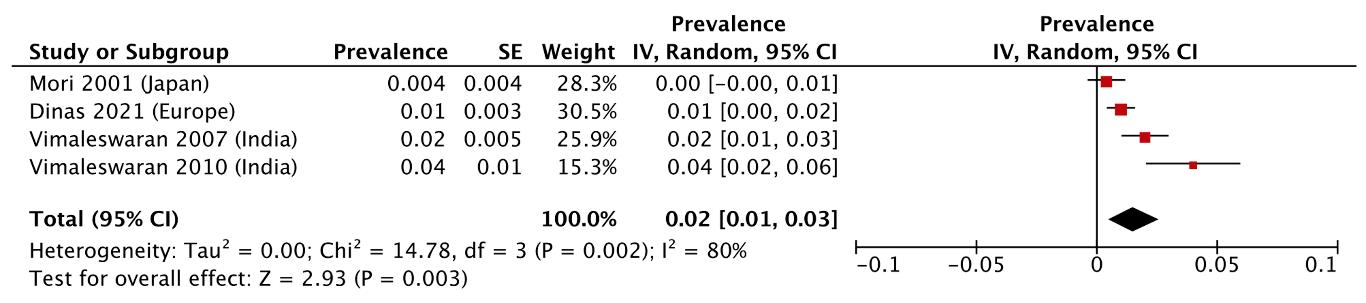

**Figure S25:** Forest plot for odds ratio of *UCP1* A-3826G / AG.

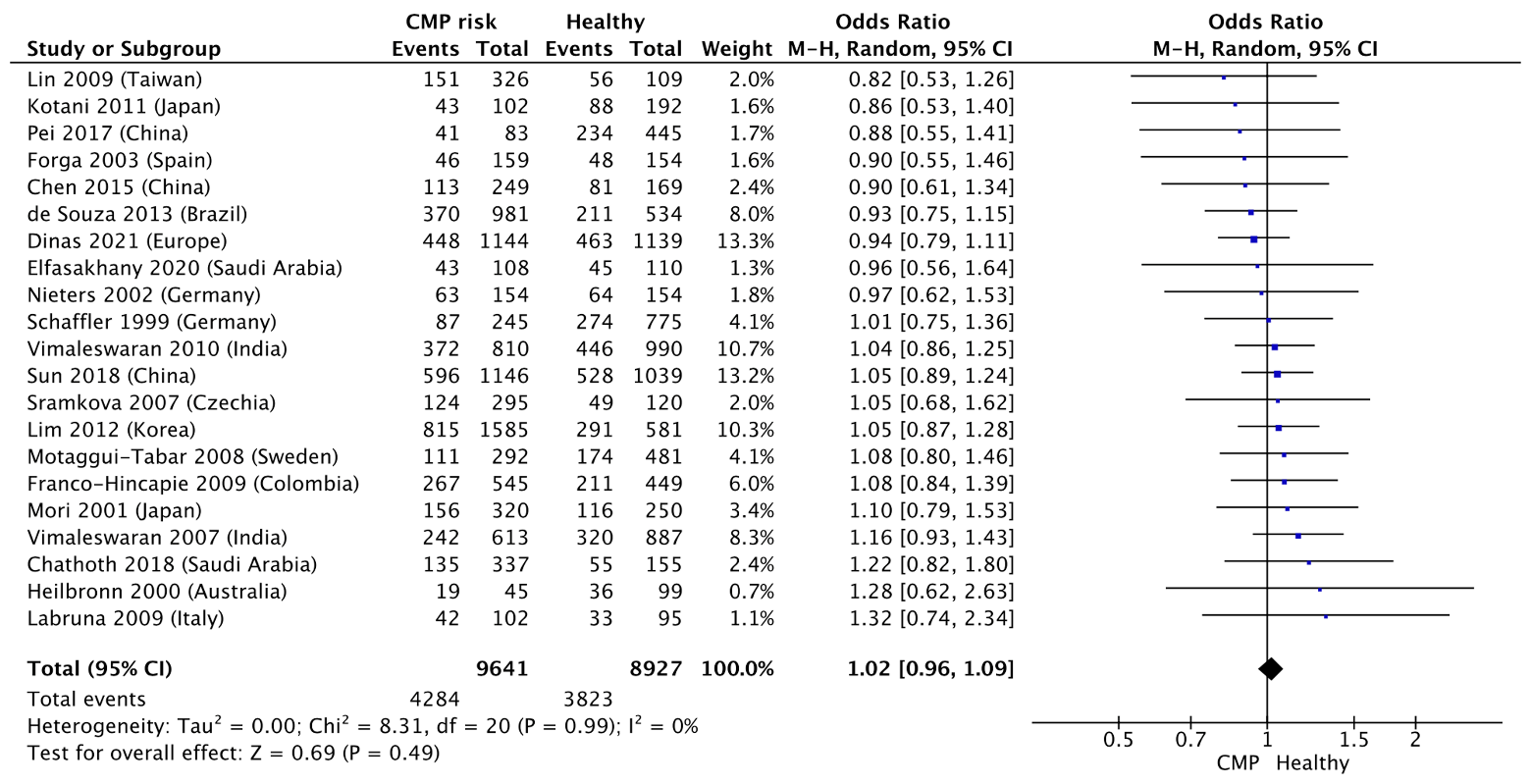

**Figure S26:** Funnel plot for odds ratio of *UCP1* A-3826G / AG.

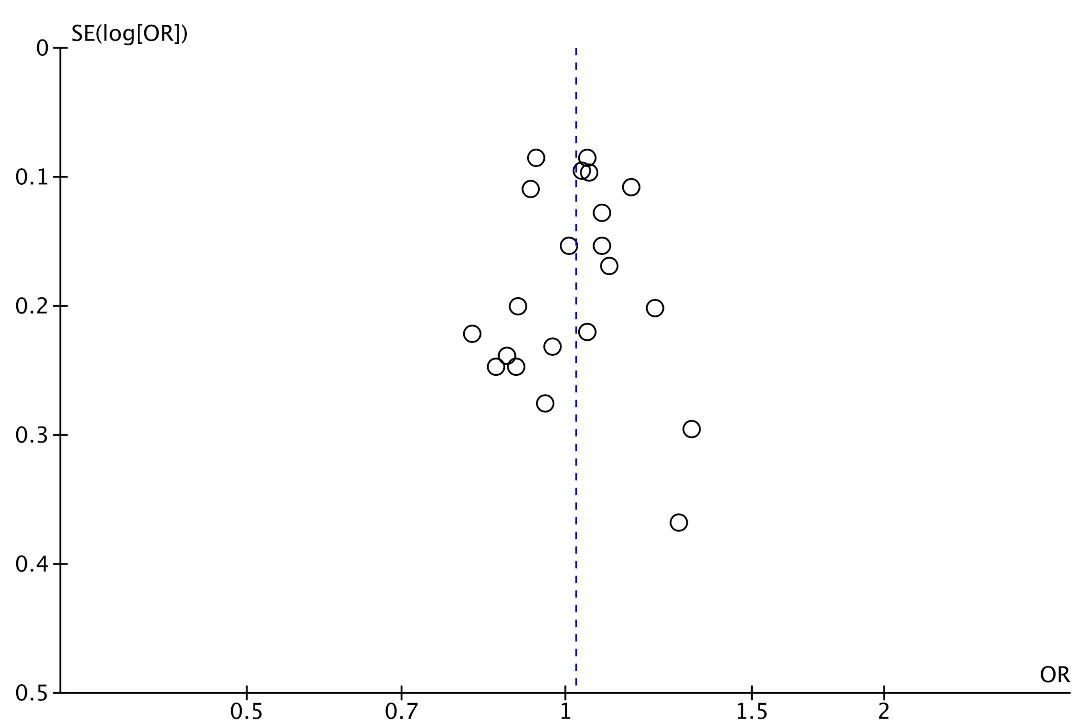

**Figure S27:** Forest plot for odds ratio of *UCP1* A-3826G / GG.

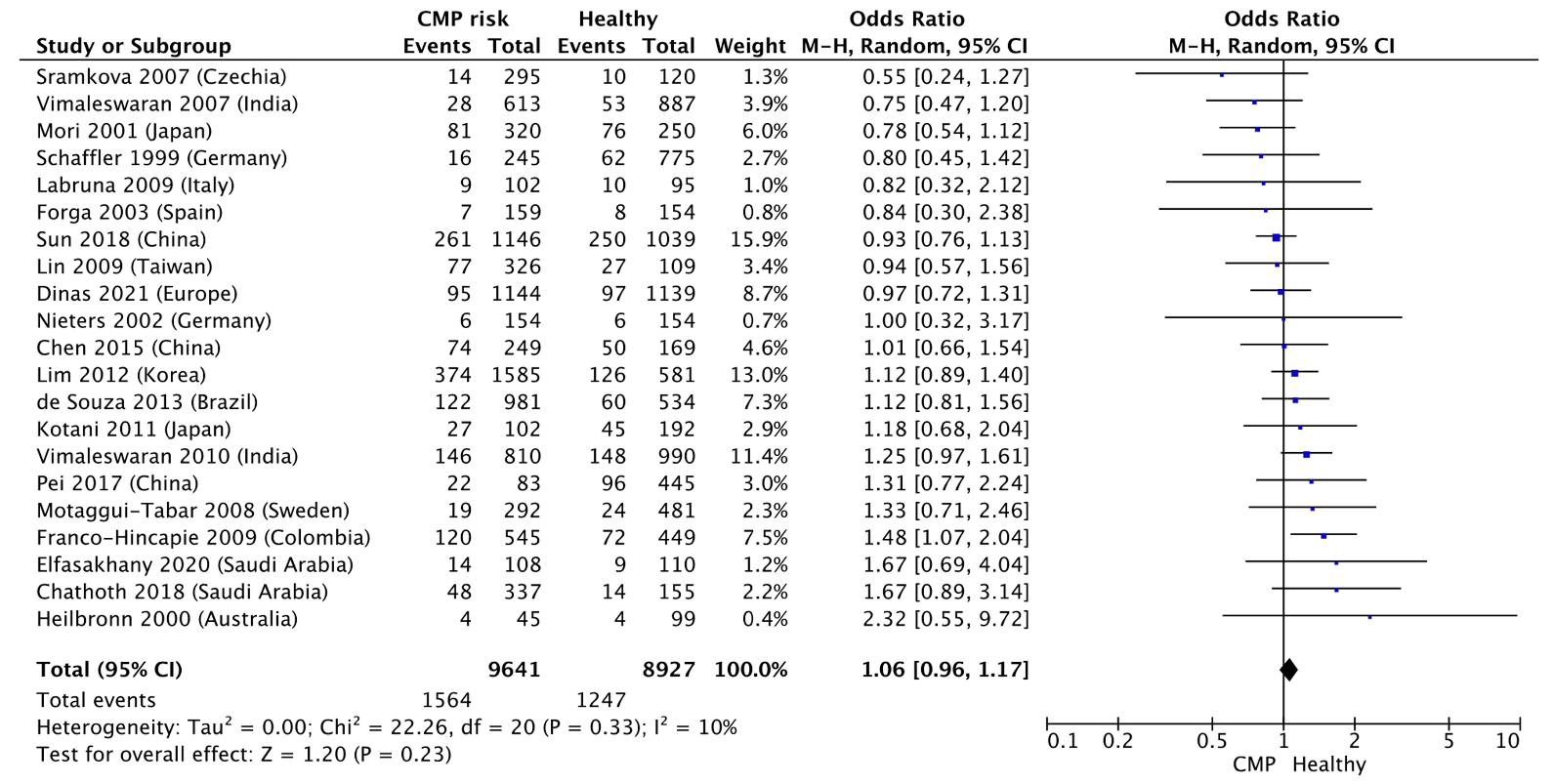

**Figure S28:** Funnel plot for odds ratio of *UCP1* A-3826G / GG.

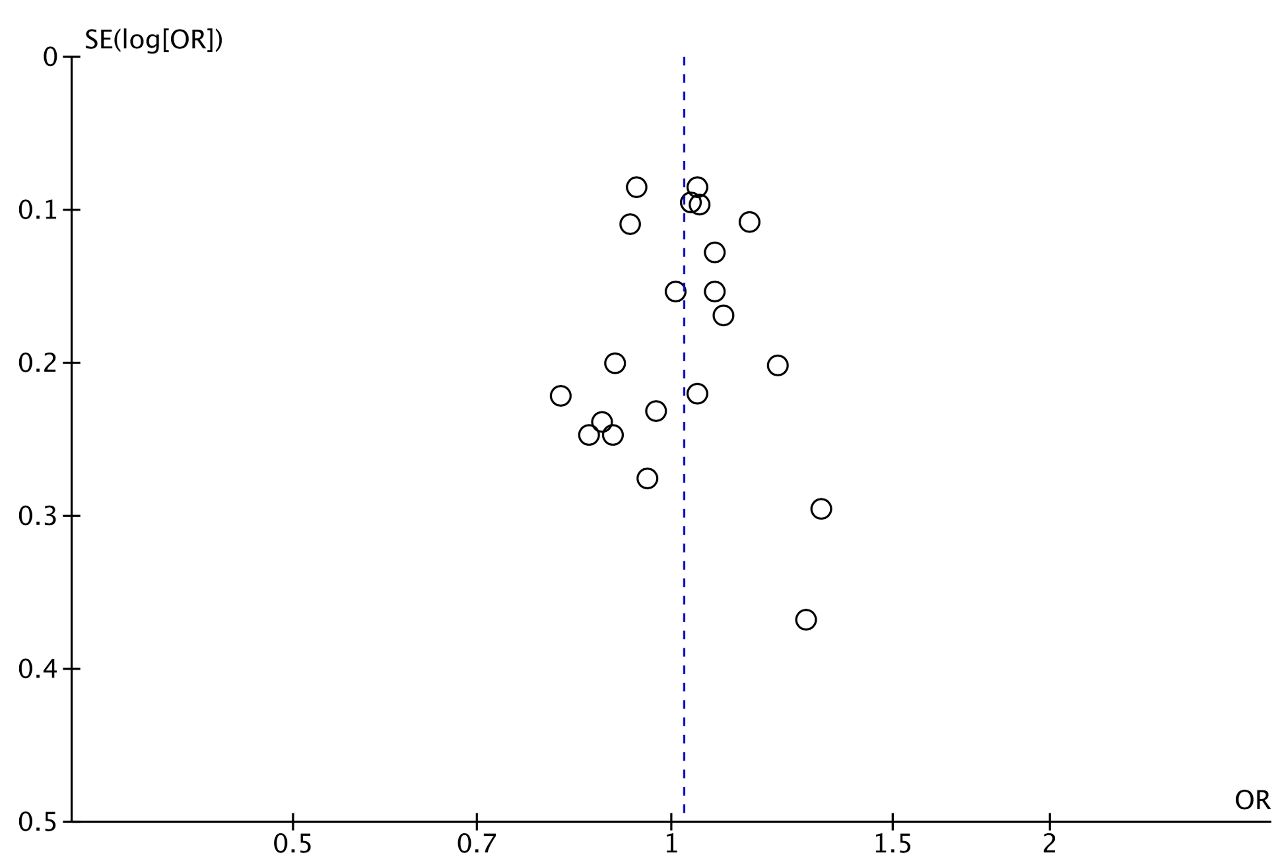
**Figure S29:** Forest plot for odds ratio of *UCP1* Ala64Thr / GA.

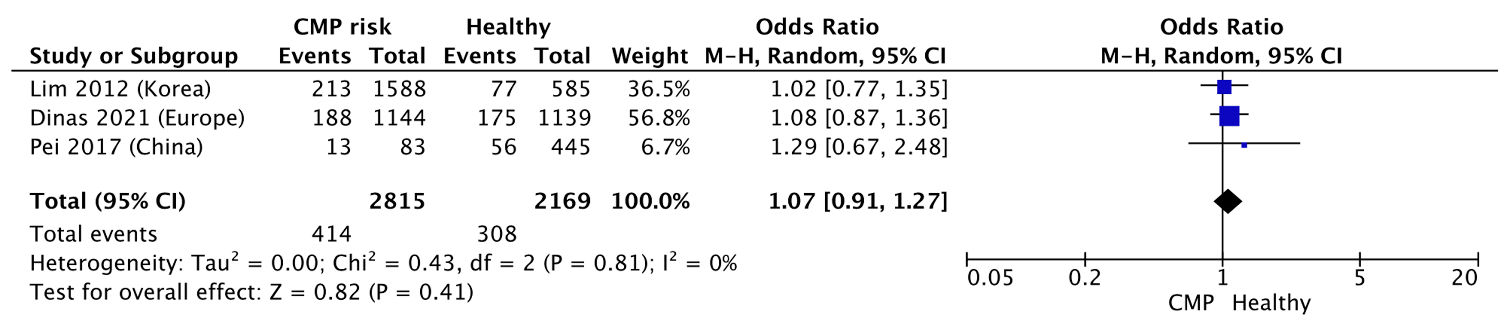

**Figure S30:** Forest plot for odds ratio of *UCP1* Ala64Thr / AA.

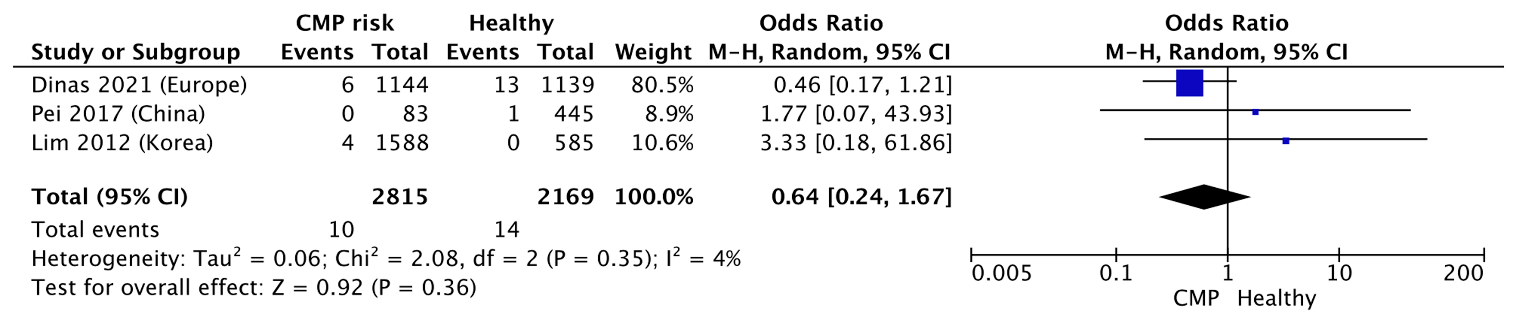

**Figure S31:** Forest plot for odds ratio of *UCP1* A-112C / AC.

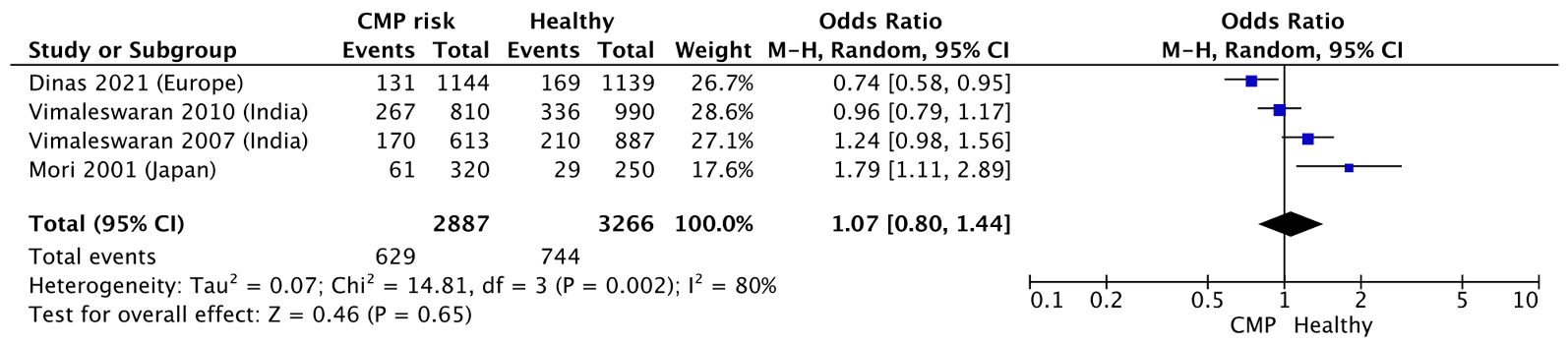

**Figure S32:** Forest plot for odds ratio of *UCP1* A-112C / CC.

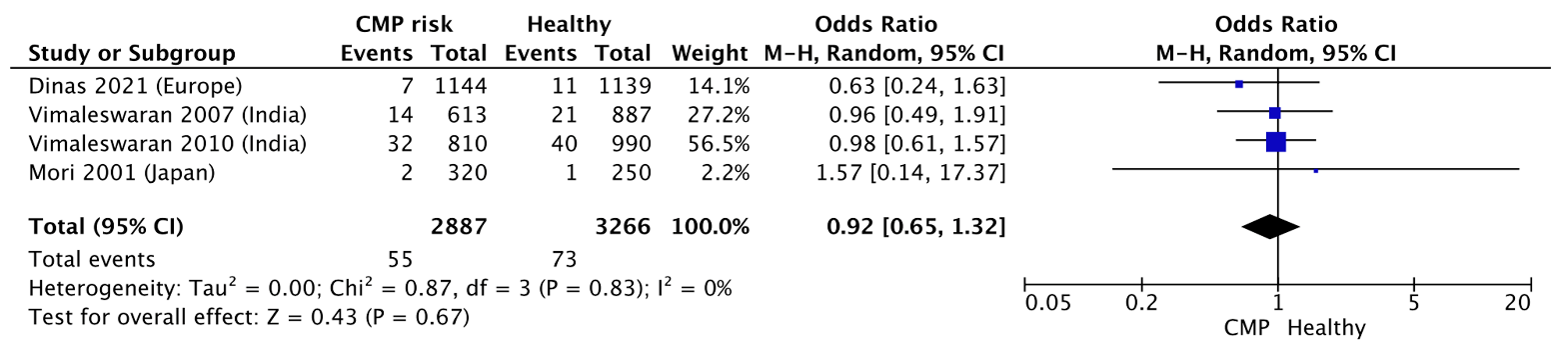

**Figure S33.** Forest plot for the odds ratio of *UCP1* A-1766G /AG.

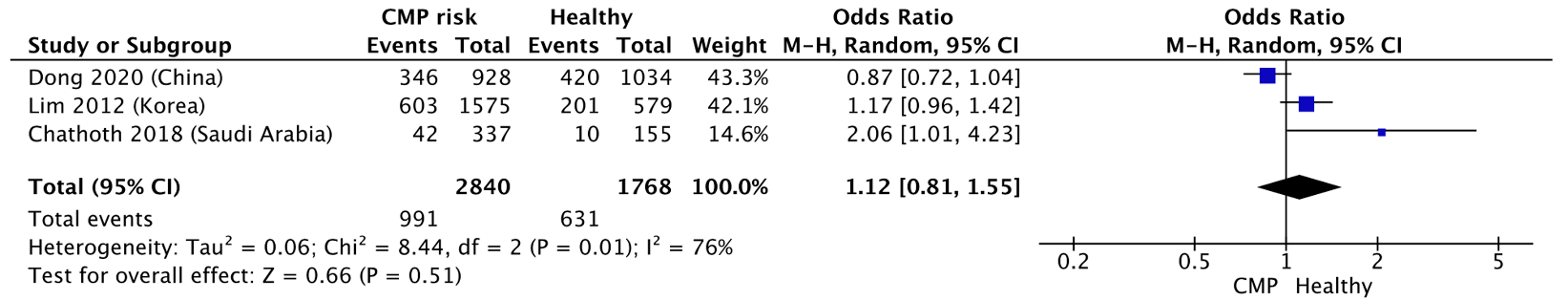

**Figure S34.** Forest plot for the odds ratio of *UCP1* A-1766G /GG.

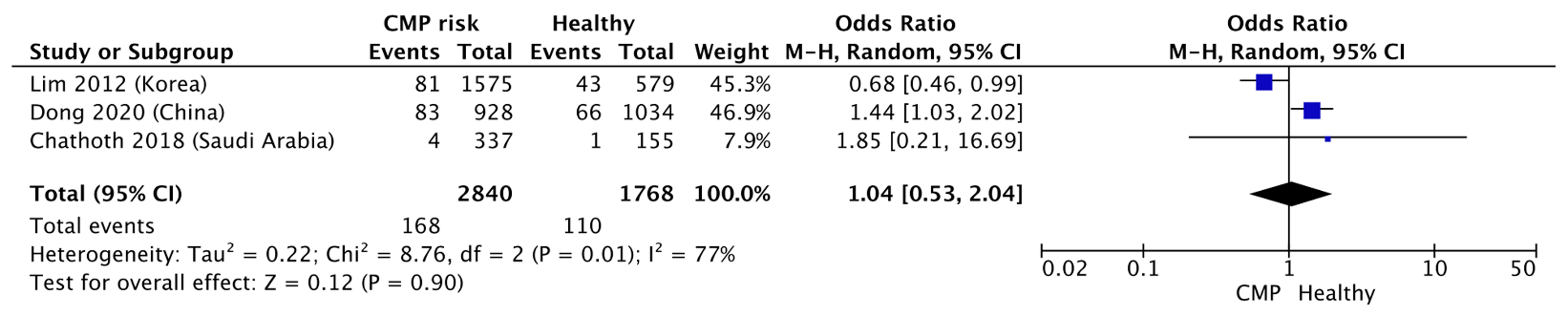

**2.2.2** The results from the allele-specific forest and funnel plots for the prevalence (Figures S37-46) and the odds ratio (Figures S47-51) for different alleles are shown below. Funnel plots were only produced for those meta-analyses that included >10 studies [5].

**Figure S35:** Forest plot for prevalence of *UCP1* A-3826G / G allele in healthy individuals

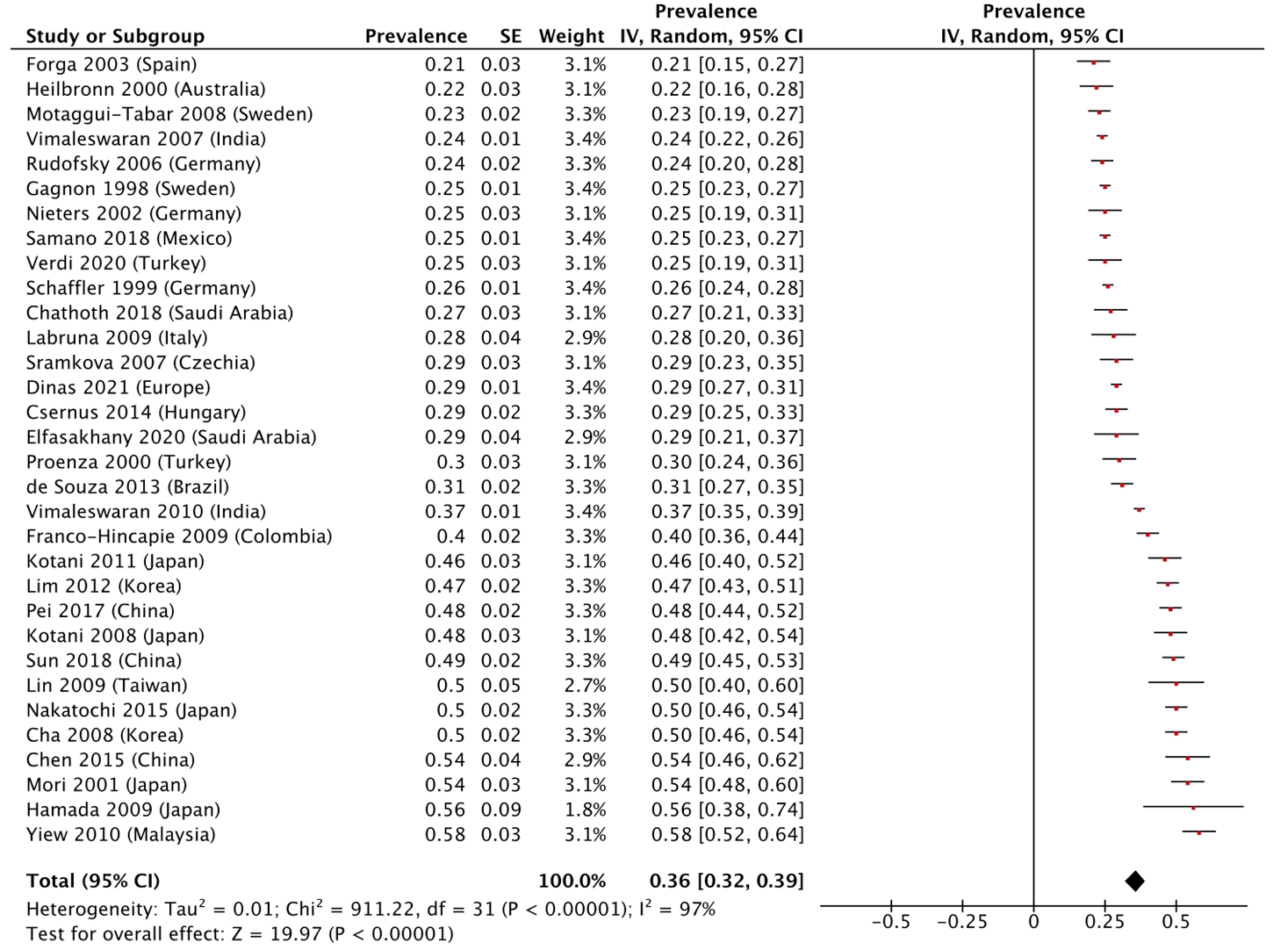

**Figure S36:** Funnel plot for prevalence of *UCP1* A-3826G / G allele in healthy individuals

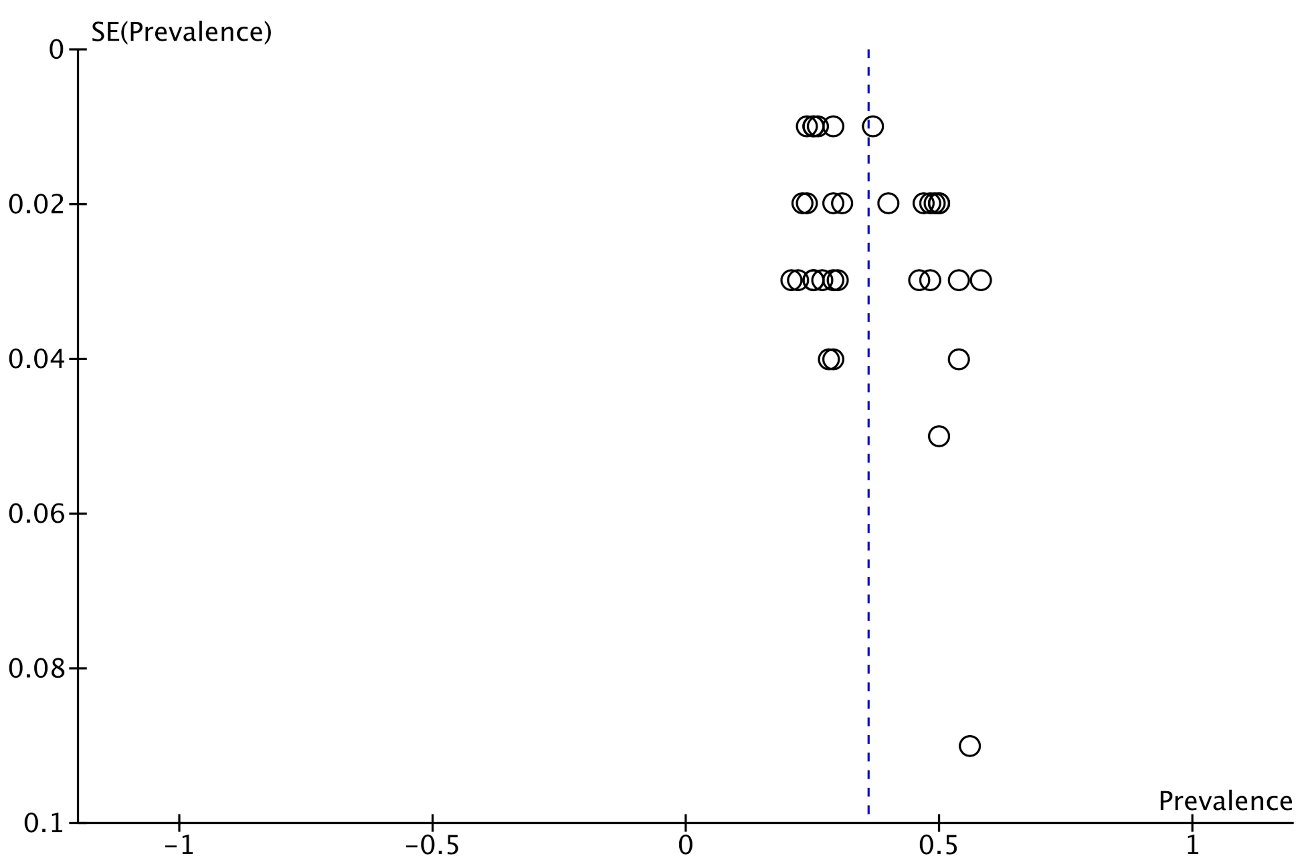

**Figure S37:** Forest plot for prevalence of *UCP1* A-3826G / G allele in CMP individuals.
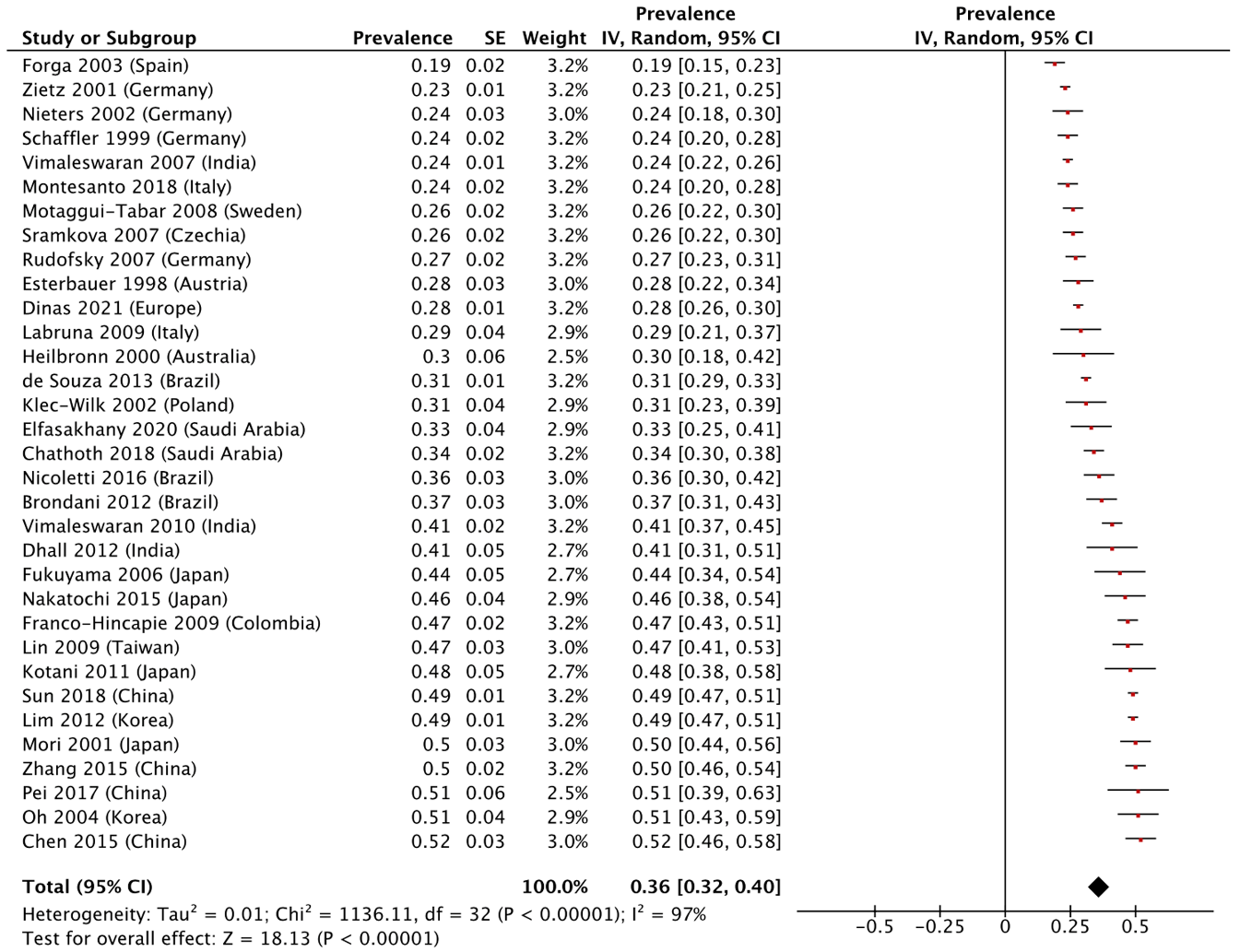

**Figure S38:** Funnel plot for prevalence of *UCP1* A-3826G / G allele in CMP individuals.

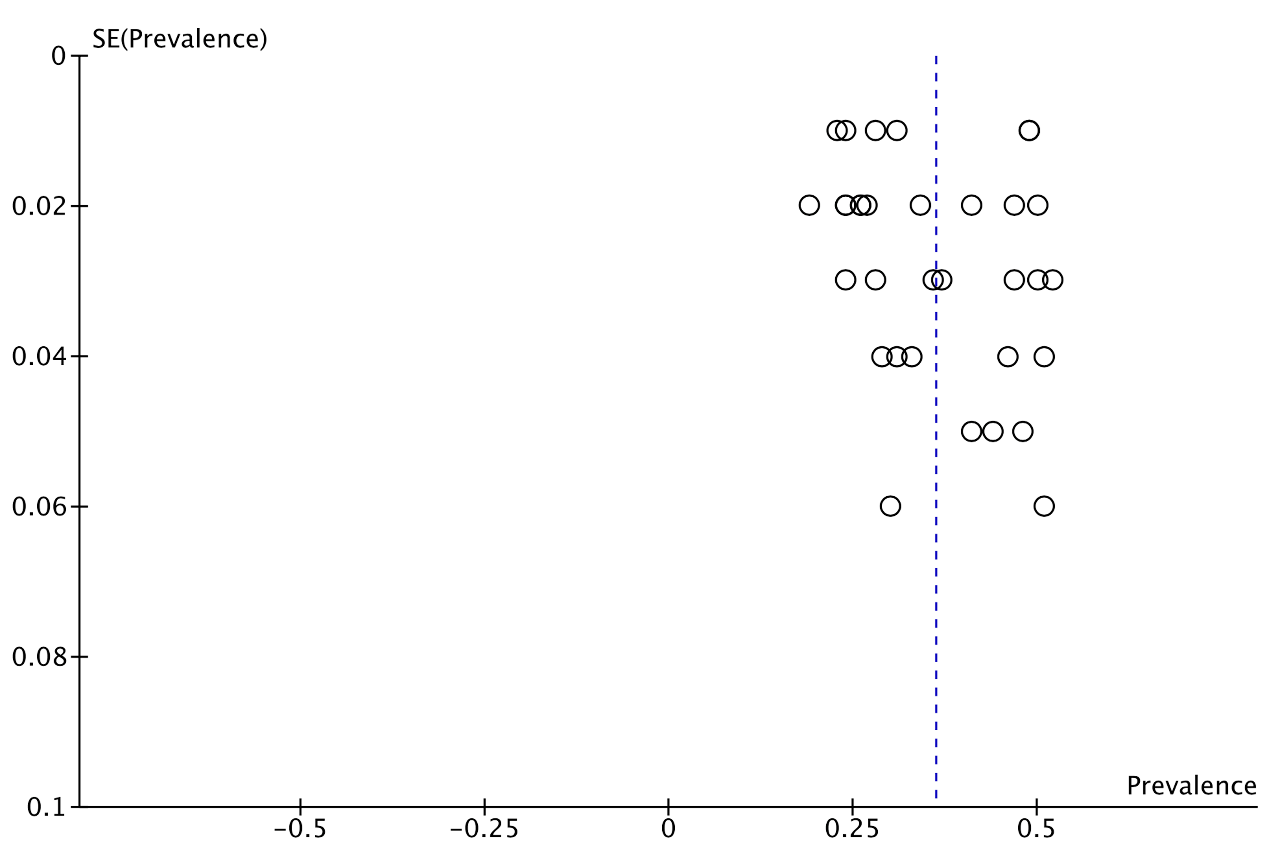

**Figure S39:** Forest plot for prevalence of *UCP1* A-1766G / G allele in healthy individuals.

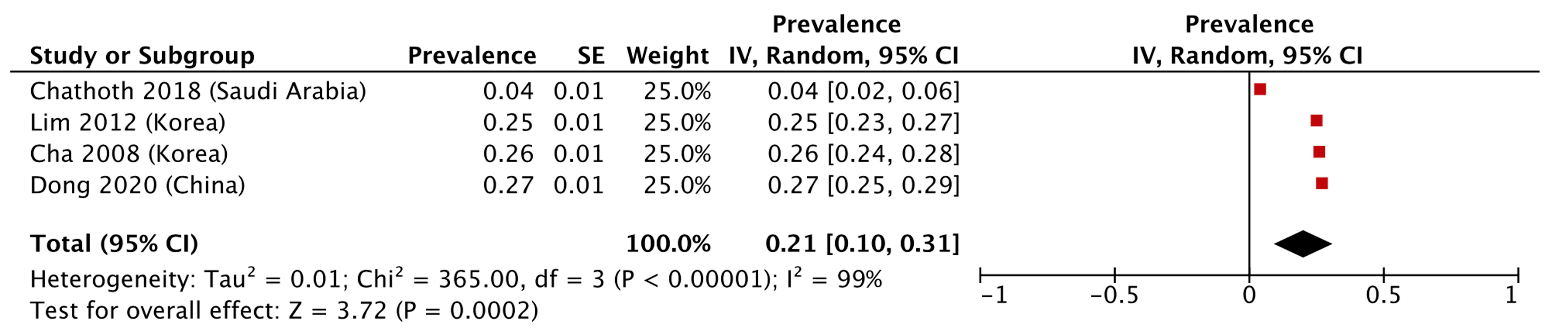

**Figure S40:** Forest plot for prevalence of *UCP1* A-1766G / G allele in CMP individuals.

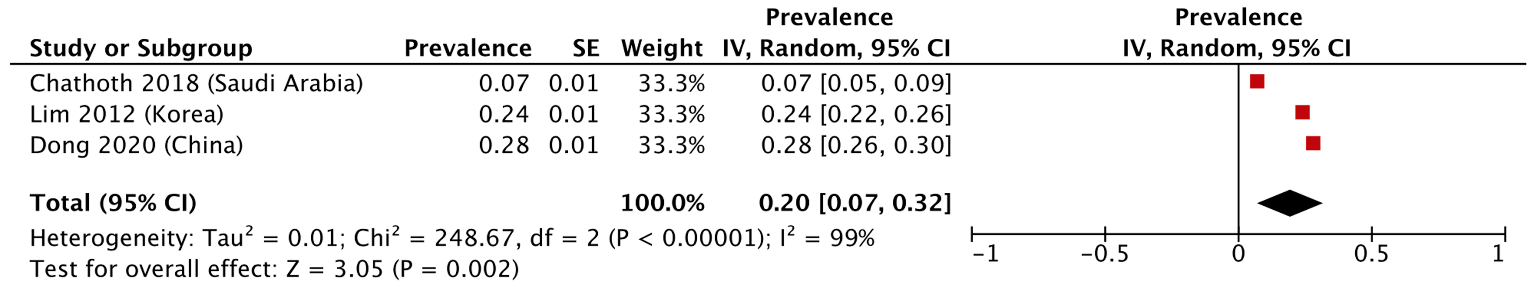

**Figure S41:** Forest plot for prevalence of *UCP1* Ala64Thr / A allele in healthy individuals.

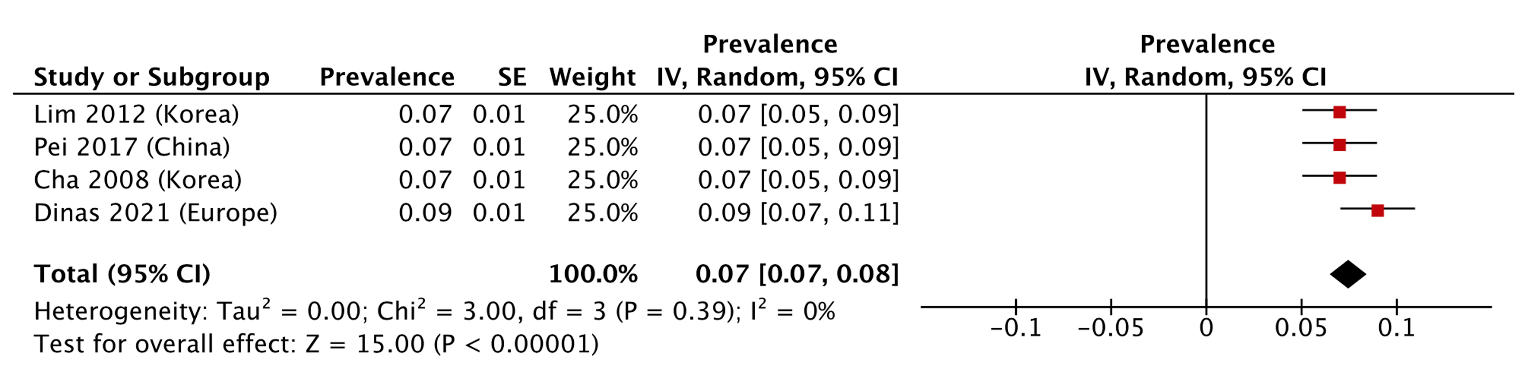

**Figure S42:** Forest plot for prevalence of *UCP1* Ala64Thr / A allele in CMP individuals.

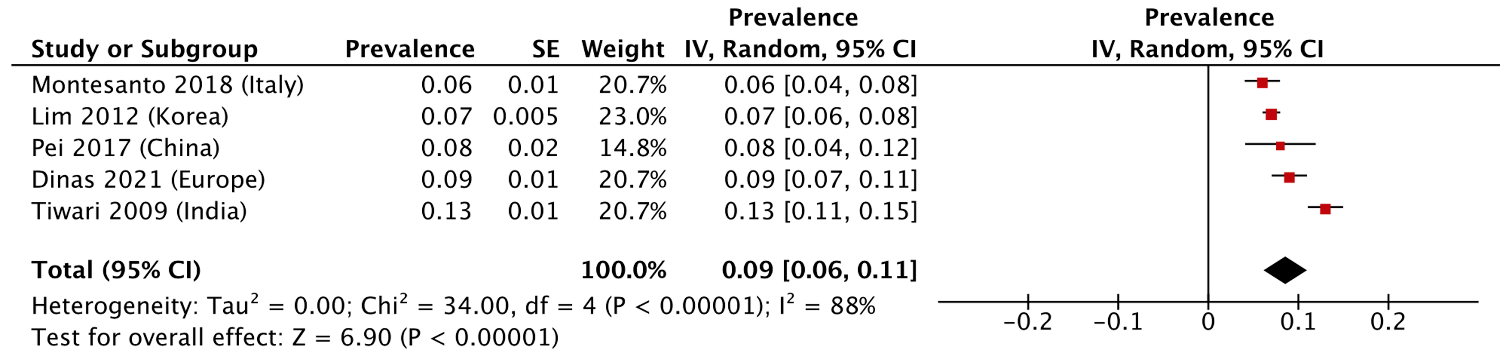

**Figure S43:** Forest plot for prevalence of *UCP1* A-112C / C allele in healthy individuals.

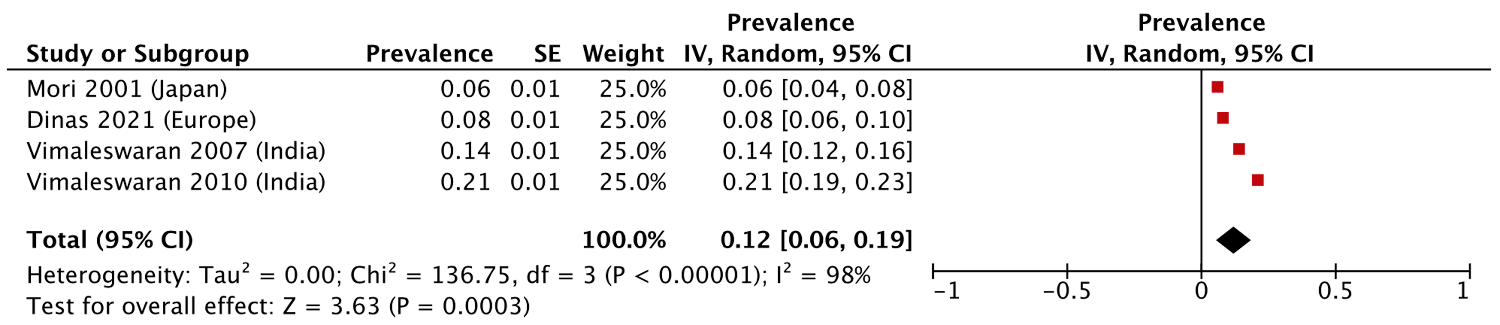

**Figure S44:** Forest plot for prevalence of *UCP1* A-112C / C allele in CMP individuals.

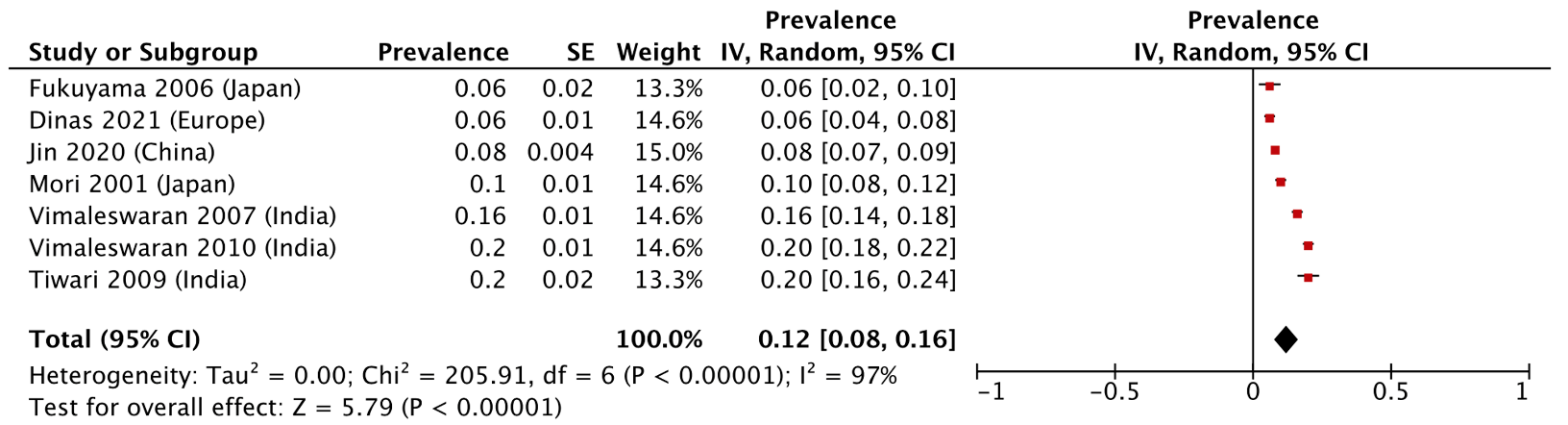

**Figure S45:** Forest plot for odds ratio of *UCP1* A-3826G / G allele

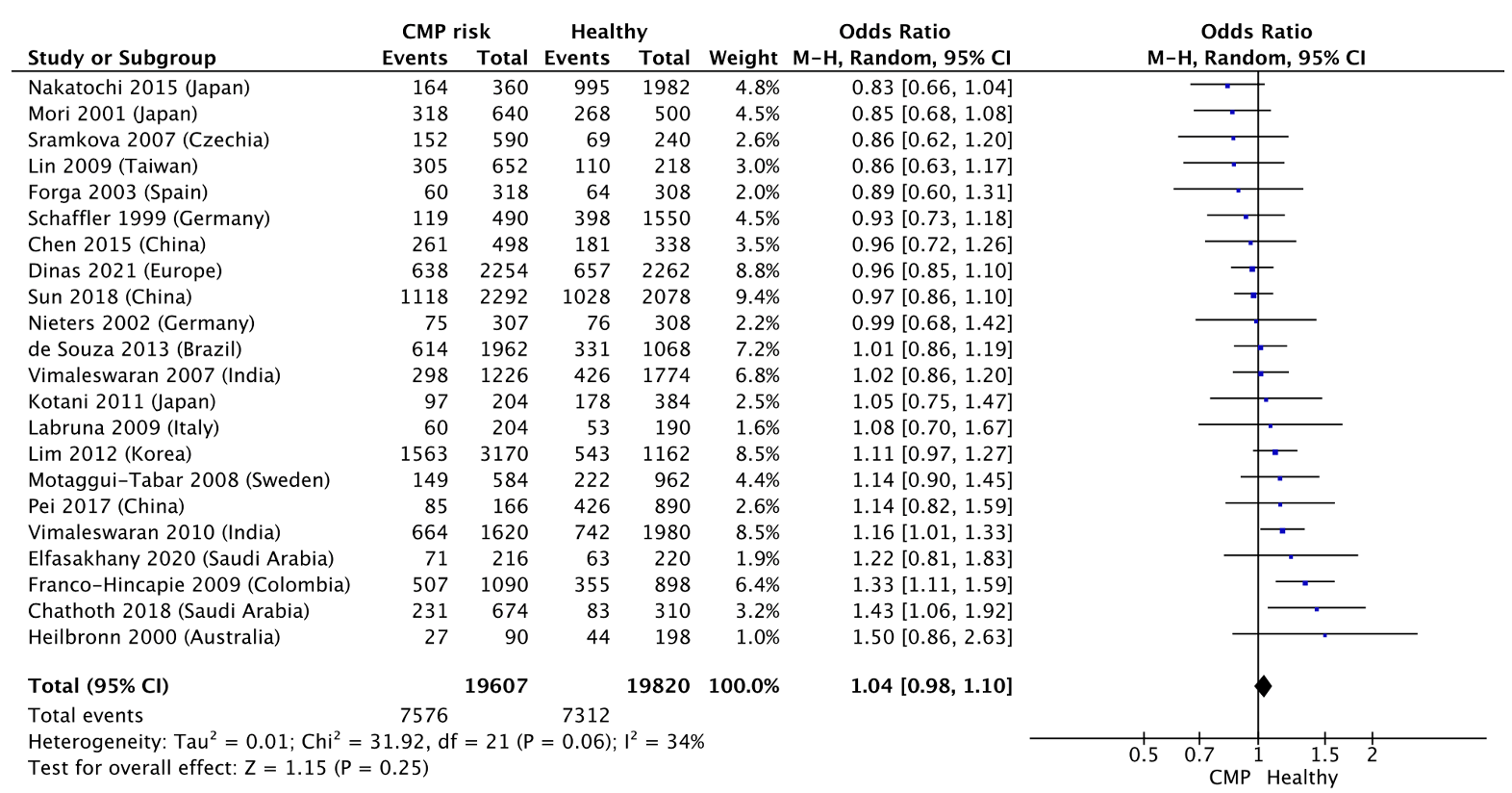

**Figure S46:** Funnel plot for odds ratio of *UCP1* A-3826G / G allele

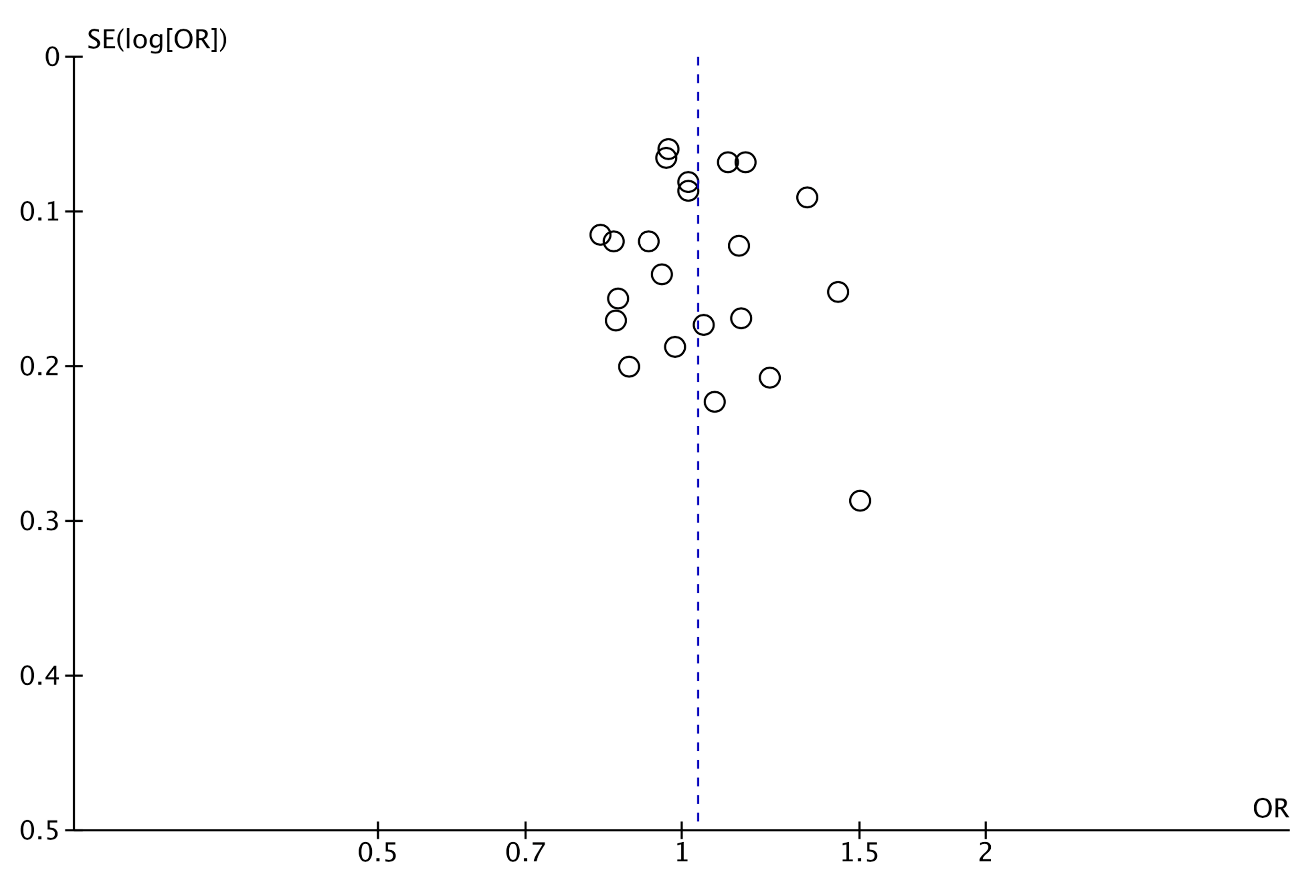

**Figure S47:** Forest plot for odds ratio of *UCP1* A-1766G / G allele

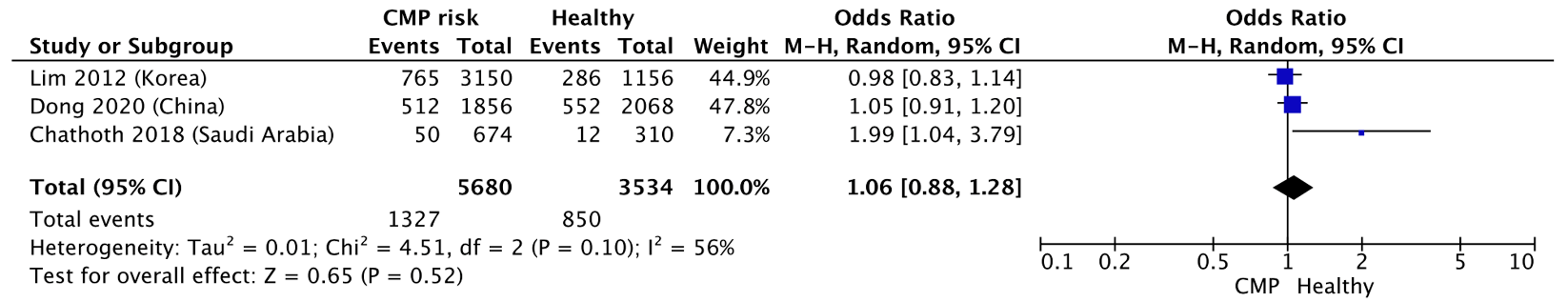

**Figure S48:** Forest plot for odds ratio of *UCP1* Ala64Thr / A allele

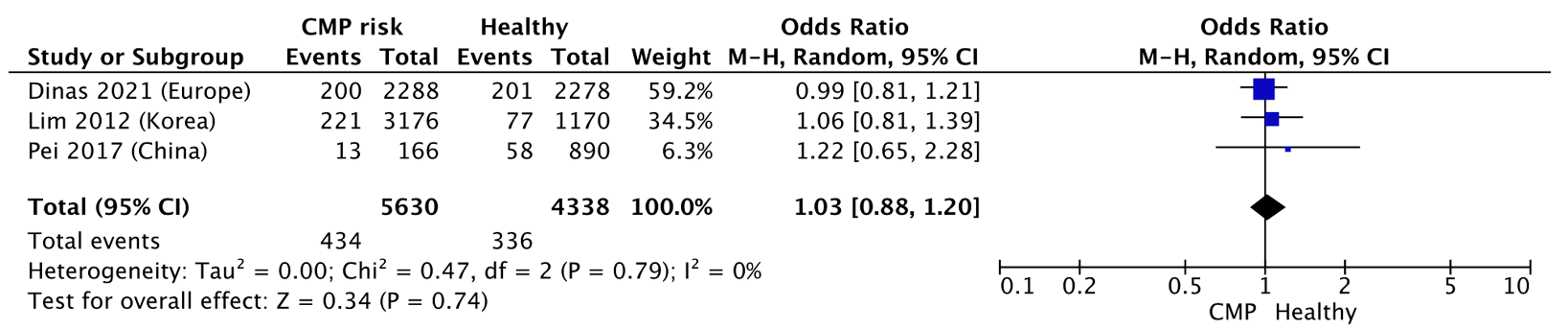

**Figure S49:** Forest plot for odds ratio of *UCP1* A-112C / C allele

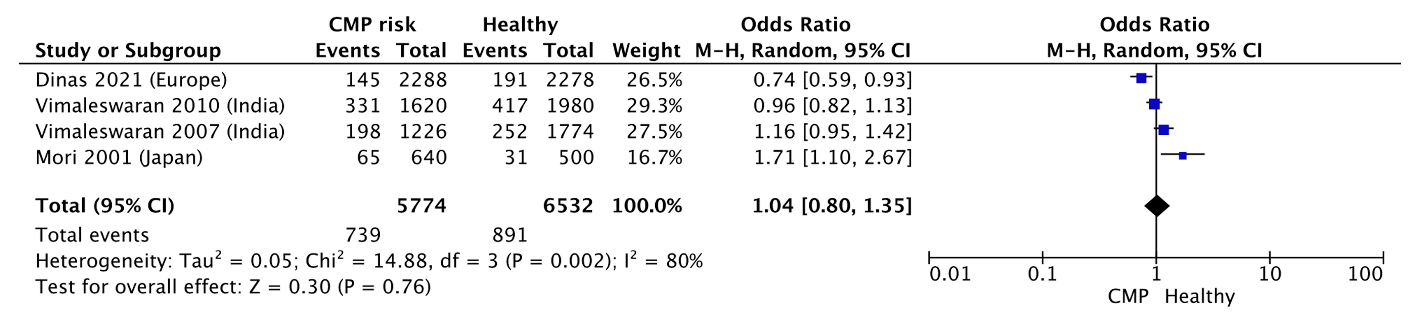
